# Age-aware genotype–phenotype architecture across 87 genetic neurodevelopmental disorders

**DOI:** 10.64898/2025.11.29.25341264

**Authors:** Ludovica Montanucci, Tobias Brünger, Christian M Boßelmann, Alina Ivaniuk, Samden Lathoo, Andres Jimenez-Gomez, Andreas Brunklaus, Christos Papadelis, Guo-Qiang Zhang, Wendy K. Chung, Costin Leu, Dennis Lal

## Abstract

Neurodevelopmental disorders (NDDs) are clinically heterogeneous, multisystem disorders, making prognosis challenging, even after a molecular diagnosis is obtained. In this study, we assembled a harmonized, age-aware genotype–phenotype atlas using data from the Simons Searchlight cohort. We analyzed 1,210 probands with pathogenic variants across 87 NDDs (75 gene, 12 recurrent copy-number variant disorders). Mapping 168 phenotypes onto 14 top-level Human Phenotype Ontology (HPO) categories showed that virtually all NDDs exhibited broad, multisystem involvement. In contrast, higher-resolution phenotype analyses uncovered 49 significant gene–phenotype enrichments (Bonferroni P < 3.9×10⁻⁵), for example, microcephaly in *DYRK1A*, macrocephaly in *PPP2R5D*, and distinct seizure-subtype patterns in *STXBP1* and *SLC6A1*. We calculated 1,368 age-dependent penetrance trajectories across 36 disorders and 139 phenotypes and use Cox models to identify 43 NDD-specific onset pattern (e.g., six times earlier absence seizures in *SYNGAP1* compared to other NDDs). Here we show that combining calibrated prevalence with developmental timing converts genetic findings into quantitative, gene-specific risk profiles. This atlas enables earlier recognition, gene-informed surveillance, and more rational trial design. The atlas is available for interactive exploration in the NDD-Portal (https://lalresearchgroup.org).

## Main text

Neurodevelopmental disorders (NDDs) arise from disruptions during key periods of brain maturation and affect multiple developmental domains, including cognition, motor coordination, language, behavior, and, in many individuals, seizure susceptibility. Rare genetic variants - including pathogenic single-gene variants and recurrent copy-number variants (CNVs) - represent the most common identifiable cause of NDDs. Advances in genomic sequencing have now implicated hundreds of genes in autism spectrum disorder and more than a thousand in epilepsies and related NDDs, alongside well-established, high-impact CNVs such as 16p11.2, 15q11.2–13, and 1q21.1^1–4^. As a result, genomic testing has become integral to the diagnostic evaluation of children with unexplained NDDs, yielding a molecular diagnosis in ∼20–40% of cases^5^, which is expected to rise as more comprehensive approaches such as long-read genome-sequencing, are adopted. These advances have created new opportunities for genetic data informed clinical care, including targeted surveillance for syndrome-specific complications and the development of emerging mechanism-based therapies^6,7^.

Despite the diagnostic revolution, leveraging genetic results for prognosis remains far more challenging than establishing a diagnosis. For most NDD-associated genes and CNVs, the full spectrum of associated phenotypes, their prevalence, and, critically, their age-dependent emergence remains incompletely characterized. Many symptoms overlap across NDDs, comorbidities are common, and individual genes typically account for <1% of children within broad diagnostic categories such as epilepsy, autism or intellectual disability. Moreover, penetrance and expressivity often evolve over a lifetime, and existing knowledge largely derives from anecdotal case series or highly enriched clinical cohorts. As a result, even when a pathogenic variant is identified, clinicians frequently lack actionable, i.e. calibrated and age-aware, information to guide anticipatory care, timing of evaluations, or surveillance for gene-specific complications.

Ongoing large-scale efforts such as the Deciphering Developmental Disorders (DDD) study and the Simons Searchlight cohort have identified tens of thousands patients with NDDs^8^ and annotated symptoms using Human Phenotype Ontology^9^ (HPO) terms^10^. However, most analyses of these data have emphasized gene discovery, similarity scoring, or clustering of related syndromes rather than generating calibrated prevalence estimates that compare and contrast phenotypes across the broad spectrum of NDDs. Population-level analyses of phenotypic overlap have provided valuable qualitative insights^11^, but a quantitative, age-aware framework that defines both the common features shared across NDDs and the gene-specific phenotypic signatures is still missing. Without such a framework, a fundamental knowledge gap about the variable clinical expressivity across NDDs remains and in clinical practice physicians lack quantitative priors for variant interpretation and cannot fully exploit the predictive and prognostic potential of genetic test results.

Although large patient data registries and EHR-linked resources are expanding rapidly^12–14^, including for NDDs, genotype-phenotype data remain fragmented across cohorts and annotation systems, limiting harmonized longitudinal inference and clinical utility. Building on the unique Simons Searchlight cohort (Simons Foundation Autism Research Initiative), which provides one of the largest and most deeply phenotyped collections of individuals with rare genetic NDDs, we developed a comprehensive knowledgebase for genetic NDDs. With this resource, we performed a large-scale, systematic, longitudinal characterization of genetic NDDs to address key knowledge gaps, including the incomplete definition of the phenotypic spectrum for most genetic NDDs, the limited understanding of phenotype prevalence within each condition, and the challenge posed by incomplete penetrance and age-dependent onset of clinical features. In this study, we analyzed data from more than 1,000 individuals in the Simons Searchlight cohort (version 13)^15^, encompassing pathogenic variants in 75 genes and 12 recurrent CNVs. We derived cross-condition trajectories of neurodevelopmental features, quantifying their age of onset and frequency to delineate gene-specific patterns of phenotypic evolution. By converting qualitative impressions into calibrated, data-driven frequencies and trajectories, our study delineates the shared multisystem architecture, distinct phenotypic signatures, and developmental timing profiles across genetic NDDs.

To support clinical translation, all results are made available through the interactive NDD-Portal (https://lalresearchgroup.org; Resources ‘NDD-Portal’), a clinician-facing atlas that enables exploration of phenotype prevalence, temporal emergence, and comparative trajectories across genetic NDDs. This integrative evidence-based atlas can support early recognition, anticipatory guidance, gene-informed surveillance, and precision-medicine trial design.

## Results

### Overlap, Timing, and Prevalence: Mapping Phenotypes Across Genetic Disorders in NDDs

To clarify the extent to which neurodevelopmental disorders share common phenotypic architectures versus exhibit gene- or CNV-specific patterns of penetrance and age-dependent expression, we systematically quantified the prevalence and temporal dynamics of 168 phenotypes across 87 genetic conditions. After mapping phenotypic categories to HPO terms, the hierarchical structure of the HPO allowed us to group the 168 phenotypes into 14 HPO terms which are direct children of the highest HPO term ‘Phenotypic abnormality - HP:0000118’ and which we call top-level HPO-terms.

Across the pooled cohort of 1210 probands, phenotypes mapped broadly across the 14 first-level HPO terms (Fig. 1A). Abnormality of the musculoskeletal system was the most prevalent term (81%), closely followed by Abnormality of the nervous system (68%). Digestive, ocular, endocrine, respiratory, and growth-related abnormalities were each present in roughly one-third to two-thirds of probands, underscoring the multisystem involvement typical of genetic NDDs. Cardiovascular, genitourinary, and immune system abnormalities were less common but still observed in >15% of the cohort. By contrast, integumentary findings, constitutional symptoms, and blood/blood-forming abnormalities were rare (<2%), illustrating that some major HPO categories contribute minimally to the phenotypic burden in this dataset. Notably, among individuals without any reported neurological phenotype, 42% carried a CNV.

**Fig. 1.**
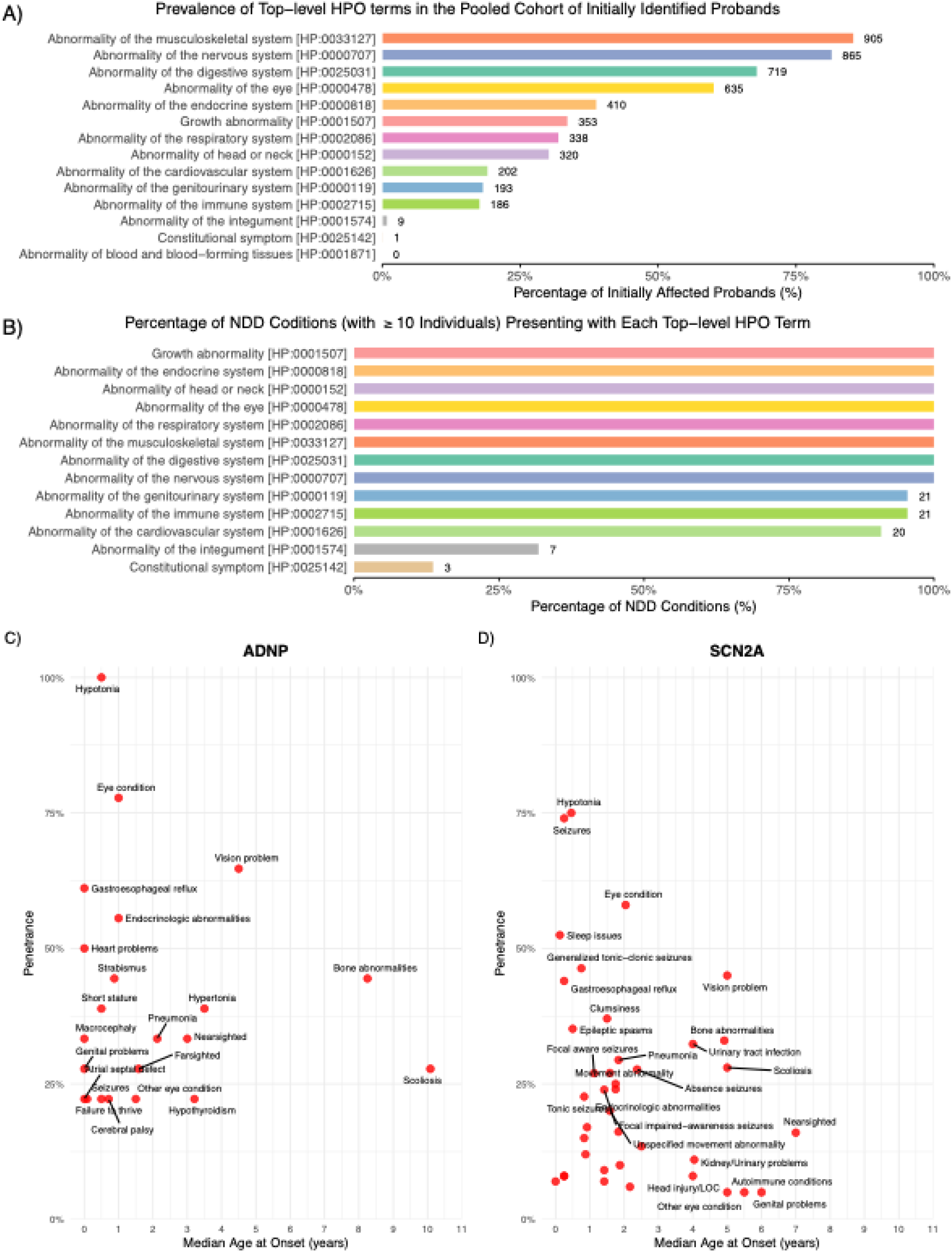
Landscape, shared architecture, and temporal dynamics of phenotypes across genetic NDDs. **A)** Prevalence of each top-level HPO term among initially affected probands across all neurodevelopmental disorders (family members excluded; N = 1210). Bars indicate the proportion of probands exhibiting at least one phenotype within each term. **B)** Percentage of genetic NDDs in which at least one individual presented a phenotype within each HPO term. This analysis was restricted to genetic conditions represented by ≥10 individuals to ensure stable estimates. Together, **A** and **B** highlight that most top-level HPO categories are broadly shared across conditions, with only a few exhibiting condition-specific representation. **C**-**D,** Penetrance and age-of-onset profiles for two representative conditions, *ADNP*- (**C**) and *SCN2A*-associated NDDs (**D**). Panels display the penetrance of each phenotype (y-axis; proportion of affected individuals) plotted against its median age of onset (x-axis) for all phenotypes reported in ≥3 individuals. The profiles illustrate marked gene-specific heterogeneity in both phenotypic frequency and developmental timing. ADNP-associated NDD shows high penetrance with predominantly early onset for hypotonia, eye conditions, and gastroesophageal reflux, whereas SCN2A-associated NDD is characterized by a substantially higher and earlier seizure burden alongside more frequent early-onset sleep and movement abnormalities. Shared features such as eye or gastrointestinal symptoms appear in both conditions but differ in penetrance and onset patterns. Together, these maps highlight the distinct phenotypic trajectories that emerge at the level of individual genes despite overlapping involvement of broad HPO systems.

When examining how these HPO systems are distributed across genetic conditions represented by at least 10 individuals, we observed a striking degree of cross-condition occurrence: 8 of the 14 top-level categories appeared in every such condition (Fig. 1B). Growth, endocrine, head/neck, eye, respiratory, musculoskeletal, digestive, and nervous system abnormalities were all universally represented. Cardiovascular and immune system features were also frequent, present in 20–21 of the 21 conditions meeting the sample size threshold. Only integumentary and constitutional categories showed marked condition-specificity, identified in 7 and 3 conditions respectively. Together, these patterns indicate that while the specific phenotypes driving each HPO term may differ, the presence of multisystem involvement is ubiquitous across genetic NDDs, reflecting a highly overlapping phenotypic architecture at the system level.

Despite the broad overlap of top-level phenotypic categories across conditions, substantial heterogeneity emerged when examining penetrance and age of onset for individual phenotypes (Fig. 1C–D). For example, among individuals with *ADNP*-associated NDD, hypotonia, eye conditions, and gastroesophageal reflux exhibited high penetrance with predominantly early childhood onset, whereas many other features - including hypertonia and bone abnormalities - showed more variable penetrance and later onset patterns. In contrast, *SCN2A*-associated NDD displayed a markedly different profile: seizures occurred with much higher penetrance and significantly earlier onset compared to *ADNP*, and early-onset sleep issues, generalized tonic–clonic seizures, and several movement-related abnormalities were also more common. Across both disorders, certain phenotypes—such as eye conditions and gastrointestinal symptoms—were shared but differed in frequency or developmental timing, illustrating that even overlapping features can follow distinct temporal trajectories. Other phenotypes showed condition specificity: for instance, several movement-abnormality terms appeared predominantly in *SCN2A*, whereas some endocrine-related and growth-related abnormalities were more characteristic of *ADNP* (Fig. 1C–D). Together, these examples illustrate how individual NDDs diverge in both the severity and developmental timing of specific phenotypes, despite belonging to a broadly shared multisystem phenotype architecture. A comprehensive map of the specific phenotype profile of penetrance and age of onset for all 87 genes and CNVs is provided in Supplementary Fig. 1, with two representative examples highlighted in Fig. 1C-D.

### Enrichment of Phenotypes in Individual Genetic NDDs Reveals Phenotypic Signatures

To identify phenotypic features that distinguish individual genetic NDDs, we performed a gene–phenotype enrichment analysis comparing the prevalence of each phenotype within a given condition to its prevalence across all other 87 NDDs. Among 1,294 comparisons, 49 associations met Bonferroni-corrected significance (P < 3.9 × 10⁻⁵), revealing 44 enriched and 5 depleted phenotype–condition pairs (Fig. 2). Although broad HPO systems were shared across most conditions (Fig. 1), this analysis identified a focused set of high-confidence, condition-specific clinical signatures. Several of the strongest enrichments involved cranial growth abnormalities, including microcephaly in *DYRK1A* and macrocephaly in *PPP2R5D*, reflecting well-established gene-specific neurodevelopmental patterns. Epilepsy-related phenotypes also featured prominently: *STXBP1*-associated NDD showed significant enrichment for seizures overall, while *SLC6A1* demonstrated strong association for specific seizure subtypes. Beyond neurological features, enriched phenotypes spanned musculoskeletal, ophthalmologic, endocrine, and gastrointestinal systems, each mapping onto discrete genetic conditions. A minority of disorders exhibited phenotype depletion, suggesting distinguishing clinical features that may aid differential diagnosis. Collectively, these enrichment results demonstrate that only a small subset of gene–phenotype pairs show strong, condition-specific prevalence against a background of widespread multisystem involvement. These robust phenotypic “fingerprints” provide clinically actionable insights that can refine diagnostic evaluation, support gene-informed prognostication, and guide targeted surveillance across neurodevelopmental disorders.

**Fig. 2.**
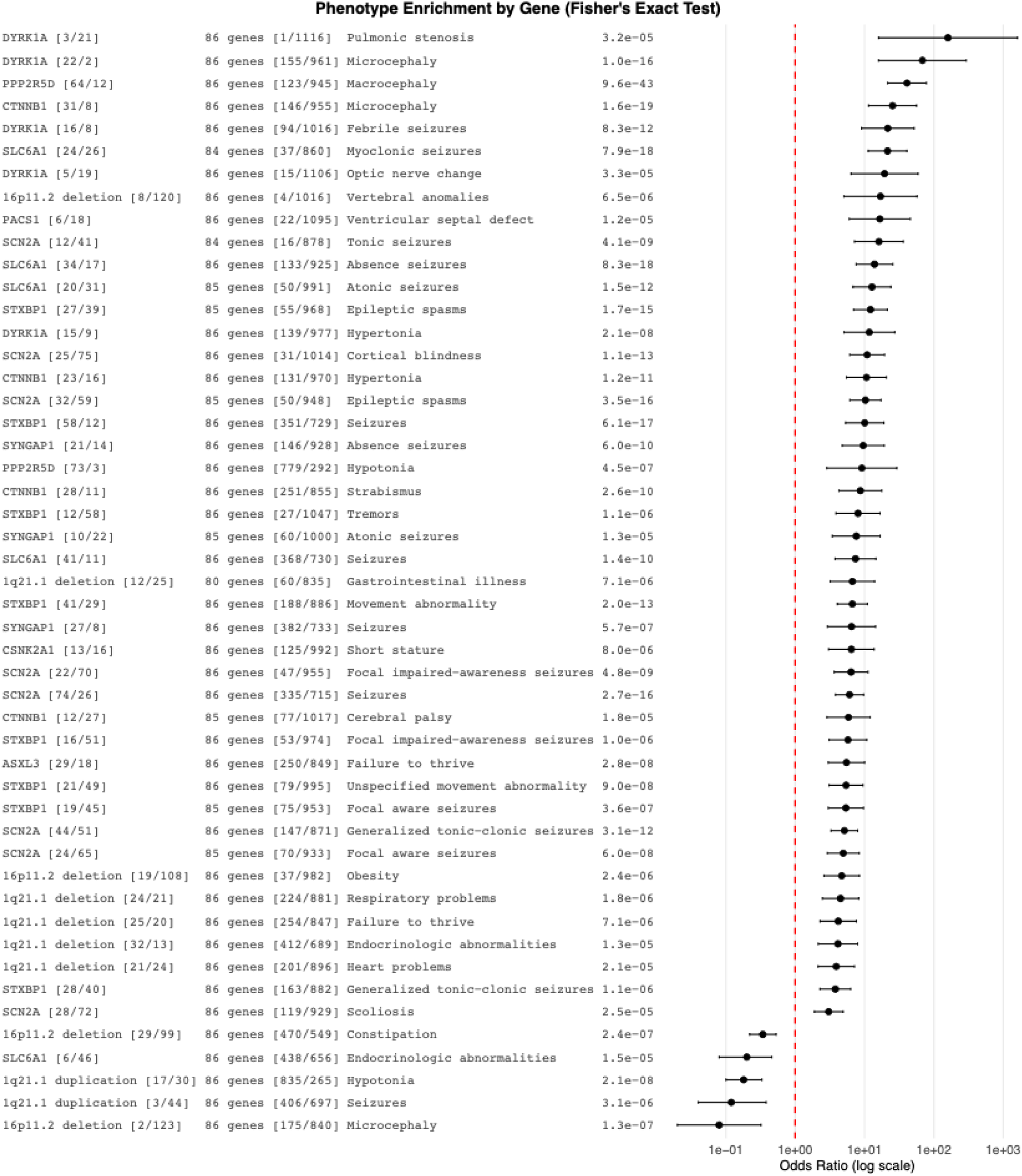
Gene-specific phenotype enrichment highlights high-risk associations. Forest plot displaying odds ratios (ORs, log scale) with 95% confidence intervals for all significant gene–phenotype enrichments identified across 1,294 gene–phenotype comparisons (Fisher’s exact test, Bonferroni-corrected P < 3.9 × 10⁻⁵). Each row represents a phenotype showing significantly higher or lower prevalence among individuals with a given genetic NDD compared with all other genetic NDDs. The left table lists the gene or CNV (with sample size), comparator cohort size, phenotype, and raw P value. Enriched phenotypes include highly specific associations such as microcephaly with *DYRK1A*, macrocephaly with *PPP2R5D*, and multiple seizure subtypes with *SCN2A* and *SLC6A1*, illustrating distinct phenotypic signatures across genetic NDDs. Only 49 associations met genome-wide multiple-testing correction, underscoring both the specificity and rarity of strong phenotype enrichments within the context of broadly overlapping multisystem involvement.

### Age-Dependent Penetrance Uncovers Clinically Actionable Variation Among Genetic NDDs

To support earlier recognition and anticipatory management, it is essential to understand not only whether a phenotype is associated with a genetic NDD but also when it typically emerges during development. While the preceding analyses revealed extensive overlap in first-level HPO terms and identified phenotype-level enrichments specific to individual genes, developmental timing represents a further axis of heterogeneity with direct clinical implications. We therefore constructed age-dependent penetrance trajectories for each phenotype and genetic NDD, estimating the cumulative proportion of individuals reported to manifest a phenotype by each age. These curves reflect documented onset (rather than latent biological onset) and thus capture the timing patterns that clinicians and caregivers encounter. To ensure stability, we included only gene–phenotype combinations with ≥20 individuals assessed and overall penetrance >1%, yielding 1,045 trajectories across 19 genetic NDDs and 121 phenotypes (Supplementary Fig. 2). Differences in follow-up age and cohort size are shown alongside each trajectory (Fig. 3 histograms) to contextualize curve slopes across conditions. Fig. 3 highlights twelve representative trajectories for *STXBP1, DYRK1A*, and *ASXL3*, chosen for their distinct and clinically recognizable developmental profiles. Four phenotypes of high clinical relevance—seizures, hypotonia, hypertonia, and failure to thrive—illustrate how overall penetrance and developmental timing diverge across genes. Although seizures (Fig. 3A) are common in both *STXBP1* and *DYRK1A*, their timing differs substantially. *STXBP1* shows a steep early rise, with ∼30% affected by 6 months, whereas seizure risk in *DYRK1A* accumulates more gradually across early childhood. *ASXL3* shows minimal seizure involvement. These differences offer practical discriminatory value in infants presenting with early-life seizures. Meanwhile, hypotonia (Fig. 3B) is highly penetrant and typically early-onset in *ASXL3* and *STXBP1*, but less common in *DYRK1A*. Although onset occurs early across all three conditions, their differing overall prevalence provides clinically useful differentiation during early developmental assessment. Conversely, hypertonia (Fig. 3C) is most prominent and earliest in *DYRK1A*, consistent with its known motor phenotype. *STXBP1* and *ASXL3* show lower and later hypertonia, distinguishing *DYRK1A* in children presenting with early motor stiffness or mixed tone abnormalities. Lastly, for failure to thrive (Fig. 3D), *ASXL3* demonstrates the highest early-life penetrance (>60% in the first year), *STXBP1* shows low early involvement (<10%), and penetrance across some domains in *DYRK1A* increases gradually throughout childhood. Together, these gene-specific trajectories reveal a multidimensional structure of phenotypic expression across NDDs—frequency, specificity, and developmental timing—that is not captured by system-level prevalence alone. Clinically, these timing profiles provide actionable signals: they can sharpen early diagnostic suspicion and inform tailored anticipatory care across infancy and childhood. The complete set of 1,368 trajectories across 36 NDDs and 139 phenotypes is provided in Supplementary Fig. 2.

**Fig. 3.**
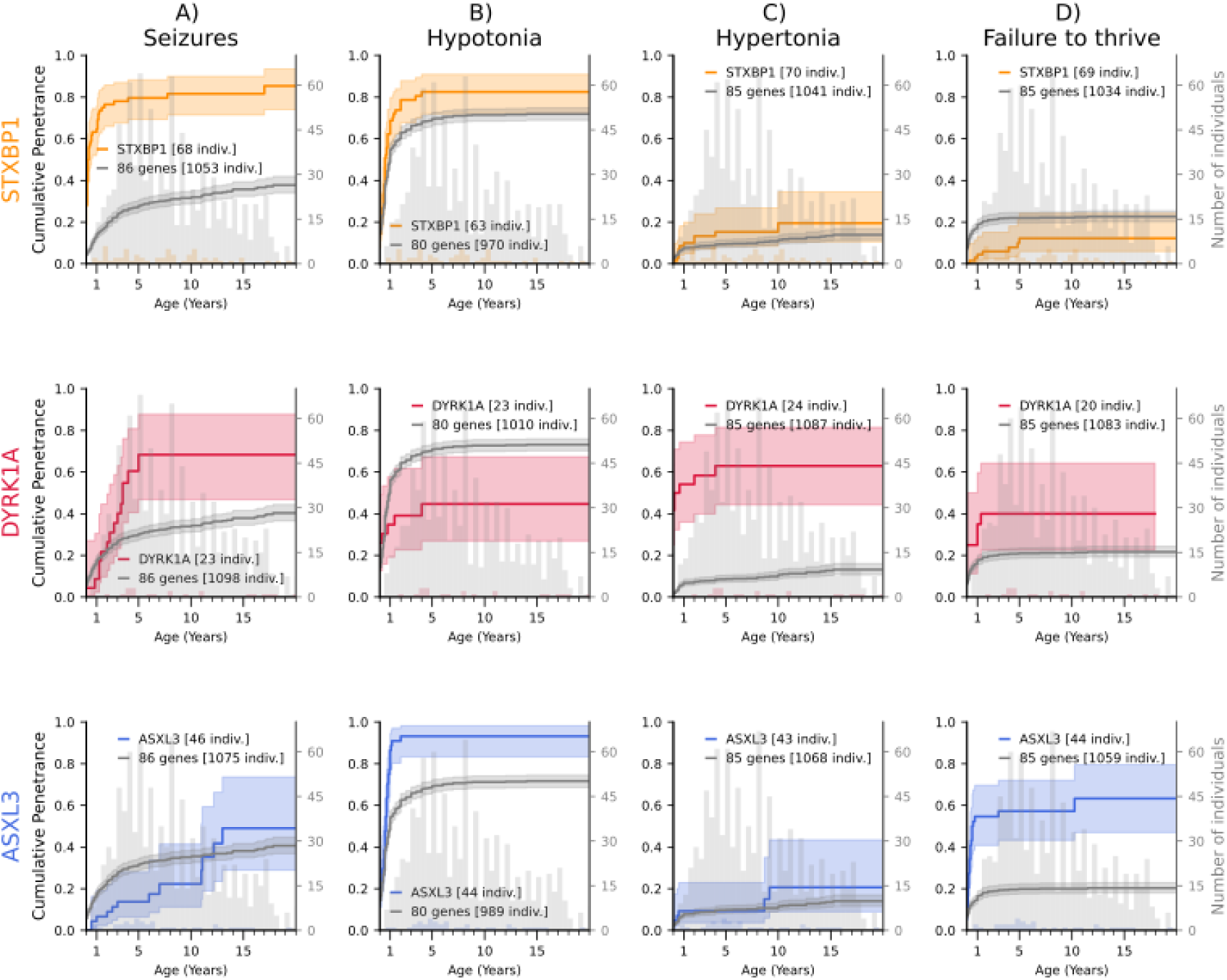
Age-dependent penetrance trajectories reveal clinically meaningful differences in symptom timing across genetic neurodevelopmental disorders. Cumulative penetrance trajectories are shown for three genetic NDDs - *DYRK1A-*, *ASXL3*-, and *STXBP1*-related disorders - across four phenotypes: seizures (A), hypotonia (B), hypertonia (C), and failure to thrive (D). Curves represent the age-dependent proportion of individuals manifesting each phenotype, illustrating when symptoms typically emerge during development. Colored lines depict gene-specific trajectories, and gray lines show pooled reference curves from all other NDDs. Histograms on the right y-axis display the distribution of the last available age of assessment for each gene (violet) and the reference group (gray), indicating follow-up coverage. Distinct gene-specific patterns in both overall penetrance and age of onset are evident. *STXBP1* shows rapid early-life accumulation of seizures and hypotonia, suggesting that these features may support early diagnostic suspicion. *ASXL3* demonstrates high early penetrance of hypotonia and failure to thrive, while *DYRK1A* is characterized by a more gradual rise in seizures and prominent early hypertonia. These trajectories highlight actionable differences in developmental timing that can inform anticipatory guidance, targeted surveillance, and gene-informed clinical management. The full set of 1,368 trajectories for 36 NDDs and 139 phenotypes is provided in Supplementary Fig. 2.

### Identifying Early- and Late-Onset Phenotypes Across Genetic NDDs

While cumulative penetrance curves show when symptoms usually develop within an NDD, Cox hazard models can quantify whether those symptoms occur significantly earlier or later than expected relative to other NDDs. Clinically, this information helps tailoring anticipatory care by identifying which genetic condition need early screening versus long-term monitoring based on how soon key symptoms typically appear. When focusing specifically on differences in age of onset compared with the broader NDD population, several gene-phenotype pairs stood out as having early manifestations (Table 1). *PPP2R5D*-related macrocephaly showed the earliest onset overall (HR = 15.3), indicating head-size abnormalities that are often evident at or shortly after birth. *SYNGAP1* variants led to absence seizures over six times earlier than in other NDDs, while *STXBP1* showed similarly early emergence of movement abnormalities and seizures (HRs ≈ 6–7), emphasizing the need for early seizure monitoring. *DYRK1A* demonstrated early presentation of hypertonia. Together, these findings identify genetic conditions with early phenotypes, guiding clinicians toward gene-informed timing of surveillance and intervention.

**Table 1.**
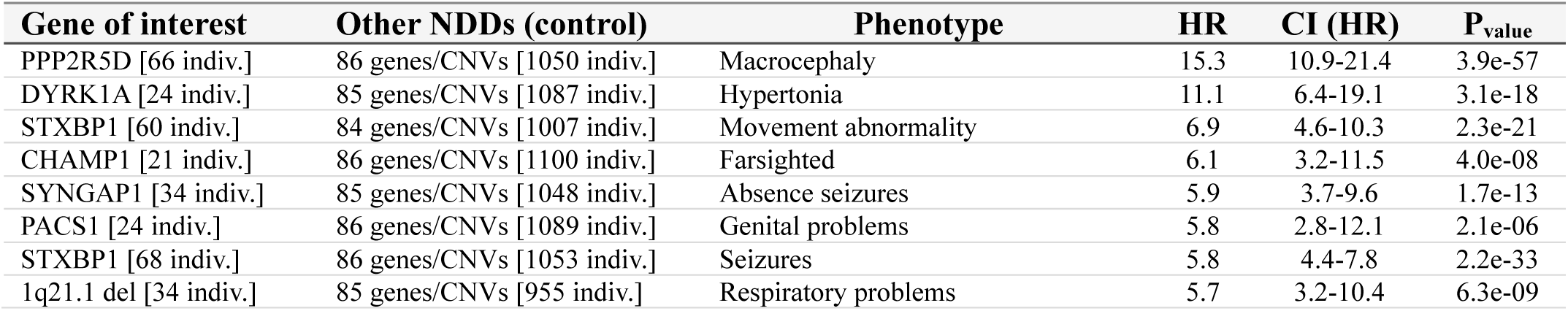
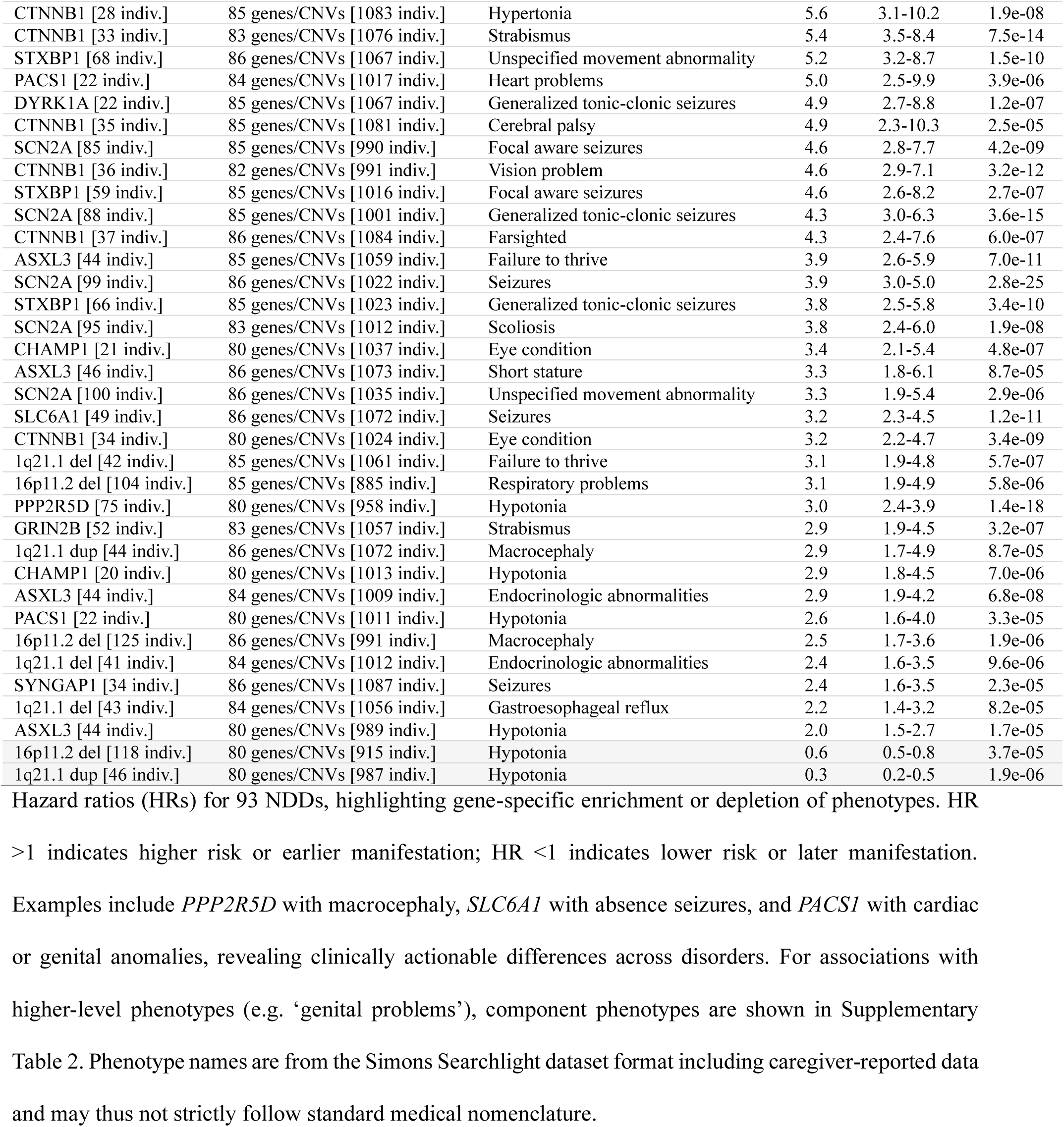
Genetic NDD-phenotype pairs with significantly earlier or later onset compared with the broader NDD cohort.

### The NDD-Portal: An Interactive Atlas of Genetic NDDs with Clinical Applications

To facilitate clinical translation of the genotype–phenotype landscape described above, we consolidated all trajectory, prevalence, and enrichment outputs into an interactive, publicly accessible resource (“NDD-Portal”). This portal represents, to our knowledge, the largest standardized phenotype atlas for genetic NDDs, enabling users to explore gene-specific phenotype profiles, age-dependent penetrance patterns, and comparative prevalence across 87 genes and CNVs (Fig. 4A). By integrating harmonized phenotype definitions with cumulative penetrance analyses, the portal transforms previously anecdotal, case-driven impressions of individual conditions into data-driven summaries that clinicians and researchers can query in real time.

**Fig. 4.**
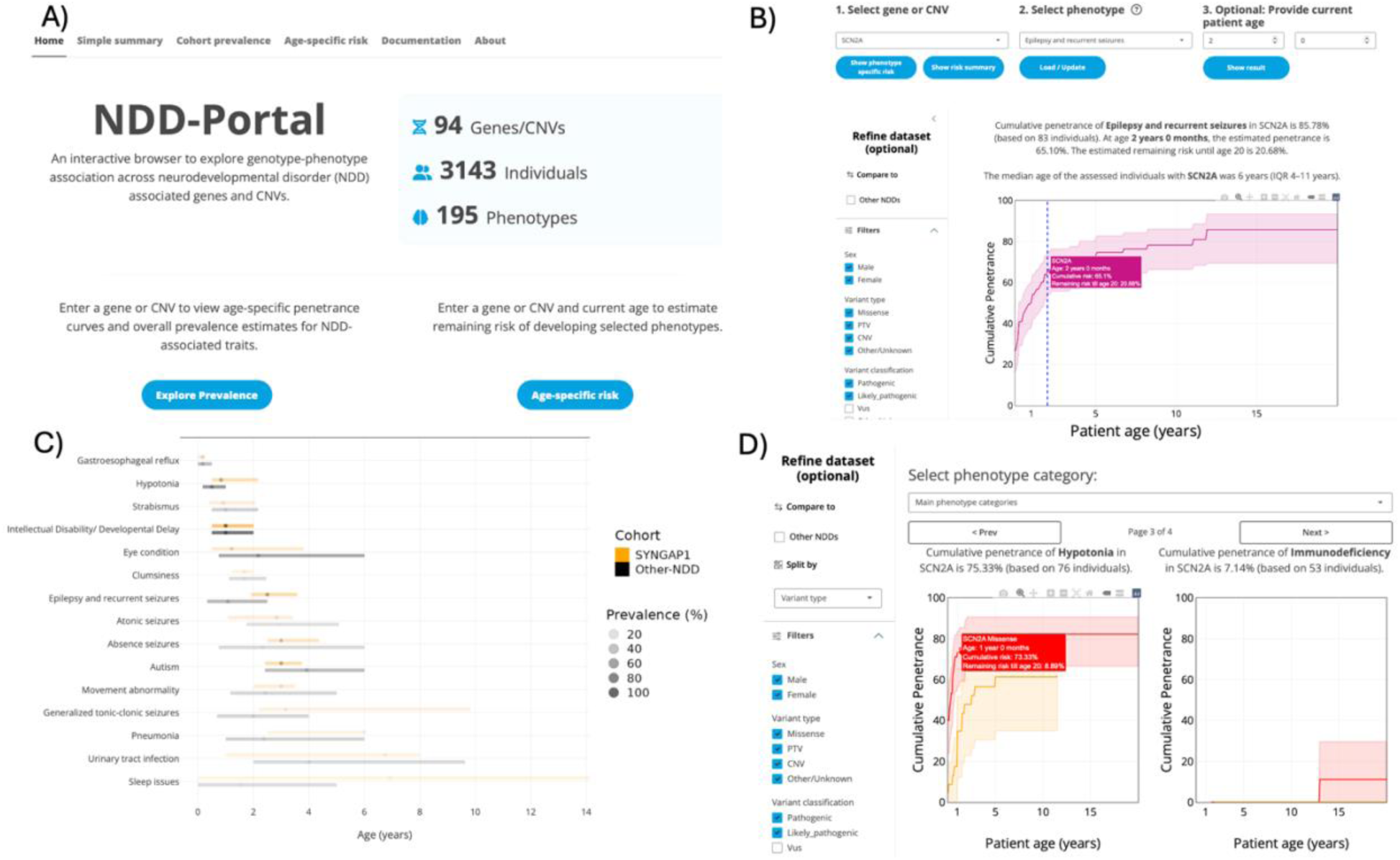
NDD-Portal: Interactive atlas of genetic NDDs with clinical applications. (**A**) Homepage of the NDD-Portal web-server. (**B**) Cumulative penetrance trajectory for seizures in *SCN2A*, providing age-dependent risk estimates. (**C**) Phenotype overview for SCN2A, showing the full spectrum of associated features and their prevalence. (**D**) Comparative cumulative penetrance curves for PTV versus missense variants in *SCN2A*. The portal enables clinicians and researchers to explore gene-specific phenotype profiles, quantify risk, and support evidence-based anticipatory care, monitoring, and counseling in pediatric NDDs.

The NDD-Portal provides three core functionalities. First, users can view the full spectrum of documented phenotypes for each genetic NDD, along with their prevalence and confidence intervals, allowing rapid visualization of system-level and phenotype-level involvement (Fig. 4B). Second, for each phenotype, the portal displays its age-dependent penetrance trajectory, reflecting the proportion of individuals reported to manifest that phenotype by each age (Fig. 4C-D). These curves convey reported developmental timing, not true biological onset, and therefore contextualize how symptoms are documented in heterogeneous clinical settings. Third, users may compare trajectories across variant classes or NDDs when sample sizes permit, illustrating differences in timing or overall penetrance—for example, cumulative seizure trajectories for *SCN2A* protein-truncating versus missense variants (Fig. 4C–D). Importantly, the portal does not generate individualized prognoses or predict outcomes for single patients. Instead, it provides population-level estimates with confidence intervals, suitable for benchmarking gene-specific patterns and informing discussions about potential trajectories rather than precise forecasts. This is particularly relevant for early-onset phenotypes, where reporting may be incomplete, and for less common phenotypes that have wide uncertainty bounds. For example, a clinician viewing cumulative seizure curves for *SCN2A* can see that seizures are frequently reported in infancy, but the portal explicitly displays confidence intervals and sample sizes, signaling that risk estimates become more uncertain at older ages or when stratified by variant class. Together, the NDD-Portal enables evidence-based exploration of gene-specific phenotype patterns, supports more informed anticipatory guidance, and allows researchers to examine population-level developmental trajectories across diverse genetic NDDs. The resource is intended as a decision-support and discovery tool, complementing, not replacing, individualized clinical assessment.

## Discussion

Neurodevelopmental disorders caused by rare genetic variants are clinically heterogeneous and often involve multiple systems. By harmonizing longitudinal phenotypes across genes and recurrent CNVs and applying time to event analyses, we provide the first age-aware atlas of genotype-phenotype relationships that translates molecular diagnoses into developmental risk profiles across 87 NDDs. We demonstrate that, at the level of top-level HPO systems, multisystem involvement is common across conditions^16^; however, at the level of specific phenotypes and their timing, disorders differ significantly. This combination of broad system overlap with gene specific penetrance and age of onset explains why cross-sectional feature lists have limited discriminative value and motivates timing-aware risk estimates^17^.

Three findings stand out. First, a focused set of gene-phenotype pairs shows statistically robust enrichment or depletion against a background of widespread multisystem involvement, yielding clinically recognizable “phenotypic signatures” (e.g., cranial growth abnormalities and specific seizure subtypes in defined genes/CNVs). Second, cumulative penetrance curves differ in both slope and ceiling across genes; these differences carry practical value for anticipatory care. Third, hazard-based comparisons identify phenotypes that are documented substantially earlier or later than expected relative to other NDDs (e.g., early seizures and movement abnormalities in *STXBP1*, early macrocephaly in *PPP2R5D*, early failure to thrive in *ASXL3*, helping in confirming diagnosis in case of novel variants, highlighting where early screening is most justified and where longer-term monitoring is warranted.

These age-aware outputs have immediate clinical applications. For example, an age-adjusted remaining seizure risk after a baseline EEG can guide the periodicity of subsequent EEGs and counseling about near term seizure likelihood. It can also guide in identifying individuals who require more aggressive seizure surveillance, and in delineating condition-specific growth curves to determine when medical complications should be investigated if growth deviates from expectations, in performing targeted imaging when structural anomalies are anticipated, and ensuring timely vision and hearing screening where appropriate.

Our resource complements existing compendia that primarily aggregate case descriptions or provide cross sectional prevalence. By converting heterogeneous clinical histories into age-indexed estimates with confidence intervals, we move from static checklists to quantitative, developmentally informed priors that can be examined gene by gene and phenotype by phenotype. To facilitate use, we provide an interactive NDD-Portal that displays prevalence, trajectories, and comparisons across relevant strata (e.g., variant classes where sample size allows). The portal reports group-level estimates and uncertainty and is not intended to predict individual outcomes. Analyses were restricted to individuals carrying pathogenic or likely pathogenic single gene variants or well established recurrent CNVs; variants of uncertain significance were excluded. These choices increase specificity for genotype-phenotype inference and align the outputs with clinical decision making after a confirmed molecular diagnosis.

Several limitations merit consideration. The Simons Searchlight cohort is research-ascertained and may not represent all care settings or ancestries. Many phenotypes are caregiver-reported and negative assessments are not necessarily clinical confirmed, which may inflate apparent penetrance for features that are actively queried and deflate others^18^. The SFARI project uses a two-layer hierarchical phenotype structure, where parent-level terms (e.g., ‘heart condition,’ ‘eye condition’) may lack the specificity needed to represent clinically actionable features. Furthermore, because our analysis is conducted at the gene level, genes with multiple mechanisms of action may encompass more than one underlying condition, which cannot be fully disentangled here. The long tail of very rare genes remains underpowered, and pooling heterogeneous disorders as a comparator can dilute disorder-specific baselines. A further limitation is our inability to disentangle whether certain phenotypes reflect the underlying genetic disorder, sporadic comorbidities, or treatment-related effects; for instance, osteoporosis may arise as a side effect of long-term antiseizure medications rather than the disorder itself. Future work could address this by mapping common medication-associated adverse effects to HPO terms and applying correction factors to down-weight these signals, particularly for phenotypes that emerge later in life. Time to event analyses assume non informative censoring and are sensitive to delayed entry/left truncation; moreover, curves reflect documented onset rather than true biological onset. These considerations argue for cautious interpretation of late age tails and of sparsely reported phenotypes.

Many of these constraints are addressable and shape our roadmap. We are prioritizing (i) prospective, clinician validated phenotyping with explicit capture of negative findings; (ii) federation across sites and linkage to EHRs to improve diversity and representativeness; (iii) variant level modeling (e.g., protein truncating vs missense variants, recurrent alleles, CNV size/orientation) where sample sizes permit; (iv) methodological refinements for delayed entry and competing risks; and (v) external replication in independent registries. Because all analyses are embedded in a living portal, updates can continuously improve precision, broaden gene coverage, and enable calibration for ancestry and health system context.

In sum, this work establishes the first cross-NDD age-aware, data-driven foundation for genotype-informed care in pediatric NDDs. By integrating prevalence with *when* features are typically documented, the atlas and portal support early recognition, more appropriately timed evaluations and surveillance, and more rational trial design while remaining explicit about uncertainty, scope, and the population level nature of the estimates.

## Online Methods

### Sample: Simons-searchlight Data Set

We conducted a retrospective cohort study using the Simons Searchlight research registry (2013–present), analyzing the lifetime dataset version 13. Participants (or caregivers) contributed standardized, web-based questionnaires across medical, developmental, behavioral, and family-history domains; earlier phases also used structured telephone interviews administered by trained staff. Data are longitudinal, allowing age-at-onset capture and repeated updates; all records were de-identified prior to investigator access. The project was approved by the Simons Foundation (Project ID: 15099.1) and the institutional review board at UTHealth Houston (IRBs: HSC-MS-24-0469 and HSC-MS-23-1129)

#### Genes and Variants

The Simons-searchlight *lifetime* dataset version 13 comprises 1,365 individuals with variants in one of 75 genes or 12 recurrent CNVs associated with neurodevelopmental conditions. Genetic results from clinical testing were abstracted and centrally reviewed by a Simons Searchlight genetic counselor. The list of the considered genes and CNVs, the number of individuals carrying variants in each of those, and the number of phenotypes presented by at least one individual carrying the variant are shown in Supplementary Table 1 in the Supplement. Individuals with no genetic variant (“Negative” genetic status) were not considered in the analysis. The number of variants in this dataset is 2,891, of which 2,048 (71%) are classified as pathogenic or likely pathogenic, and 312 (11%) as VUS. The remaining 524 (18%) have an unspecified variant or lack a classification. For all our analysis we only retained individuals with a pathogenic or likely pathogenic variant.

#### Phenotypes

Of the approximately 1,600 phenotypic fields annotated for each individual in the dataset, 209 correspond to disease-related phenotypes with recorded age of onset. From these, we selected 168 phenotypes relevant to neurodevelopmental disorders (NDDs), as detailed in Supplementary Table 2 in the Supplement. Developmental Delay/Intellectual Disability (DD/ID) was not captured in this survey and was therefore added following the procedure proposed in Stevelink et al ^19^.

#### Mapping of Searchlight Phenotypes into top-level HPOs

Of the approximately Searchlight phenotypes were used as input for RAG-HPO, a tool for automated analysis of clinical information using large language models with Retrieval Augmented Generation^20^. Further, two neurologists with an expert in both genetic neurodevelopmental disorders and computational phenotyping acted as independent raters and manually assigned an HPO term for each SFARI phenotype. Inter-rater agreement was assessed using Cohen’s Kappa for exact matches, and Lin’s semantic similarity to represent phenotypic similarity^21^. Human raters showed excellent agreement on exact HPO term selection (Cohen’s Kappa 0.885). When they disagreed, the terms chosen were still semantically very similar (Lin’s similarity 0.970). The automated approach using RAG-HPO had lower coverage (127/168 SFARI phenotypes mapped to HPO terms vs 164/168 and 167/168 for the two raters, respectively) and moderate exact agreement (Cohen’s Kappa 0.558-0.573) but still maintained high phenotypic similarity (Lin’s similarity 0.930-0.950). Disagreements were resolved by consensus. After consensus was reached, all Searchlight phenotypes were matched to HPO terms. After mapping, we assigned HPO terms to their top-level categories to broadly represent system-wide abnormalities. To do this, we propagated each HPO term until the nearest children of the node ‘Phenotypic abnormality’ (HP:0000118) was reached.

### Age-dependent Cumulative Penetrance Estimations

To characterize age-dependent penetrance of genotypes in the Searchlight dataset, we calculated cumulative penetrance curves, defined as the proportion of variant carriers who have developed a given phenotype by each age, for each gene/CNV–phenotype pair. We included only genotype-phenotype pairs where at least 20 individuals carrying a variant in the corresponding gene or CNV had been assessed for that phenotype, and where the overall penetrance exceeded 1%. Age-specific cumulative penetrance was estimated using the *cumulative_density* function from the *KaplanMeierFitter* module from the *lifelines* Python package (version 3.10.12). For each gene-phenotype combination, we also computed the cumulative penetrance curves derived from all other genes/CNVs in the dataset, providing a reference benchmark for cross-condition comparisons.

### Statistical Analyses

#### Fisher’s Exact Test for Phenotype Enrichment in Genetic NDDs

We performed genotype-phenotype enrichment analysis using Fisher’s exact test. For each genetic NDD, phenotype prevalence among its carriers was compared with prevalence among carriers of all other genetic NNDs in the dataset. We tested 168 phenotypes across 87 genetic NDDs; 1,295 genetic NDD/phenotype pairs had sufficient data to populate the 2×2 contingency table required for Fisher’s exact test. Therefore, the Bonferroni corrected significance threshold was set at alpha = 0.05/1,295 = 3.9×10⁻⁵. The test was performed through the *fisher_exact* function of the *scipy.stats* method of Python3.4.

#### Cox Proportional Hazards Analysis of Cumulative Penetrance in Genetic NDDs

To distinguish earlier versus later onset, we fit Cox proportional hazards models comparing each genetic NDD-phenotype trajectory with the pooled comparator (time scale: age at onset; event: first recorded manifestation). Only phenotypes showing an overall prevalence of at least 5% and assessed in at least 20 individuals per genetic NDD were included. A total of 464 tests were performed, with a Bonferroni-corrected significance threshold of alpha = 0.05/464 = 1×10⁻^4^. Analyses were implemented using the *CoxPHFitter* function of the *lifelines* package in Python3.4.

## Data Availability

All data produced are available online at https://nddportals.shinyapps.io/NDD-Portal/

## References

1. Litman A, Sauerwald N, Green Snyder L, et al. Decomposition of phenotypic heterogeneity in autism reveals underlying genetic programs. Nat Genet. 2025;57(7):1611–1619. doi:10.1038/s41588-025-02224-z

2. Zhang X, Grove J, Gu Y, et al. Polygenic and developmental profiles of autism differ by age at diagnosis. Nature. Published online October 1, 2025:1–12. doi:10.1038/s41586-025-09542-6

3. Bai D, Yip BHK, Windham GC, et al. Association of Genetic and Environmental Factors With Autism in a 5-Country Cohort. JAMA Psychiatry. 2019;76(10):1035–1043. doi:10.1001/jamapsychiatry.2019.1411

4. Sandin S, Lichtenstein P, Kuja-Halkola R, Hultman C, Larsson H, Reichenberg A. The Heritability of Autism Spectrum Disorder. JAMA. 2017;318(12):1182. doi:10.1001/jama.2017.12141

5. Stefanski A, Calle-López Y, Leu C, Pérez-Palma E, Pestana-Knight E, Lal D. Clinical sequencing yield in epilepsy, autism spectrum disorder, and intellectual disability: A systematic review and meta-analysis. Epilepsia. 2021;62(1):143–151. doi:10.1111/epi.16755

6. Eisfeldt J, Ek M, Nordenskjöld M, Lindstrand A. Toward clinical long-read genome sequencing for rare diseases. Nat Genet. 2025;57(6):1334–1343. doi:10.1038/s41588-025-02160-y

7. Del Gobbo GF, Boycott KM. The additional diagnostic yield of long-read sequencing in undiagnosed rare diseases. Genome Res. 2025;35(4):559–571. doi:10.1101/gr.279970.124

8. Zhou X, Feliciano P, Shu C, et al. Integrating de novo and inherited variants in 42,607 autism cases identifies mutations in new moderate-risk genes. Nat Genet. 2022;54(9):1305–1319. doi:10.1038/s41588-022-01148-2

9. Köhler S, Carmody L, Vasilevsky N, et al. Expansion of the Human Phenotype Ontology (HPO) knowledge base and resources. Nucleic Acids Res. 2019;47(D1):D1018–D1027. doi:10.1093/nar/gky1105

10. Antaki D, Guevara J, Maihofer AX, et al. A phenotypic spectrum of autism is attributable to the combined effects of rare variants, polygenic risk and sex. Nat Genet. 2022;54(9):1284–1292. doi:10.1038/s41588-022-01064-5

11. Darvish H, Azcona LJ, Tafakhori A, et al. Phenotypic and genotypic characterization of families with complex intellectual disability identified pathogenic genetic variations in known and novel disease genes. Sci Rep. 2020;10(1):968. doi:10.1038/s41598-020-57929-4

12. Kuchenbaecker KB, Hopper JL, Barnes DR, et al. Risks of Breast, Ovarian, and Contralateral Breast Cancer for BRCA1 and BRCA2 Mutation Carriers. JAMA. 2017;317(23):2402–2416. doi:10.1001/jama.2017.7112

13. Chakravarty D, Gao J, Phillips S, et al. OncoKB: A Precision Oncology Knowledge Base. JCO Precis Oncol. 2017;(1):1–16. doi:10.1200/PO.17.00011

14. Tate JG, Bamford S, Jubb HC, et al. COSMIC: the Catalogue Of Somatic Mutations In Cancer. Nucleic Acids Res. 2019;47(D1):D941–D947. doi:10.1093/nar/gky1015

15. Abrahams BS, Arking DE, Campbell DB, et al. SFARI Gene 2.0: a community-driven knowledgebase for the autism spectrum disorders (ASDs). Molecular Autism. 2013;4(1):36. doi:10.1186/2040-2392-4-36

16. Gidziela A, Ahmadzadeh YI, Michelini G, et al. A meta-analysis of genetic effects associated with neurodevelopmental disorders and co-occurring conditions. Nat Hum Behav. 2023;7(4):642–656. doi:10.1038/s41562-023-01530-y

17. Arnett AB, Wang T, Eichler EE, Bernier RA. Reflections on the genetics-first approach to advancements in molecular genetic and neurobiological research on neurodevelopmental disorders. Journal of Neurodevelopmental Disorders. 2021;13(1):24. doi:10.1186/s11689-021-09371-4

18. Bain JM, Snyder LG, Helbig KL, Cooper DD, Chung WK, Goodspeed K. Consistency of parent-report SLC6A1 data in Simons Searchlight with Provider-Based Publications. J Neurodev Disord. 2022;14:40. doi:10.1186/s11689-022-09449-7

19. Stevelink R, Piet M, Bos Y, Koeleman BPC. Penetrance of pathogenic epilepsy variants is low and shaped by common genetic background.

20. Garcia BT, Westerfield L, Yelemali P, et al. Improving automated deep phenotyping through large language models using retrieval-augmented generation. Genome Medicine. 2025;17(1):91. doi:10.1186/s13073-025-01521-w

21. Lin D. An Information-Theoretic Definition of Similarity. In: Proceedings of the Fifteenth International Conference on Machine Learning. ICML’98. Morgan Kaufmann Publishers Inc.; 1998:296–304.

