## Supplementary material for "Age-aware genotype–phenotype architecture across 87 genetic neurodevelopmental disorders"

**Article Type**: Original Investigation

**Corresponding Author**: Dennis Lal, PhD

Address: UTHealth, 1133 John Freeman Blvd, Houston, TX 77030-2809 (Texas, USA).

### Supplementary Material

**Supplementary Table 1** List of 75 genes and 12 CNVs included in the SFARI dataset 13.0.

| Gene/CNV | $N_{ind}$ | Age (years) | $N_{pheno}$ |
|---|---|---|---|
| **16p11.2 deletion** | **129** | **8.2 [6-14]** | **100** |
| **SCN2A** | **100** | **8.0 [4-12]** | **101** |
| **PPP2R5D** | **76** | **6.5 [4-11]** | **101** |
| **STXBP1** | **70** | **7.9 [3-11]** | **81** |
| **16p11.2 duplication** | **57** | **11.9 [6-21]** | **87** |
| **GRIN2B** | **53** | **8.1 [5-13]** | **76** |
| **SLC6A1** | **52** | **8.0 [3-12]** | **74** |
| **1q21.1 duplication** | **48** | **11.4 [6-34]** | **77** |
| **ASXL3** | **48** | **6.8 [4-13]** | **76** |
| **1q21.1 deletion** | **45** | **8.6 [4-16]** | **94** |
| **CTNNB1** | **39** | **4.6 [3-9]** | **70** |
| **SYNGAP1** | **36** | **8.8 [6-13]** | **63** |
| **CSNK2A1** | **31** | **8.5 [4-13]** | **79** |
| **DYRK1A** | **24** | **8.6 [4-16]** | **80** |
| **PACS1** | **24** | **7.6 [4-11]** | **82** |
| **CHAMP1** | **23** | **5.7 [2-10]** | **64** |
| **HNRNPH2** | **23** | **7.8 [4-16]** | **60** |
| **MED13L** | **21** | **6.1 [4-12]** | **58** |
| **SETBP1** | **20** | **7.8 [3-10]** | **53** |
| HIVEP2 | 19 | 8.4 [6-11] | 51 |
| ADNP | 18 | 9.3 [6-14] | 69 |
| DISTAL 16p11.2 del | 12 | 12.7 [6-21] | 52 |
| 7q11.23 dup | 9 | 10.4 [9-14] | 43 |
| ARID1B | 8 | 3.2 [2-5] | 37 |
| CHD2 | 8 | 9.1 [5-11] | 35 |
| CHD8 | 8 | 8.0 [6-9] | 36 |
| GRIN1 | 8 | 4.5 [2-13] | 38 |
| FOXP1 | 7 | 11.4 [10-14] | 47 |
| ANKRD11 | 6 | 11.5 [7-15] | 44 |
| DISTAL 16p11.2 dup | 6 | 12.0 [9-13] | 37 |
| SETD5 | 6 | 7.7 [6-9] | 44 |
| VPS13B | 6 | 4.5 [4-6] | 37 |
| PPP2R1A | 5 | 3.5 [3-17] | 51 |
| PTCHD1 | 5 | 6.2 [5-7] | 38 |
| TBR1 | 5 | 11.1 [8-16] | 45 |
| TRIP12 | 5 | 10.4 [4-12] | 24 |
| AUTS2 | 4 | 9.8 [7-15] | 29 |
| IRF2BPL | 4 | 16.6 [12-17] | 29 |
| 15q13.3 deletion | 3 | 14.1 [8-19] | 19 |
| 16p12.2 deletion | 3 | 16.8 [14-32] | 31 |
| 17q12 duplication | 3 | 15.4 [11-23] | 21 |
| 2p16.3_NRXN1 | 3 | 15.5 [7-30] | 14 |
| CTBP1 | 3 | 5.6 [4-8] | 25 |
| CUL3 | 3 | 12.3 [8-36] | 6 |
| DDX3X | 3 | 4.8 [4-6] | 17 |
| DNMT3A | 3 | 9.9 [6-25] | 25 |
| GIGYF1 | 3 | 13.8 [12-14] | 26 |
| KCNB1 | 3 | 8.9 [5-26] | 24 |
| KMT2C | 3 | 6.9 [6-8] | 11 |
| KMT2E | 3 | 6.7 [5-8] | 30 |
| TLK2 | 3 | 4.9 [3-6] | 23 |
| 15q11.2 BP1-BP2 del | 2 | 31.4 [22-40] | 11 |
| AHDC1 | 2 | 6.0 [4-7] | 18 |
| ASH1L | 2 | 6.8 [6-7] | 21 |
| CHD3 | 2 | 27.1 [27-27] | 19 |
| EHMT1 | 2 | 9.8 [7-12] | 11 |
| GRIN2A | 2 | 16.8 [13-19] | 22 |
| KDM6B | 2 | 7.8 [5-10] | 5 |
| MBD5 | 2 | 23.6 [18-28] | 26 |
| MED13 | 2 | 11.2 [9-13] | 19 |
| NR4A2 | 2 | 16.5 [12-20] | 11 |
| POGZ | 2 | 8.6 [7-10] | 34 |
| SCN1A | 2 | 30.2 [21-38] | 16 |
| SIN3A | 2 | 3.8 [3-4] | 25 |
| TRIO | 2 | 8.6 [7-9] | 12 |
| USP9X | 2 | 4.6 [3-5] | 20 |
| WAC | 2 | 6.6 [5-7] | 17 |
| 17q12 deletion | 1 | 9.8 [9-9] | 7 |
| ATRX | 1 | 17.5 [17-17] | 32 |
| BCL11A | 1 | 9.9 [9-9] | 13 |
| CAPRIN1 | 1 | 24.2 [24-24] | 20 |
| CNOT3 | 1 | 21.8 [21-21] | 13 |
| CSDE1 | 1 | 5.5 [5-5] | 8 |
| EIF3F | 1 | 12.7 [12-12] | 1 |
| HNRNPU | 1 | 13.2 [13-13] | 22 |
| IQSEC2 | 1 | 3.2 [3-3] | 9 |
| KMT2A | 1 | 17.2 [17-17] | 12 |
| KMT5B | 1 | 6.4 [6-6] | 8 |
| NAA15 | 1 | 21.9 [21-21] | 6 |
| NCKAP1 | 1 | 3.6 [3-3] | 24 |
| NEXMIF | 1 | 17.8 [17-17] | 7 |
| PHF21A | 1 | 9.2 [9-9] | 11 |
| SMARCC2 | 1 | 20.6 [20-20] | 4 |
| SOX5 | 1 | 1.6 [1-1] | 7 |
| SYNCRIP | 1 | 29.5 [29-29] | 20 |
| TANC2 | 1 | 15.9 [15-15] | 10 |
| ZNF292 | 1 | 5.1 [5-5] | 6 |

In the first column, *dup* indicates duplication and *del* indicates deletion. **$N_{ind}$**: total number of individuals carrying the CNV or a variant in the gene. **Age**: The median age in years with the $25^{th}$ and $75^{th}$ percentiles in square brakets. **$N_{pheno}$**: number of distinct phenotypes observed in at least one individual carrying the CNV or gene variant. In bold are shown genes/CNVs for which more than 20 individuals are present.

**Supplementary Table 2** List of the 168 neurodevelopmental phenotypes from the SFARI (Simons Foundation Autism Research Initiative) survey dataset with recorded age of onset.

| Phenotype | Question |
|---|---|
| Brain cancer | please specify Cancer type: brain |
| Cerebral palsy | Does the proband have cerebral palsy? |
| Spastic cerebral palsy | Does the proband have spastic cerebral palsy? |
| Clumsiness | Is the proband excessively clumsy or uncoordinated? |
| Constipation | Have there ever been symptoms of constipation (very hard stool)? |
| Diarrhea | Have there ever been symptoms of diarrhea (watery stool)? |
| Gastroesophageal reflux | Have there ever been symptoms of gastroesophageal reflux (GERD) (reverse travel of food or liquid significant enough to require medication)? |
| Gastrointestinal illness | Have there ever been symptoms of another gastrointestinal illness? |
| Cranial nerve disorder | Have there ever been symptoms of a cranial nerve disorder? |
| Encephalitis / Meningitis | Have there ever been symptoms of encephalitis / meningitis? |
| Febrile seizures | Have there ever been febrile seizures (with fever)? |
| Head injury/LOC | Has there ever been a head injury / loss of consciousness? |
| Hypertonia | Is the proband hypertonic (tight, spastic)? |
| Hypotonia | Is the proband hypotonic (low muscle tone, floppy)? |
| Macrocephaly | Have there ever been symptoms of macrocephaly? |
| Microcephaly | Have there ever been symptoms of microcephaly? |
| Pneumonia | Was pneumonia reported? |
| Positive lead screening | Has there ever been a positive lead screening? |
| Syringomyelia | Have there ever been symptoms of syringomyelia (fluid filled spinal cyst)? |
| Tourette’s/Tics | Have there ever been symptoms of Tourette’s or tics? |
| Urinary tract infection | Was a urinary tract infection reported? |
| Vision problem | Does the proband have a vision problem? |
| **Autoimmune conditions** | **Were Autoimmune conditions reported?** |
| Addison's disease | Please specify the Autoimmune type(s): Addison's disease |
| Alopecia areata | Please specify the Autoimmune type(s): alopecia areata |
| Chronic fatigue | Please specify the Autoimmune type(s): chronic fatigue |
| Fibromyalgia | Please specify the Autoimmune type(s): fibromyalgia |
| Fibromyositis | Please specify the Autoimmune type(s): fibromyositis |
| Guillain-Barre syndrome | Please specify the Autoimmune type(s): Guillain-Barré syndrome |
| Immune dysfunction syndrome | Please specify the Autoimmune type(s): immune dysfunction syndrome |
| Juvenile arthritis | Please specify the Autoimmune type(s): juvenile arthritis |
| Lupus | Please specify the Autoimmune type(s): lupus |
| Meniere’s disease | Please specify the Autoimmune type(s): Meniere’s disease |
| Multiple sclerosis | Please specify the Autoimmune type(s): multiple sclerosis |
| Mysthenia gravis | Please specify the Autoimmune type(s): myasthenia gravis |
| Pernicious anemia | Please specify the Autoimmune type(s): pernicious anemia |
| Psoriasis | Please specify the Autoimmune type(s): psoriasis |
| Raynaud phenomenon | Please specify the Autoimmune type(s): Raynaud phenomenon |
| Rheumatic fever | Please specify the Autoimmune type(s): rheumatic fever |
| Sarcoidosis | Please specify the Autoimmune type(s): sarcoidosis |
| Scleroderma | Please specify the Autoimmune type(s): scleroderma |
| Sjögren syndrome | Please specify the Autoimmune type(s): Sjögren syndrome |
| Vasculitis | Please specify the Autoimmune type(s): vasculitis |
| Vitiligo | Please specify the Autoimmune type(s): vitiligo |
| Other autoimmune | Please specify the Autoimmune type(s): other |
| **Bone abnormalities** | **Were Bone abnormalities reported?** |
| Craniosynostosis | Please specify the Bone abnormalities type(s): craniosynostosis |
| Osteoarthritis | Please specify the Bone abnormalities type(s): osteoarthritis |
| Osteoporosis | Please specify the Bone abnormalities type(s): osteoporosis |
| Pectus carinatum | Please specify the Bone abnormalities type(s): pectus carinatum |
| Pectus excavatum | Please specify the Bone abnormalities type(s): pectus excavatum |

| | |
|---|---|
| Polydactyly | Please specify the Bone abnormalities type(s): polydactyly |
| Rheumatoid arthritis | Please specify the Bone abnormalities type(s): rheumatoid arthritis |
| Rib anomalies | Please specify the Bone abnormalities type(s): rib anomalies |
| Scoliosis | Please specify the Bone abnormalities type(s): scoliosis |
| Vertebral anomalies | Please specify the Bone abnormalities type(s): vertebral anomalies |
| Other bone abnormality | Please specify the Bone abnormalities type(s): other |
| **Dental issues** | **Were dental issues reported?** |
| Early tooth eruption | Reported dental issue |
| Enamel defects | Reported dental issue |
| Late tooth eruption | Reported dental issue |
| Misshapen teeth | Reported dental issue |
| Missing teeth | Reported dental issue |
| Roots long/shallow | Reported dental issue |
| Too many teeth | Reported dental issue |
| Other dental issues | Reported dental issue |
| **Endocrinologic abnormalities** | **Were Endocrinologic abnormalities reported?** |
| Diabetes | Please specify the Endocrinologic abnormalities type(s) diabetes |
| Failure to thrive | Please specify the Endocrinologic abnormalities type(s) failure to thrive |
| Hypothyroidism | Please specify the Endocrinologic abnormalities type(s) hypothyroidism |
| Hyperthyroidism | Please specify the Endocrinologic abnormalities type(s) hyperthyroidism |
| Irregular menses | Please specify the Endocrinologic abnormalities type(s) irregular menses |
| Obesity | Please specify the Endocrinologic abnormalities type(s) obesity |
| Precocious puberty | Please specify the Endocrinologic abnormalities type(s) precocious puberty |
| Short stature | Please specify the Endocrinologic abnormalities type(s) short stature |
| Other endocrinologic abnormality | Please specify the Endocrinologic abnormalities type(s) other |
| **Eye condition** | **Does the child have any eye conditions?** |
| Amblyopia | Please specify the existing condition(s): amblyopia |
| Anatomical blindness | Please specify the existing condition(s): Anatomical blindness |
| Astigmatism | Please specify the existing condition(s): astigmatism |
| Cataract | Please specify the existing condition(s): cataract |
| Choroidal hemangioma | Please specify the existing condition(s): choroidal hemangioma |
| Coloboma | Please specify the existing condition(s): coloboma |
| Color blindness | Please specify the existing condition(s): color blindness |
| Conjunctival abnormality | Please specify the existing condition(s): conjunctival abnormality |
| Cortical blindness | Please specify the existing condition(s): cortical blindness |
| Depth perception problem | Please specify the existing condition(s): depth perception problem |
| Eye movement abnormalities | Please specify the existing condition(s): eye movement abnormalities |
| Glaucoma | Please specify the existing condition(s): glaucoma |
| Farsighted | Please specify the existing condition(s): farsighted |
| Nearsighted | Please specify the existing condition(s): nearsighted |
| Nystagmus | Please specify the existing condition(s): nystagmus |
| Optic nerve change | Please specify the existing condition(s): optic nerve change |
| Ptosis | Please specify the existing condition(s): ptosis |
| Retinal detachment | Please specify the existing condition(s): retinal detachment |
| Strabismus | Please specify the existing condition(s): strabismus |
| Synophrys | Please specify the existing condition(s): synophrys |
| Visual field impairment | Please specify the existing condition(s): visual field impairment |
| Other eye condition | Please specify the existing condition(s): other |
| **Genital problems** | **Were Genital problems reported?** |
| Ambiguous genitalia | Please specify the genital problem type(s): ambiguous genitalia |
| Bicornate uterus | Please specify the genital problem type(s): bicornate uterus |
| Chordae | Please specify the genital problem type(s): chordae |
| Hypospadias | Please specify the genital problem type(s): hypospadias |
| Infertility | Please specify the genital problem type(s): infertility |
| Undescended testicles | Please specify the genital problem type(s): undescended testicles |
| Other genital problems | Please specify the genital problem type(s): other |
| **Heart problems** | **Were heart problems reported?** |
| Arrhythmias | Please specify the heart problem type(s): arrhythmias |
| Atrial septal defect | What type of congenital heart problem was reported? atrial septal defect |

| | |
|---|---|
| Atrioventricular canal | What type of congenital heart problem was reported? atrioventricular canal |
| Bicuspid aortic valve | What type of congenital heart problem was reported? bicuspid aortic valve |
| Cardiomyopathy | Please specify the heart problem type(s): cardiomyopathy |
| Coarctation of the aorta | What type of congenital heart problem was reported? coarctation of the aorta |
| Hypoplastic right heart | What type of congenital heart problem was reported? hypoplastic right heart |
| Hypoplastic left heart | What type of congenital heart problem was reported? hypoplastic left |
| Interrupted aortic arch | What type of congenital heart problem was reported? interrupted aortic arch |
| Partial anomalous pulmonary venous return | What type of congenital heart problem was reported? partial anomalous pulmonary venous return |
| Patent ductus | What type of congenital heart problem was reported? patent ductus |
| Pulmonic stenosis | What type of congenital heart problem was reported? pulmonic stenosis |
| Shones complex | What type of congenital heart problem was reported? Shone's complex |
| Tetralogy of Fallot | What type of congenital heart problem was reported? tetralogy of Fallot |
| Total anomalous pulmonary venous return | What type of congenital heart problem was reported? total anomalous pulmonary venous return |
| Transposition of the great arteries | What type of congenital heart problem was reported? transposition of the great arteries |
| Ventricular septal defect | What type of congenital heart problem was reported? ventricular septal defect |
| Other heart problem | Please specify the heart problem type(s): other |
| **Immunodeficiency** | **Was immunodeficiency reported?** |
| Common immunodeficiency | Reported immunodeficiency |
| Hyper IgA Syndrome | Reported immunodeficiency |
| Mast cell immunodeficiency | Reported immunodeficiency |
| Unspecified immunodeficiency | Reported immunodeficiency |
| Other immunodeficiency | Reported immunodeficiency |
| **Kidney/Urinary problems** | **Were Kidney or Urinary problems reported?** |
| Hydronephrosis | Please specify the kidney or urinary problem type(s): hydronephrosis |
| Nephritis | Please specify the kidney or urinary problem type(s): nephritis |
| Nephrotic syndrome | Please specify the kidney or urinary problem type(s): nephrotic syndrome |
| Posterior urethral valves | Please specify the kidney or urinary problem type(s): posterior urethral valves |
| Renal agenesis | Please specify the kidney or urinary problem type(s): renal agenesis |
| Urinary reflux | Please specify the kidney or urinary problem type(s): urinary reflux |
| Other kidney/urinary problem | Please specify the kidney or urinary problem type(s): other |
| **Movement abnormality** | **Have there ever been any movement abnormalities? (e.g. tremor, ataxia, dystonic movements)** |
| Ataxia | Reported movement abnormality: Ataxia |
| Chorea | Reported movement abnormality: Chorea |
| Choreathetosis | Reported movement abnormality: Choreathetosis |
| Dystonia | Reported movement abnormality: Dystonia |
| Myoclonus | Reported movement abnormality: Myoclonus |
| Tremors | Reported movement abnormality: Tremors |
| Unspecified movement abnormality | Reported movement abnormality: Unspecified |
| Other movement abnormality | Reported movement abnormality: Other |
| **Respiratory problems** | **Were respiratory problems reported?** |
| Recurrent pneumonia | Please specify the respiratory problem type(s): Recurrent pneumonia |
| Other respiratory problem | Please specify the respiratory problem type(s): Other respiratory problem |
| **Seizures** | Have there ever been seizures? |
| Absence seizures | Is there a presence of petit mal or absence seizures (staring spell)? |
| Atonic seizures | Is there a presence of atonic or drop attack seizures (sudden loss of muscle strength)? |
| Epileptic spasms | Is there a presence of infantile spasm seizures (sudden jerk followed by stiffening)? |
| Focal aware seizures | Is there a presence of simple partial - focal seizures (consciousness unaltered)? |
| Focal impaired-awareness seizures | Is there a presence of complex partial seizures (altered consciousness)? |
| Generalized tonic-clonic seizures | Is there a presence of grand mal or generalized tonic clonic seizures (loss of consciousness and violent convulsions)? |
| Myoclonic seizures | Is there a presence of myoclonic seizures? |
| Tonic seizures | Is there a presence of tonic seizures? |
| **Sleep issues** | **Were sleep issues reported?** |
| Daytime sleepiness | Reported sleep issue |
| Issues falling asleep | Reported sleep issue |
| Issues staying asleep | Reported sleep issue |

| | |
|---|---|
| Sleep Apnea | Reported sleep issue |
| Sleep problems | Reported sleep issue |
| Sleeping too much | Reported sleep issue |
| Sleep (unspecified) | Reported sleep issue |

**Supplementary Table 3** List of SFARI phenotypes with corresponding top-level HPO categorization.

| HPO category | SFARI Phenotypes |
|---|---|
| HP:0000707 Abnormality of the nervous system | Brain cancer<br>Cerebral palsy<br>Spastic cerebral palsy<br>Clumsiness<br>Cranial nerve disorder<br>Encephalitis / Meningitis<br>Febrile seizures<br>Microcephaly<br>Syringomyelia<br>Tourettes/Tics<br>Guillain-Barre syndrome<br>Movement abnormality<br>Ataxia<br>Chorea<br>Choreathetosis<br>Dystonia<br>Myoclonus<br>Tremors<br>Unspecified movement abnormality<br>Other movement abnormality<br>Seizures<br>Absence seizures<br>Atonic seizures<br>Epileptic spasms<br>Focal aware seizures<br>Focal impaired-awareness seizures<br>Generalized tonic-clonic seizures<br>Myoclonic seizures<br>Tonic seizures<br>Sleep issues<br>Daytime sleepiness<br>Issues falling asleep<br>Issues staying asleep<br>Sleep Apnea<br>Sleep problems |

| | |
|---|---|
| | Sleeping too much<br>Sleep (unspecified) |
| HP:0025031<br>Abnormality of the digestive system | Constipation<br>Diarrhea<br>Gastroesophageal reflux<br>Gastrointestinal illness |
| HP:0033127<br>Abnormality of the musculoskeletal system | Hypertonia<br>Hypotonia<br>Fibromyositis<br>Juvenile arthritis<br>Mysthenia gravis<br>Bone abnormalities<br>Osteoarthritis<br>Osteoporosis<br>Pectus carinatum<br>Pectus excavatum<br>Polydactyly<br>Rheumatoid arthritis<br>Rib anomalies<br>Scoliosis<br>Vertebral anomalies<br>Other bone abnormality |
| HP:0000152<br>Abnormality of head or neck | Macrocephaly<br>Craniosynostosis<br>Dental issues<br>Early eruption<br>Enamel defects<br>Late eruption<br>Misshapen teeth<br>Missing teeth<br>Roots long/shallow<br>Too many teeth<br>Other dental issues<br>Conjunctival abnormality |
| HP:0002086<br>Abnormality of the respiratory system | Pneumonia<br>Partial anomalous pulmonary venous return<br>Total anomalous pulmonary venous return<br>Respiratory problems<br>Recurrent pneumonia<br>Other respiratory problem |
| HP:0002715<br>Abnormality of the immune system | Urinary tract infection<br>Autoimmune conditions<br>Immune dysfunction syndrome<br>Lupus<br>Other autoimmune<br>Immunodeficiency<br>Common immunodeficiency<br>Hyper IgA Syndrome<br>Mast cell immunodeficiency<br>Unspecified immunodeficiency<br>Other immunodeficiency |
| HP:0000478<br>Abnormality of the eye | Vision problem<br>Eye condition |

| | |
|---|---|
| | Amblyopia<br>Anatomical blindness<br>Astigmatism<br>Coloboma<br>Color blindness<br>Cortical blindness<br>Depth perception problem<br>Eye movement abnormalities<br>Glaucoma Farsighted<br>Nearsighted<br>Nystagmus<br>Optic nerve change<br>Ptosis Retinal detachment<br>Strabismus<br>Visual field impairment<br>Other eye condition |
| HP:0000818<br>Abnormality of the endocrine system | Addison's disease<br>Endocrinologic abnormalities<br>Diabetes<br>Hypothyroidism<br>Hyperthyroidism<br>Precocious puberty<br>Other endocrinologic abnormality |
| HP:0001574<br>Abnormality of the integument | Alopecia areata<br>Psoriasis<br>Scleroderma<br>Vitiligo<br>Synophrys |
| HP:0025142<br>Constitutional symptom | Chronic fatigue<br>Fibromyalgia |
| HP:0001871<br>Abnormality of blood and blood-forming tissues | Pernicious anemia |
| HP:0001626<br>Abnormality of the cardiovascular system | Raynaud phenomenon<br>Vasculitis<br>Choroidal hemangioma<br>Heart problems<br>Arrhythmias<br>Atrial septal defect<br>Atrioventricular canal<br>Bicuspid aortic valve<br>Cardiomyopathy<br>Coarctation of the aorta<br>Hypoplastic right heart<br>Hypoplastic left heart<br>Interrupted aortic arch<br>Patent ductus<br>Pulmonic stenosis<br>Tetralogy of fallot<br>Transposition of the great arteries<br>Ventricular septal defect<br>Other heart problem |
| HP:0001507 | Failure to thrive |

| | |
|---|---|
| Growth abnormality | Obesity<br>Short stature |
| HP:0000119<br>Abnormality of the genitourinary system | Irregular menses<br>Genital problems<br>Ambiguous genitalia<br>Bicornate uterus<br>Chordae<br>Hypospadias<br>Infertility<br>Undescended testicles<br>Other genital problems<br>Kidney/Urinary problems<br>Hydronephrosis<br>Nephritis<br>Nephrotic syndrome<br>Posterior urethral valves<br>Renal agenesis<br>Urinary reflux<br>Other kidney/urinary problem |

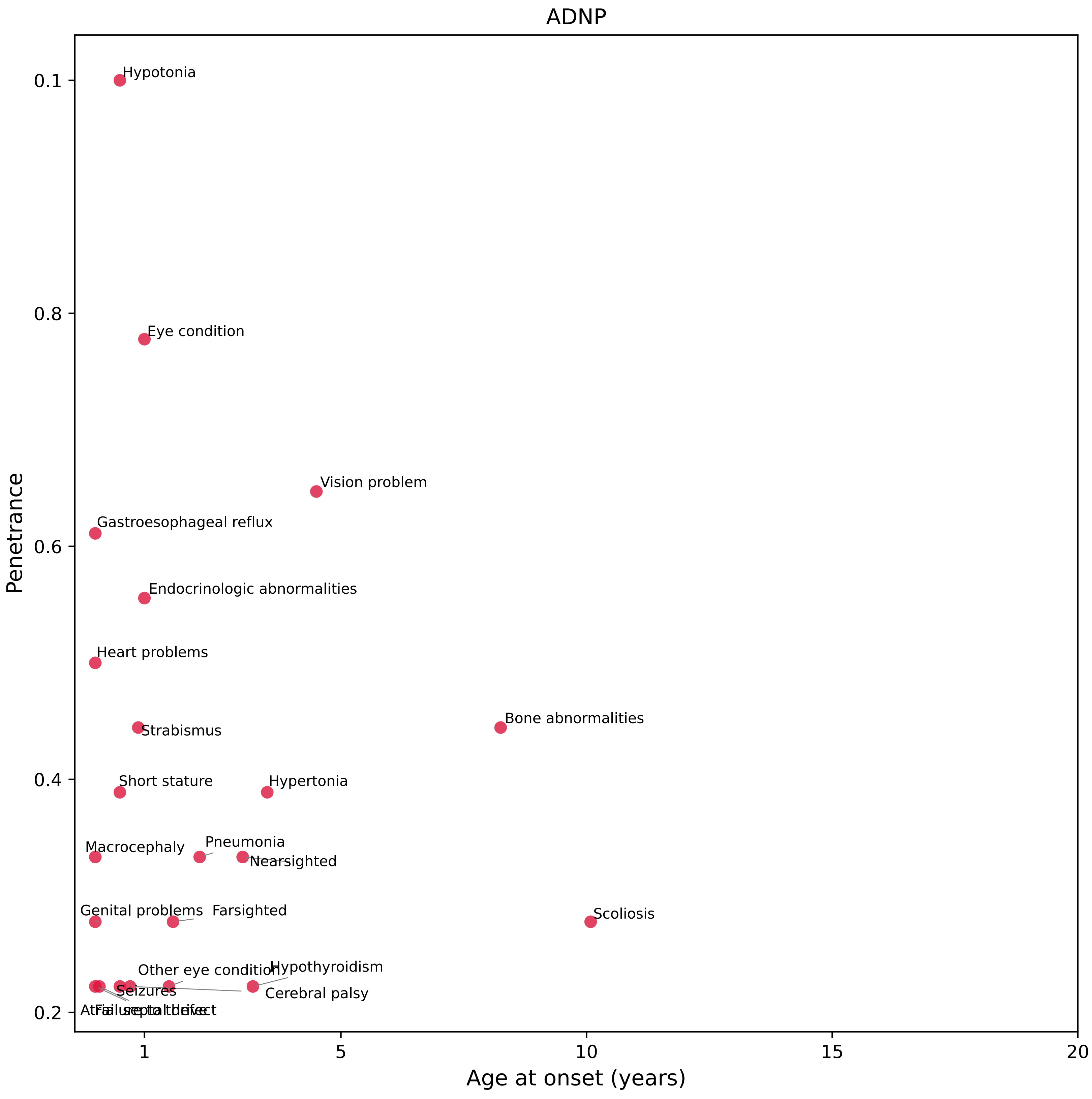
ADNP
Penetrance
Age at onset (years)
0.1
0.8
0.6
0.4
0.2
1
5
10
15
20
Hypotonia
Eye condition
Vision problem
Gastroesophageal reflux
Endocrinologic abnormalities
Heart problems
Strabismus
Bone abnormalities
Short stature
Hypertonia
Macrocephaly
Pneumonia
Nearsighted
Genital problems
Farsighted
Scoliosis
Other eye condition
Hypothyroidism
Seizures
Cerebral palsy

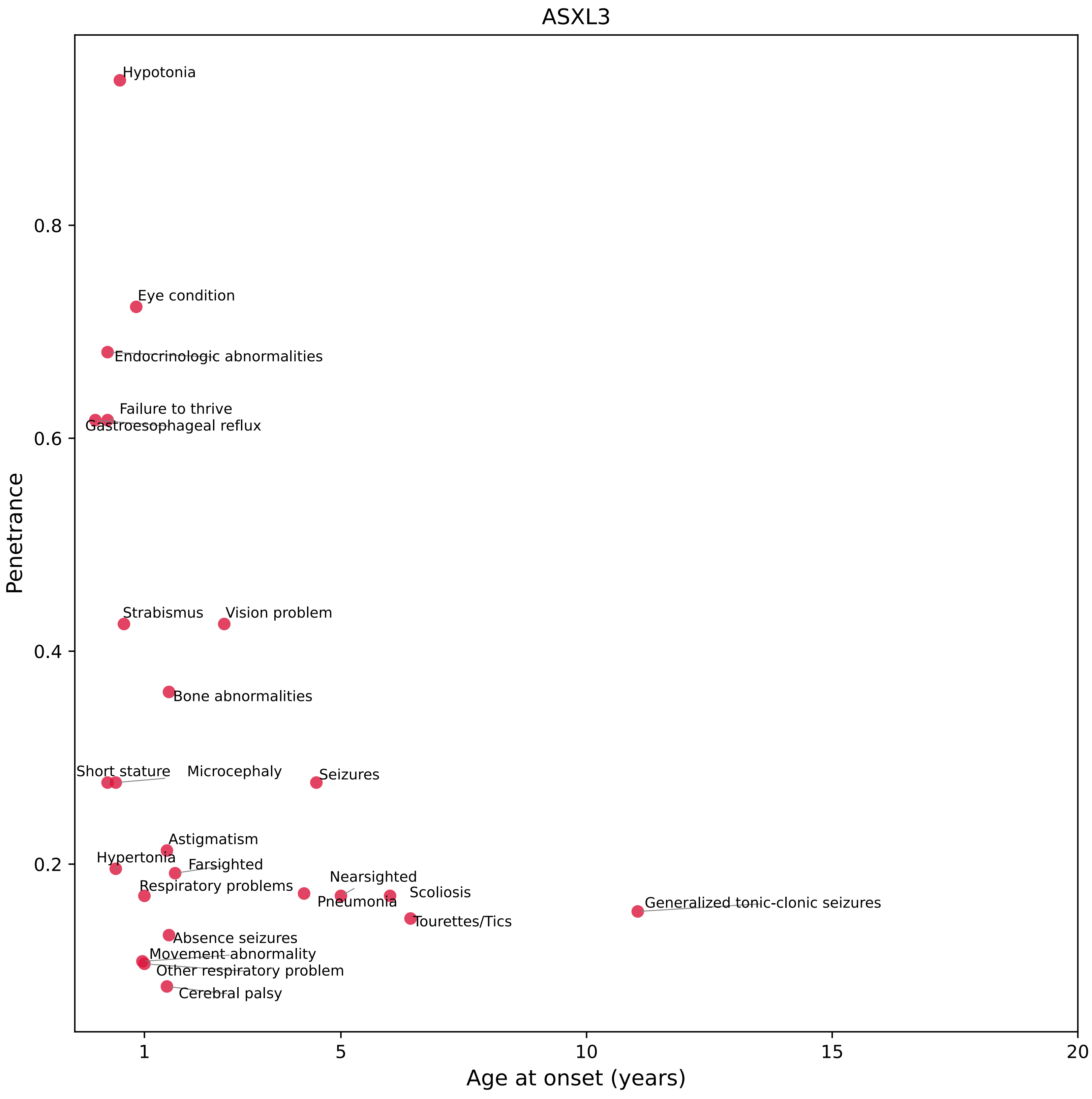

ASXL3
Hypotonia
Eye condition
Endocrinologic abnormalities
Failure to thrive
Gastroesophageal reflux
Strabismus
Vision problem
Bone abnormalities
Short stature
Microcephaly
Seizures
Astigmatism
Hypertonia
Farsighted
Nearsighted
Respiratory problems
Scoliosis
Pneumonia
Generalized tonic-clonic seizures
Tourettes/Tics
Absence seizures
Movement abnormality
Other respiratory problem
Cerebral palsy
Penetrance
0.8
0.6
0.4
0.2
1
5
10
15
20
Age at onset (years)

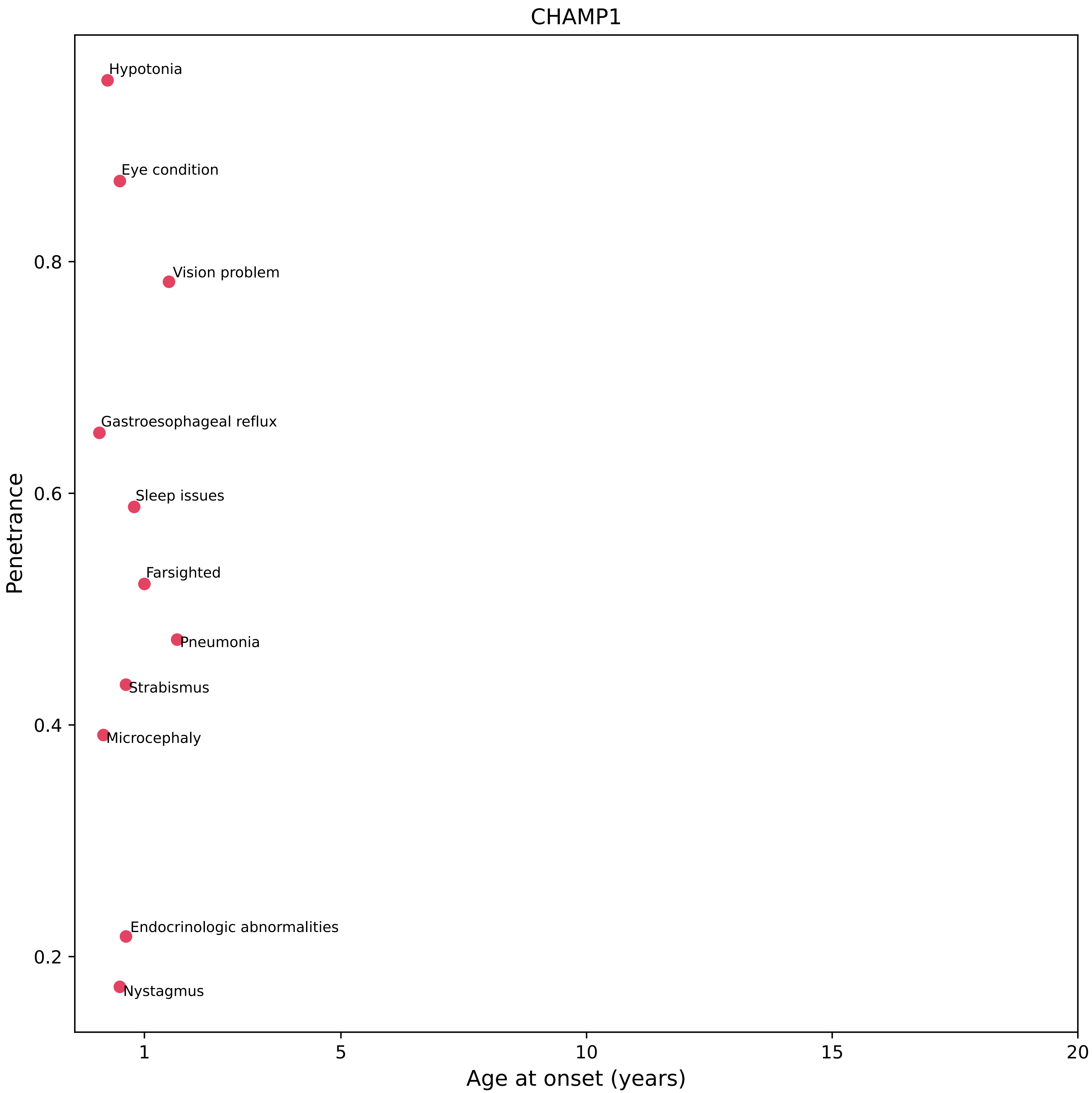
CHAMP1
Hypotonia
Eye condition
Vision problem
Gastroesophageal reflux
Sleep issues
Farsighted
Pneumonia
Strabismus
Microcephaly
Endocrinologic abnormalities
Nystagmus
Penetrance
0.8
0.6
0.4
0.2
1
5
10
15
20
Age at onset (years)

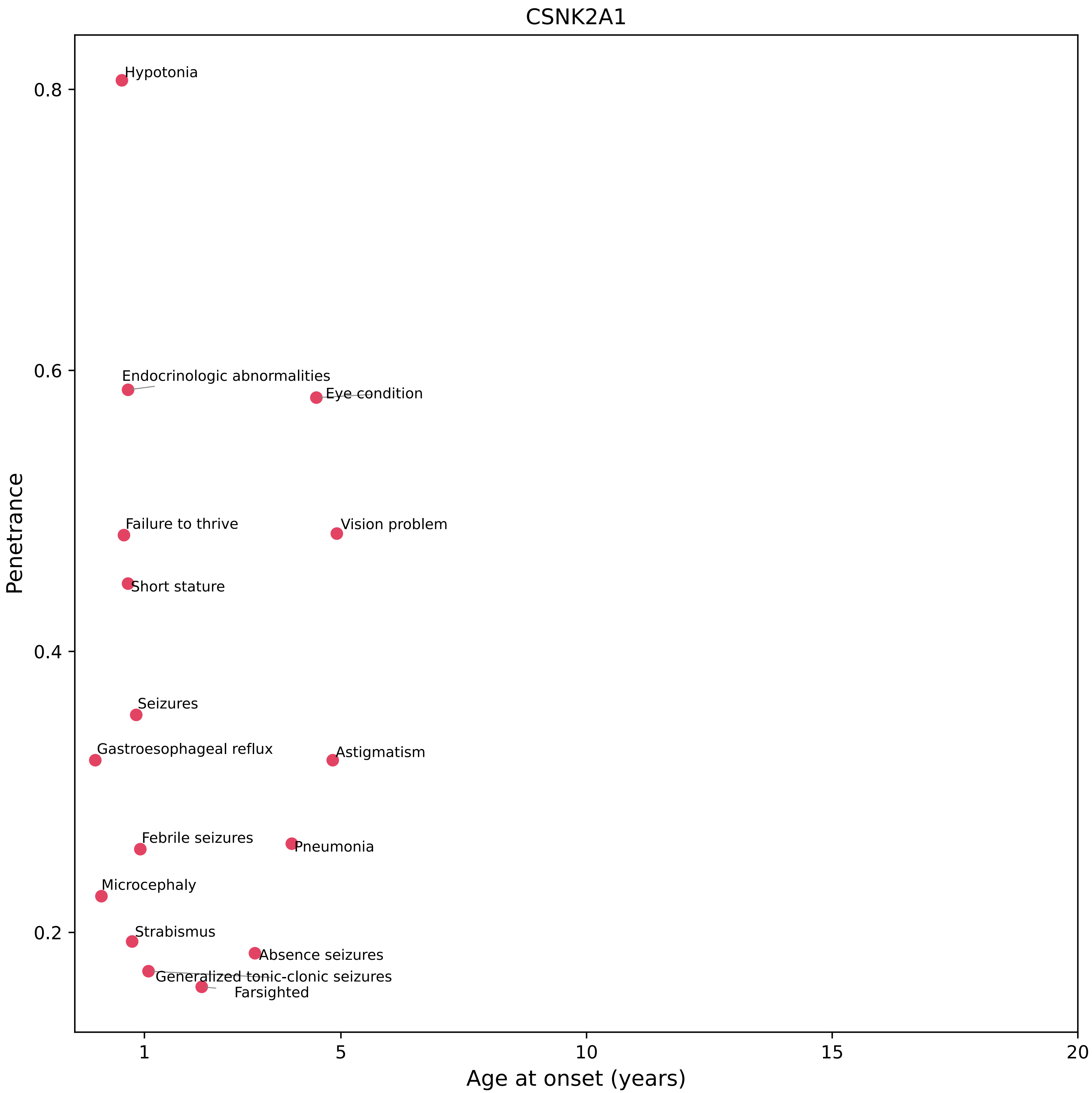

CSNK2A1
Hypotonia
Endocrinologic abnormalities
Eye condition
Failure to thrive
Vision problem
Short stature
Seizures
Gastroesophageal reflux
Astigmatism
Febrile seizures
Pneumonia
Microcephaly
Strabismus
Absence seizures
Generalized tonic-clonic seizures
Farsighted
0.8
0.6
0.4
0.2
Penetrance
1
5
10
15
20
Age at onset (years)

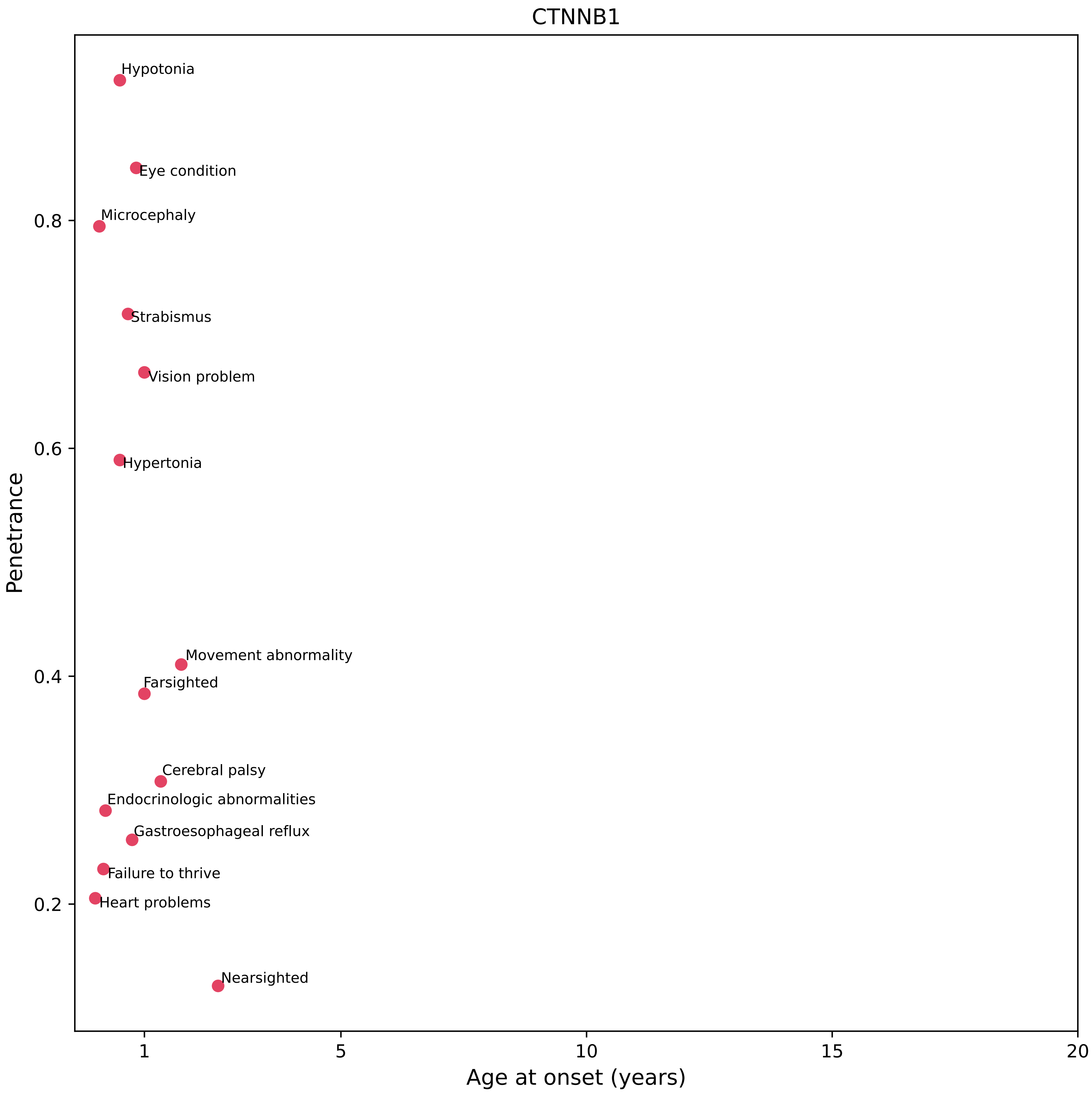
CTNNB1
Hypotonia
Eye condition
Microcephaly
Strabismus
Vision problem
Hypertonia
Movement abnormality
Farsighted
Cerebral palsy
Endocrinologic abnormalities
Gastroesophageal reflux
Failure to thrive
Heart problems
Nearsighted
Penetrance
0.8
0.6
0.4
0.2
1
5
10
15
20
Age at onset (years)

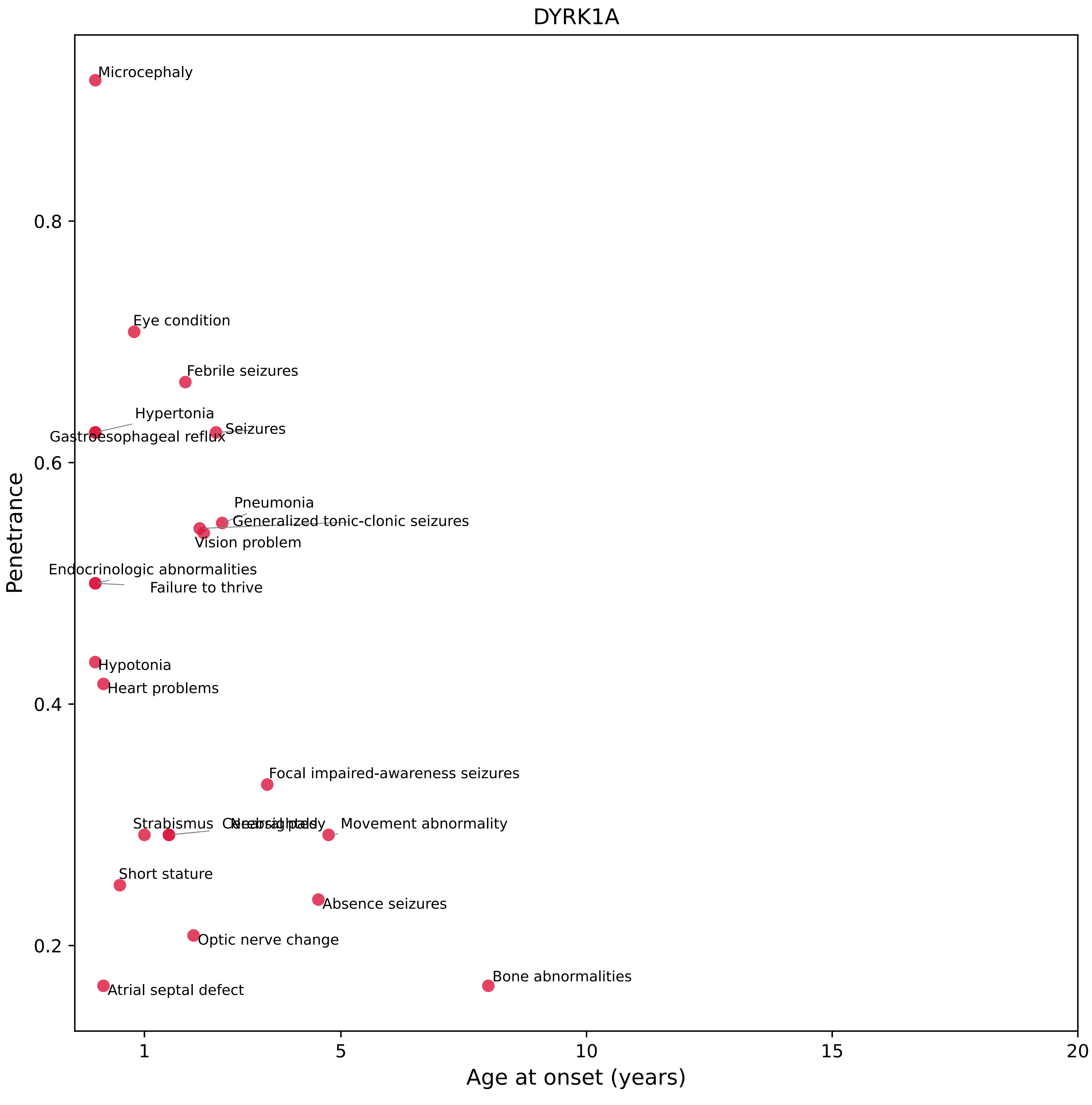
DYRK1A
Microcephaly
Eye condition
Febrile seizures
Hypertonia
Gastroesophageal reflux
Seizures
Pneumonia
Generalized tonic-clonic seizures
Vision problem
Endocrinologic abnormalities
Failure to thrive
Hypotonia
Heart problems
Focal impaired-awareness seizures
Strabismus
Movement abnormality
Short stature
Absence seizures
Optic nerve change
Atrial septal defect
Bone abnormalities
0.8
0.6
0.4
0.2
Penetrance
1
5
10
15
20
Age at onset (years)

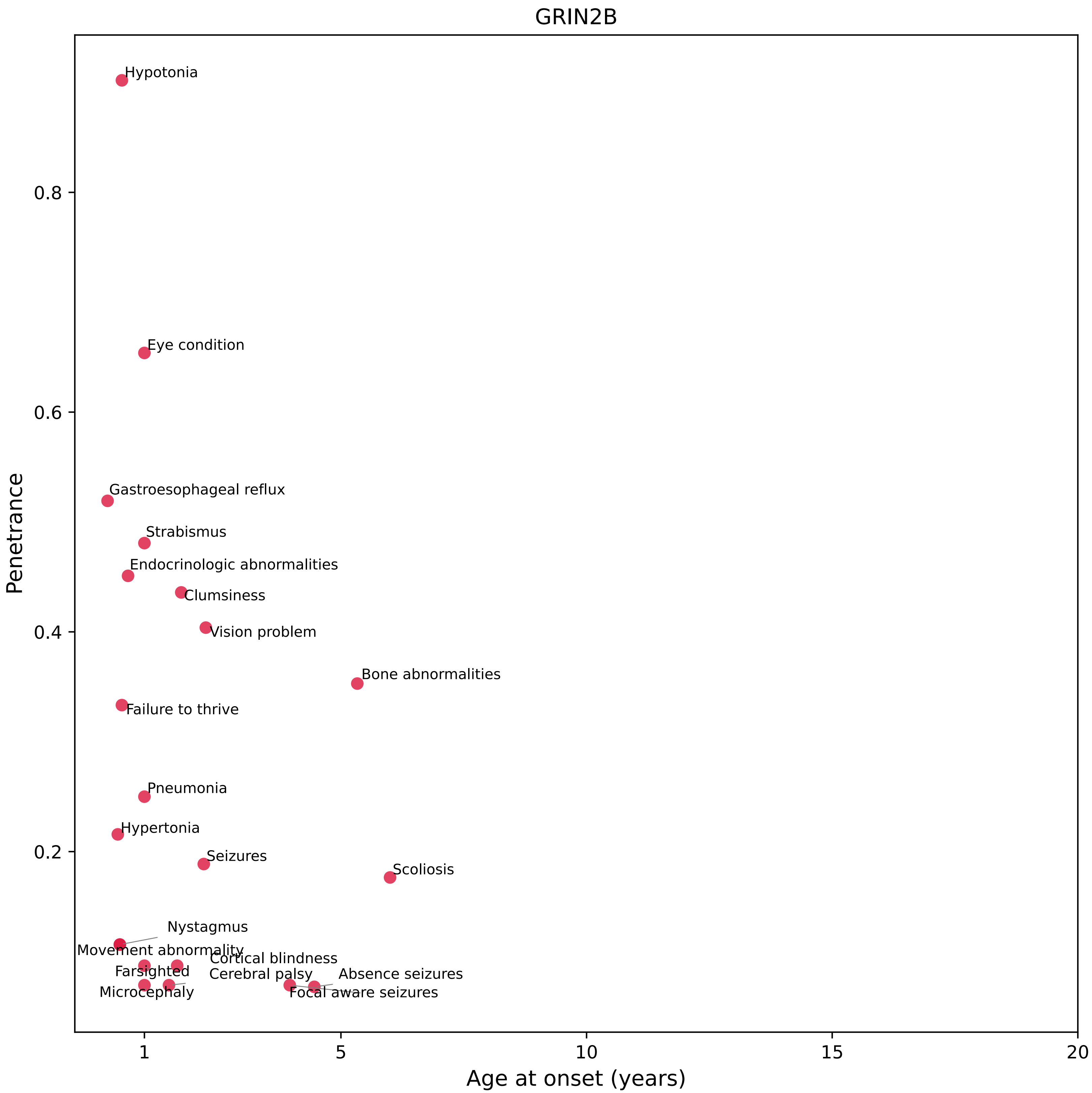

GRIN2B
Hypotonia
Eye condition
Gastroesophageal reflux
Strabismus
Endocrinologic abnormalities
Clumsiness
Vision problem
Bone abnormalities
Failure to thrive
Pneumonia
Hypertonia
Seizures
Scoliosis
Nystagmus
Movement abnormality
Cortical blindness
Farsighted
Cerebral palsy
Absence seizures
Microcephaly
Focal aware seizures
Penetrance
0.8
0.6
0.4
0.2
1
5
10
15
20
Age at onset (years)

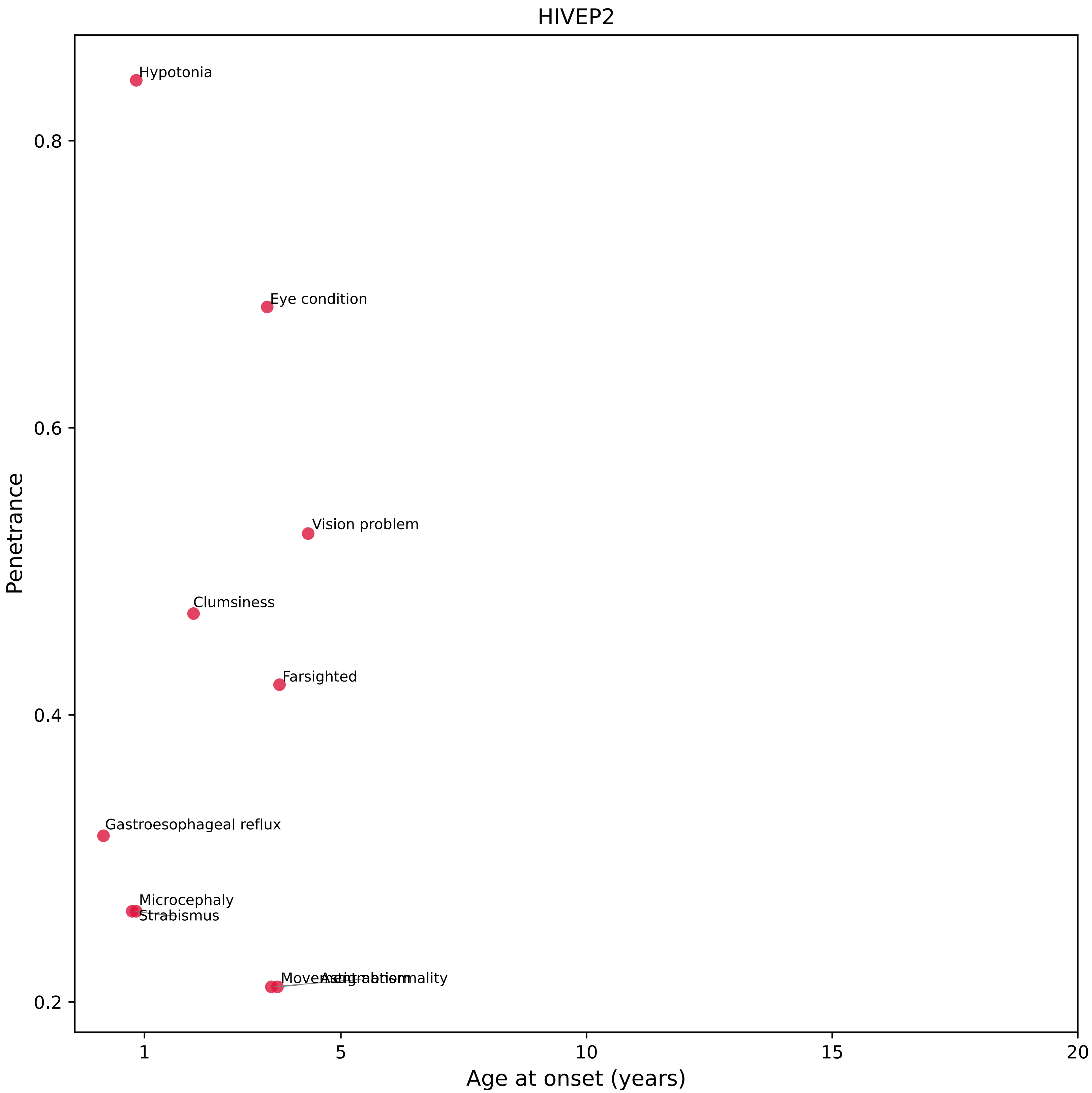
HIVEP2
Hypotonia
Eye condition
Vision problem
Clumsiness
Farsighted
Gastroesophageal reflux
Microcephaly
Strabismus
Movement abnormality
Astigmatism
0.8
0.6
0.4
0.2
Penetrance
1
5
10
15
20
Age at onset (years)

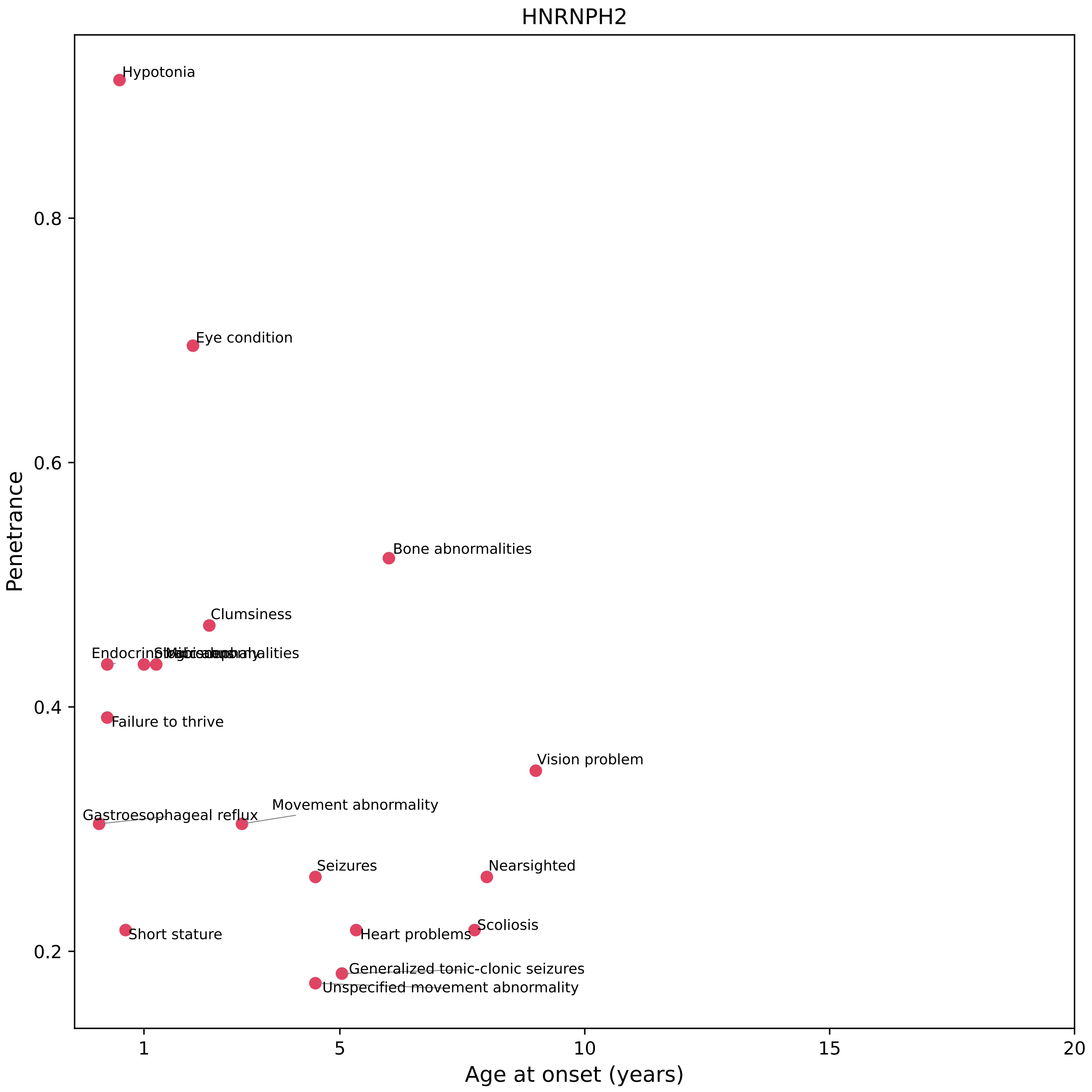
HNRNPH2
Hypotonia
Eye condition
Bone abnormalities
Clumsiness
Failure to thrive
Vision problem
Gastroesophageal reflux
Movement abnormality
Seizures
Nearsighted
Scoliosis
Short stature
Heart problems
Generalized tonic-clonic seizures
Unspecified movement abnormality
0.8
0.6
0.4
0.2
Penetrance
1
5
10
15
20
Age at onset (years)

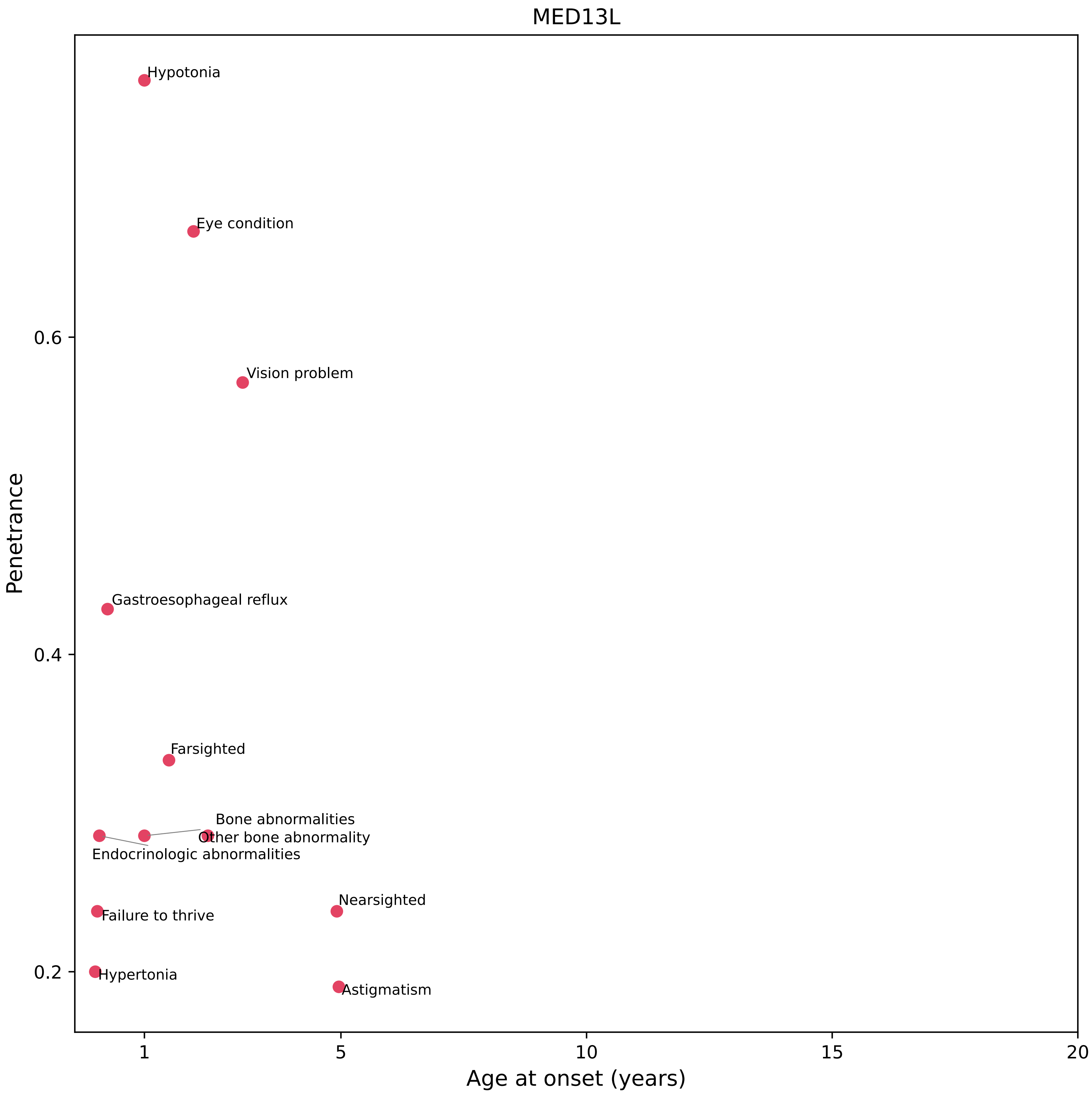
MED13L
Hypotonia
Eye condition
Vision problem
Gastroesophageal reflux
Farsighted
Bone abnormalities
Other bone abnormality
Endocrinologic abnormalities
Failure to thrive
Nearsighted
Hypertonia
Astigmatism
Penetrance
0.6
0.4
0.2
1
5
10
15
20
Age at onset (years)

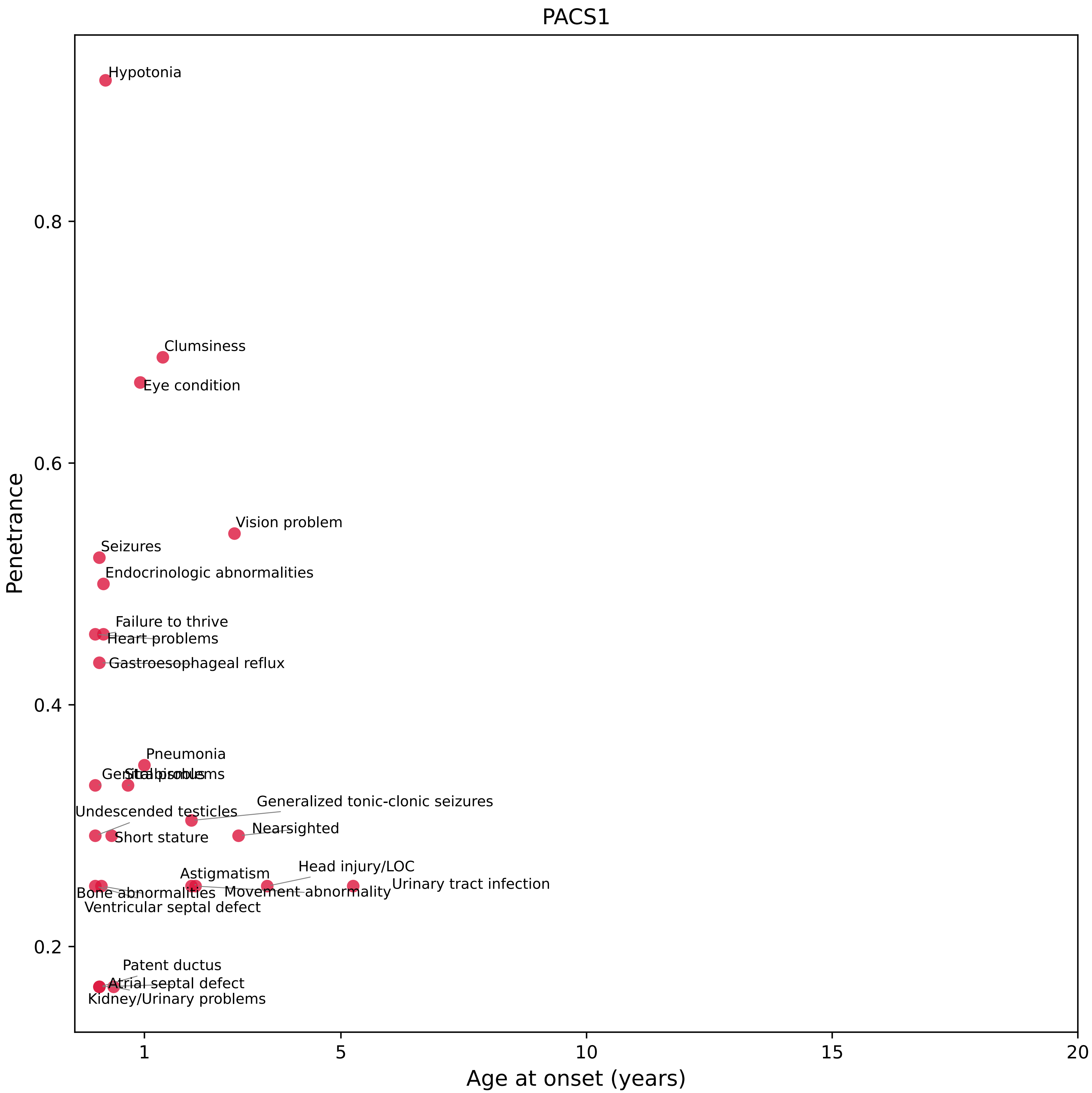
PACS1
Penetrance
Age at onset (years)
0.2
0.4
0.6
0.8
1
5
10
15
20
Hypotonia
Clumsiness
Eye condition
Vision problem
Seizures
Endocrinologic abnormalities
Failure to thrive
Heart problems
Gastroesophageal reflux
Pneumonia
Strabismus
Generalized tonic-clonic seizures
Undescended testicles
Nearsighted
Short stature
Head injury/LOC
Astigmatism
Urinary tract infection
Movement abnormality
Bone abnormalities
Ventricular septal defect
Patent ductus
Atrial septal defect
Kidney/Urinary problems

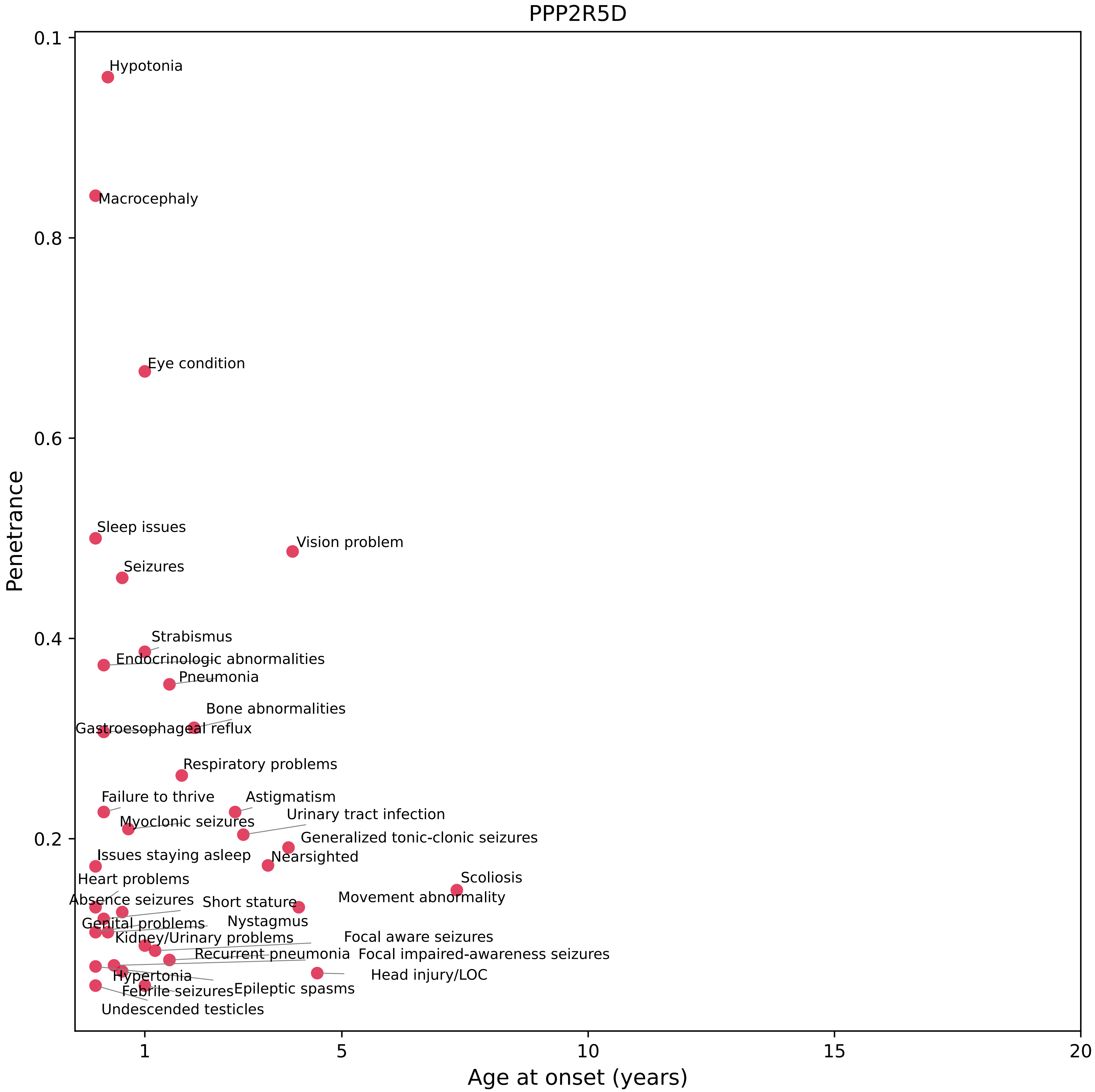
PPP2R5D
Penetrance
Age at onset (years)
0.1
0.8
0.6
0.4
0.2
1
5
10
15
20
Hypotonia
Macrocephaly
Eye condition
Sleep issues
Vision problem
Seizures
Strabismus
Endocrinologic abnormalities
Pneumonia
Bone abnormalities
Gastroesophageal reflux
Respiratory problems
Failure to thrive
Astigmatism
Urinary tract infection
Myoclonic seizures
Generalized tonic-clonic seizures
Issues staying asleep
Nearsighted
Scoliosis
Heart problems
Movement abnormality
Absence seizures
Short stature
Nystagmus
Genital problems
Kidney/Urinary problems
Focal aware seizures
Recurrent pneumonia
Focal impaired-awareness seizures
Hypertonia
Head injury/LOC
Febrile seizures
Epileptic spasms
Undescended testicles

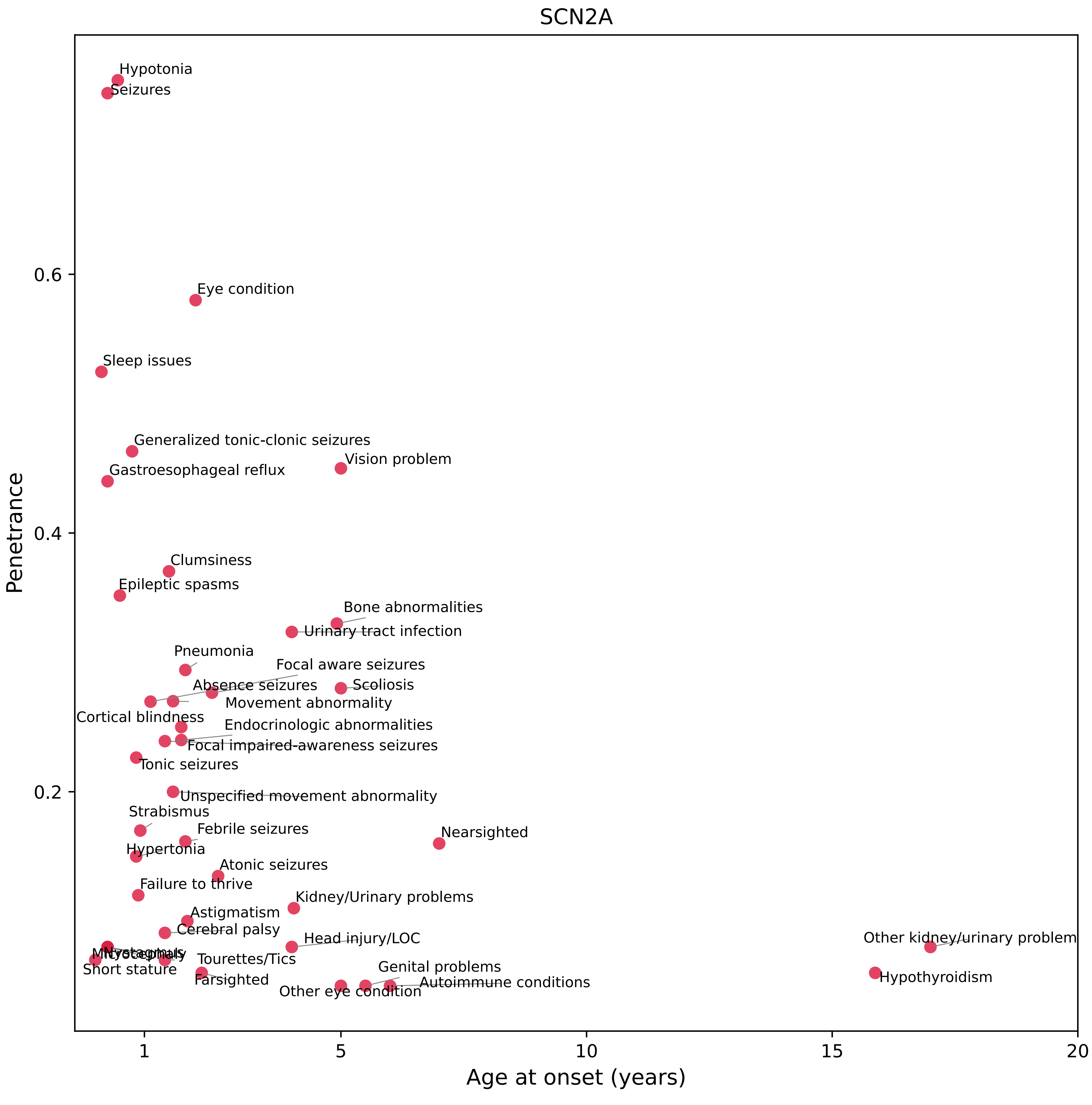
SCN2A
Penetrance
Age at onset (years)
0.2
0.4
0.6
1
5
10
15
20
Hypotonia
Seizures
Eye condition
Sleep issues
Generalized tonic-clonic seizures
Vision problem
Gastroesophageal reflux
Clumsiness
Epileptic spasms
Bone abnormalities
Urinary tract infection
Pneumonia
Focal aware seizures
Absence seizures
Scoliosis
Movement abnormality
Cortical blindness
Endocrinologic abnormalities
Focal impaired-awareness seizures
Tonic seizures
Unspecified movement abnormality
Strabismus
Febrile seizures
Nearsighted
Hypertonia
Atonic seizures
Failure to thrive
Kidney/Urinary problems
Astigmatism
Cerebral palsy
Head injury/LOC
Other kidney/urinary problem
Nystagmus
Tourettes/Tics
Short stature
Farsighted
Genital problems
Autoimmune conditions
Other eye condition
Hypothyroidism

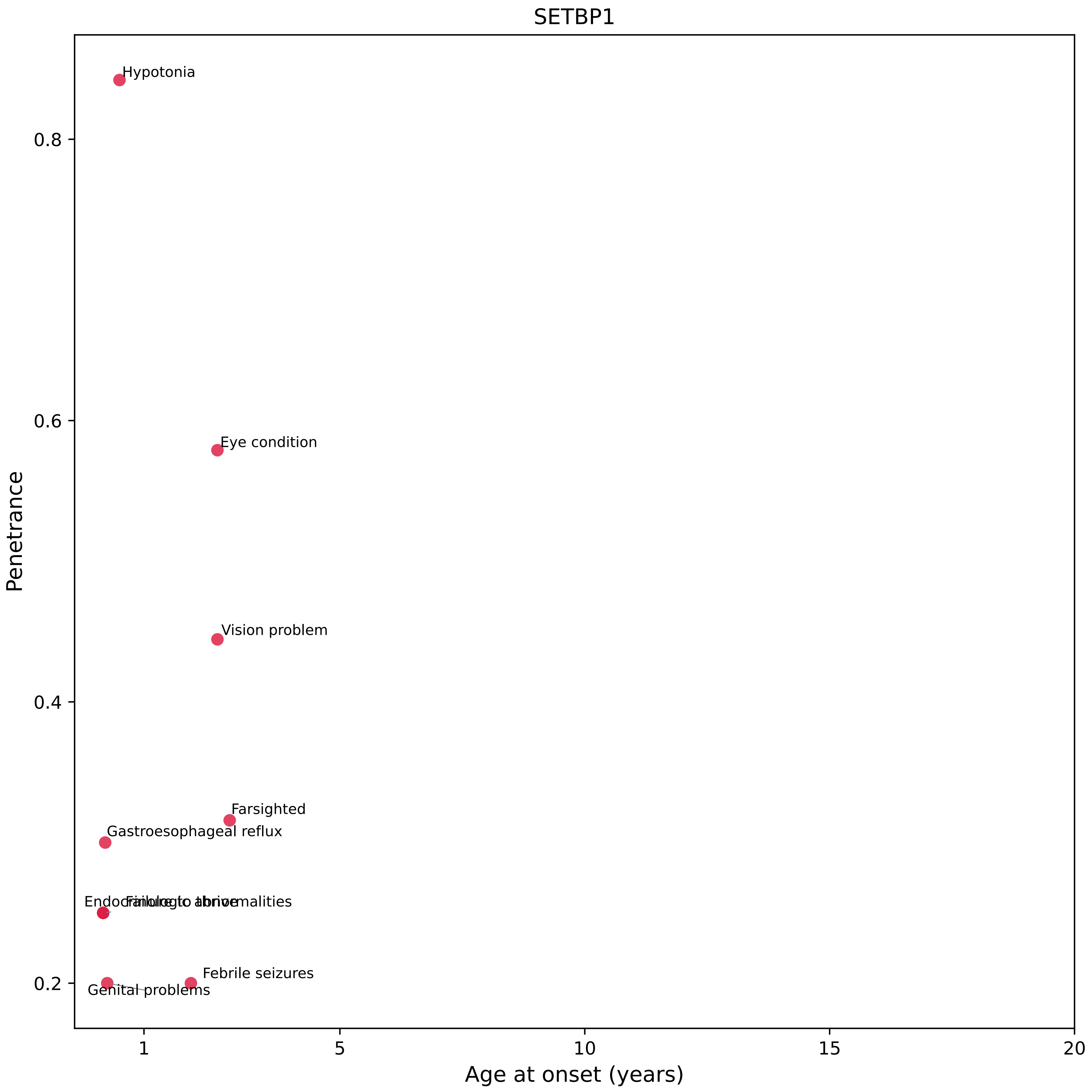
SETBP1
Hypotonia
Eye condition
Vision problem
Farsighted
Gastroesophageal reflux
Febrile seizures
Genital problems
0.8
0.6
0.4
0.2
Penetrance
1
5
10
15
20
Age at onset (years)

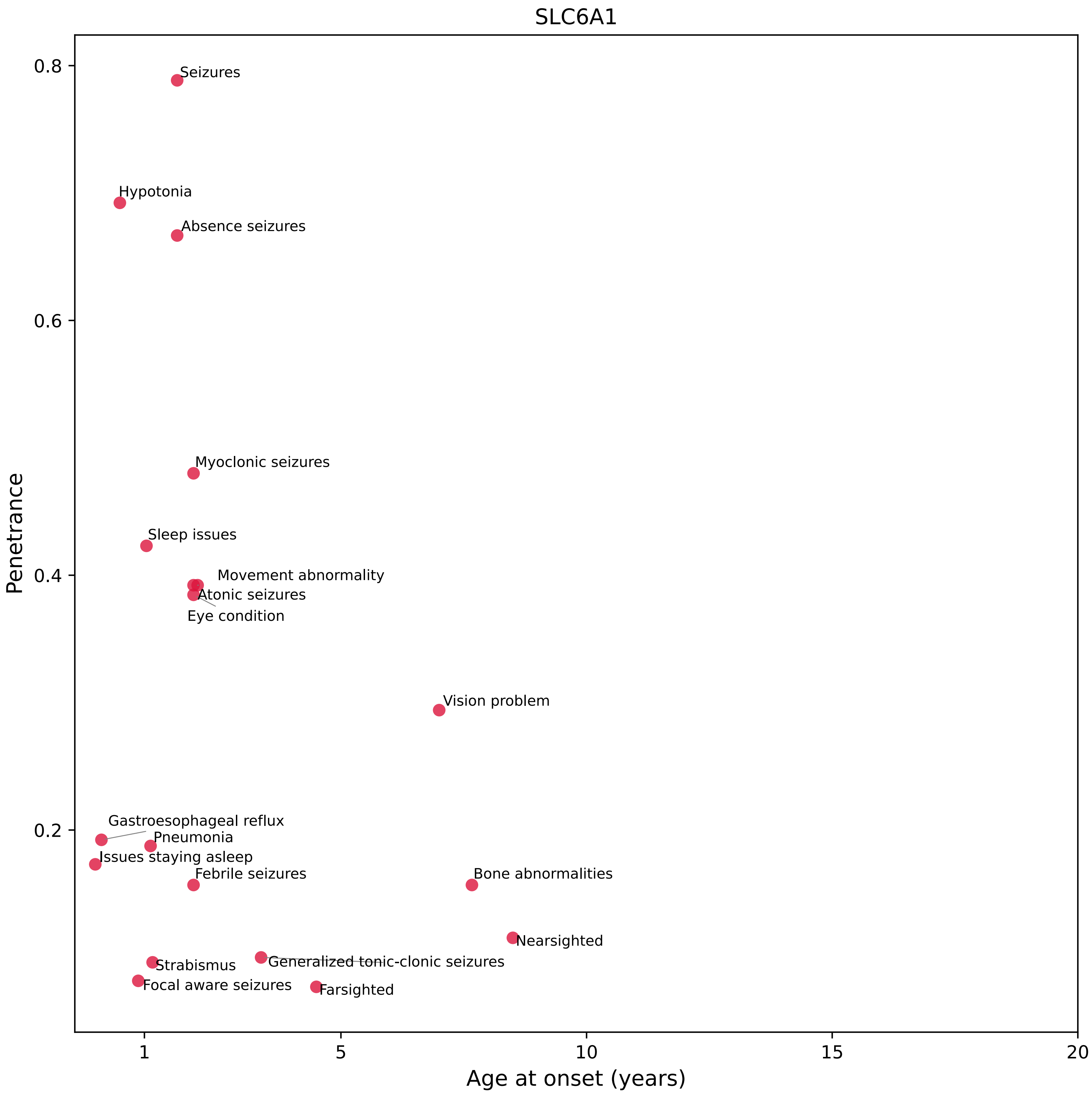
SLC6A1
Penetrance
Age at onset (years)
0.8
0.6
0.4
0.2
1
5
10
15
20
Seizures
Hypotonia
Absence seizures
Myoclonic seizures
Sleep issues
Movement abnormality
Atonic seizures
Eye condition
Vision problem
Gastroesophageal reflux
Pneumonia
Issues staying asleep
Febrile seizures
Bone abnormalities
Nearsighted
Strabismus
Generalized tonic-clonic seizures
Focal aware seizures
Farsighted

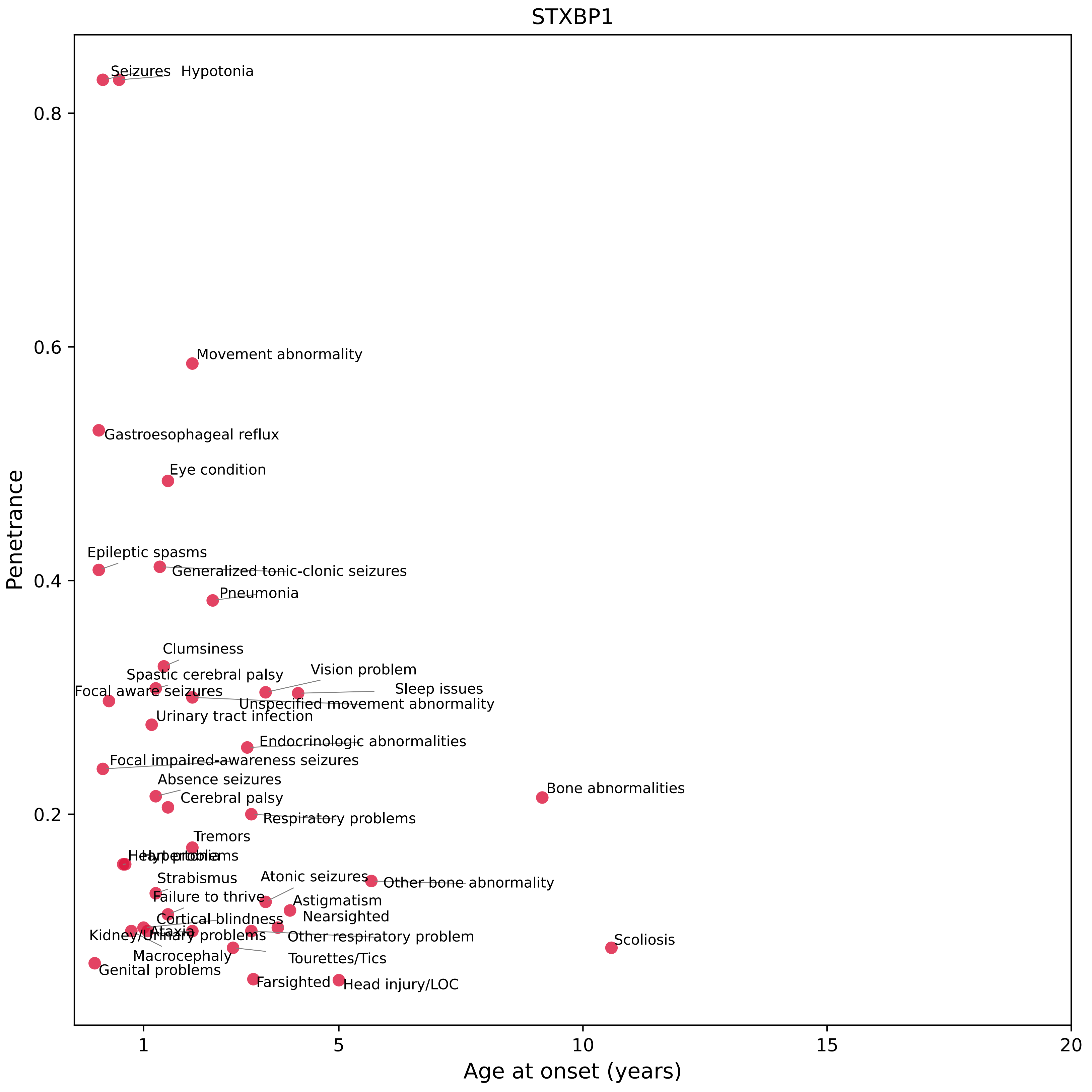

STXBP1
Penetrance
Age at onset (years)
0.8
0.6
0.4
0.2
1
5
10
15
20
Seizures
Hypotonia
Movement abnormality
Gastroesophageal reflux
Eye condition
Epileptic spasms
Generalized tonic-clonic seizures
Pneumonia
Clumsiness
Spastic cerebral palsy
Vision problem
Sleep issues
Focal aware seizures
Unspecified movement abnormality
Urinary tract infection
Endocrinologic abnormalities
Focal impaired-awareness seizures
Absence seizures
Bone abnormalities
Cerebral palsy
Respiratory problems
Tremors
Heart problems
Hypertonia
Strabismus
Atonic seizures
Other bone abnormality
Failure to thrive
Astigmatism
Nearsighted
Cortical blindness
Ataxia
Kidney/Urinary problems
Other respiratory problem
Scoliosis
Macrocephaly
Tourettes/Tics
Genital problems
Farsighted
Head injury/LOC

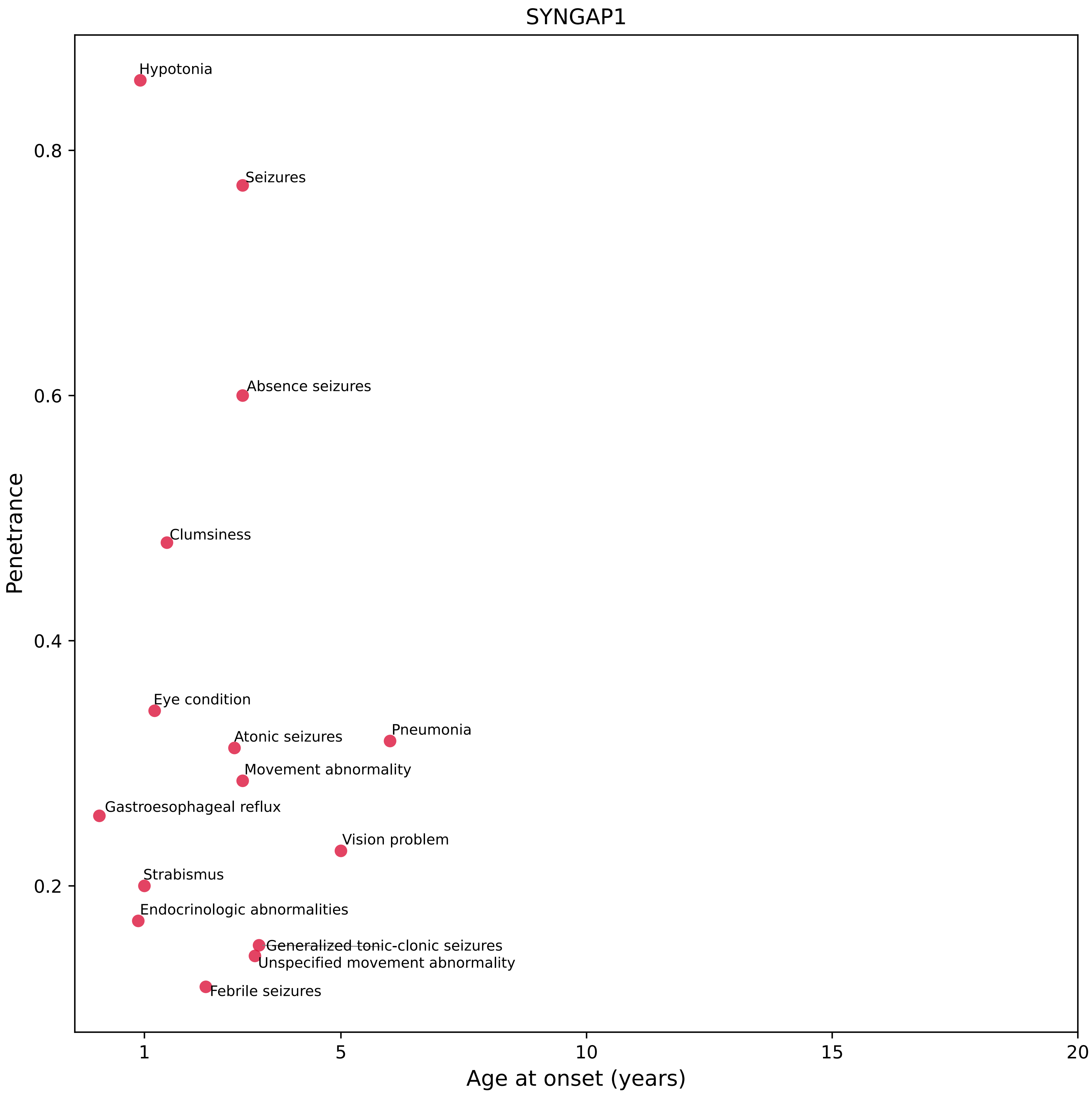

SYNGAP1
Penetrance
Age at onset (years)
0.2
0.4
0.6
0.8
1
5
10
15
20
Hypotonia
Seizures
Absence seizures
Clumsiness
Eye condition
Atonic seizures
Pneumonia
Movement abnormality
Gastroesophageal reflux
Vision problem
Strabismus
Endocrinologic abnormalities
Generalized tonic-clonic seizures
Unspecified movement abnormality
Febrile seizures

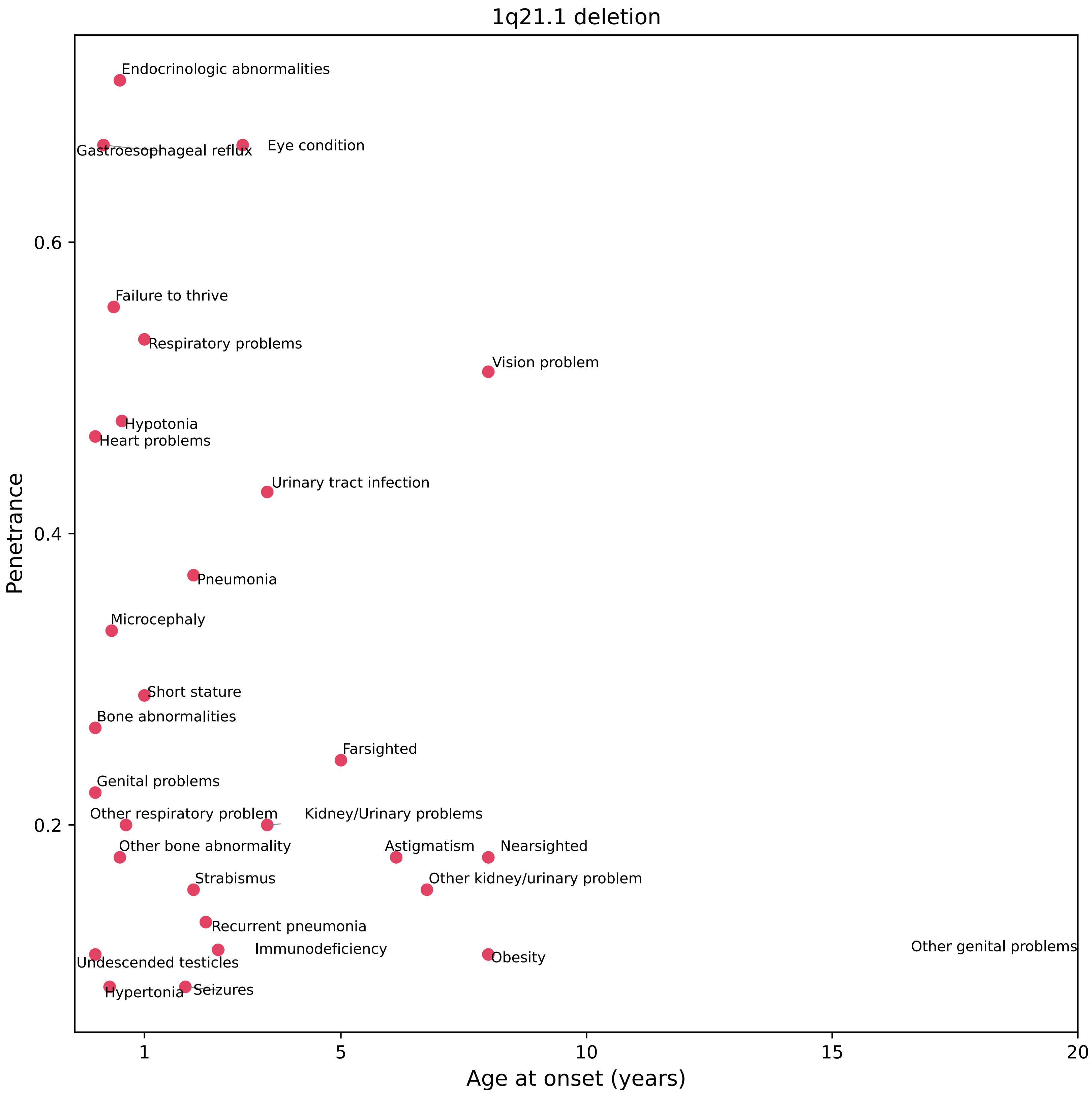

1q21.1 deletion
Endocrinologic abnormalities
Gastroesophageal reflux
Eye condition
Failure to thrive
Respiratory problems
Vision problem
Hypotonia
Heart problems
Urinary tract infection
Pneumonia
Microcephaly
Short stature
Bone abnormalities
Farsighted
Genital problems
Other respiratory problem
Kidney/Urinary problems
Other bone abnormality
Astigmatism
Nearsighted
Strabismus
Other kidney/urinary problem
Recurrent pneumonia
Immunodeficiency
Other genital problems
Undescended testicles
Obesity
Hypertonia
Seizures
Penetrance
0.6
0.4
0.2
Age at onset (years)
1
5
10
15
20

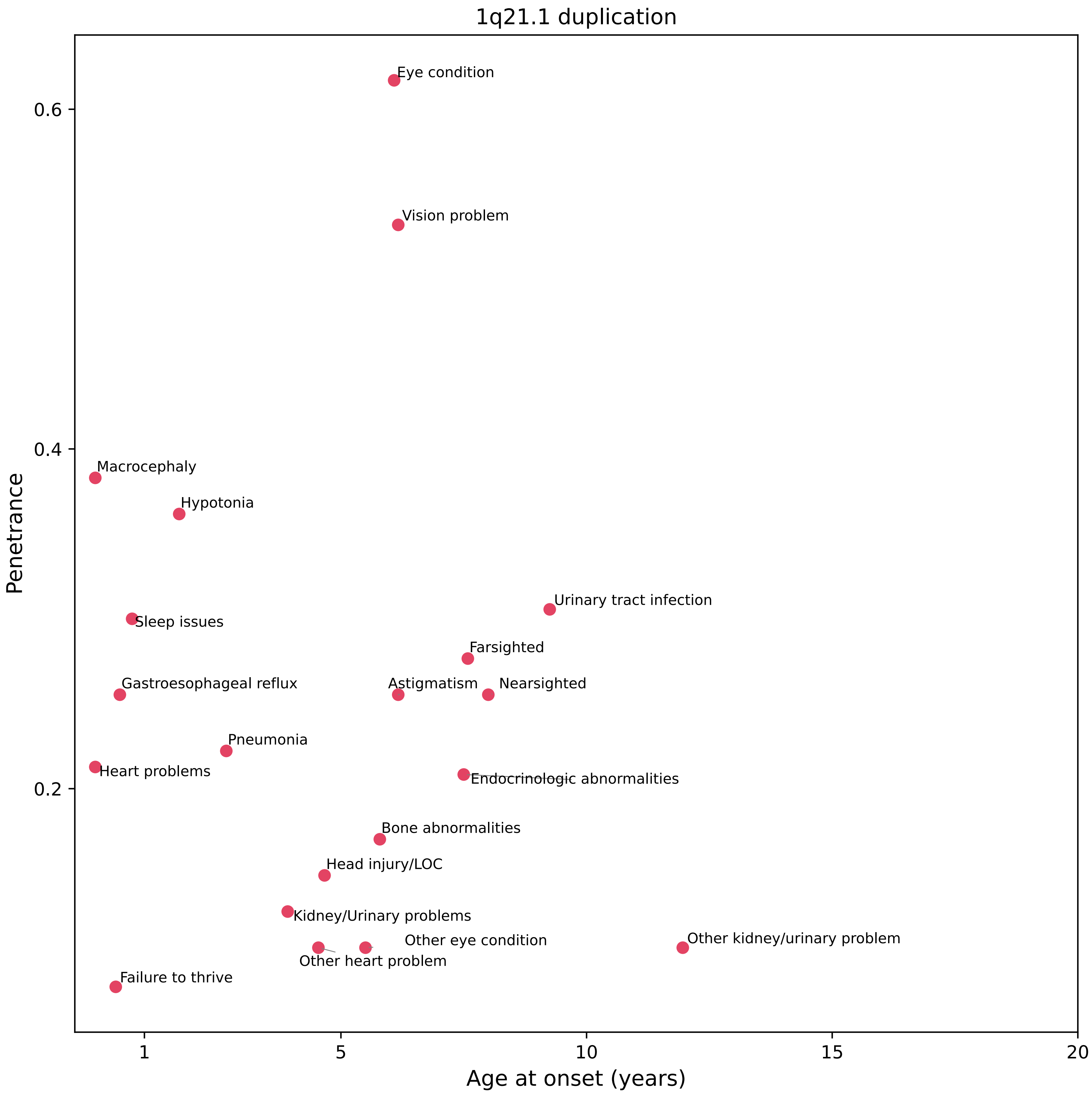
1q21.1 duplication
Penetrance
Age at onset (years)
0.6
0.4
0.2
1
5
10
15
20
Eye condition
Vision problem
Macrocephaly
Hypotonia
Urinary tract infection
Sleep issues
Farsighted
Gastroesophageal reflux
Astigmatism
Nearsighted
Pneumonia
Heart problems
Endocrinologic abnormalities
Bone abnormalities
Head injury/LOC
Kidney/Urinary problems
Other eye condition
Other kidney/urinary problem
Other heart problem
Failure to thrive

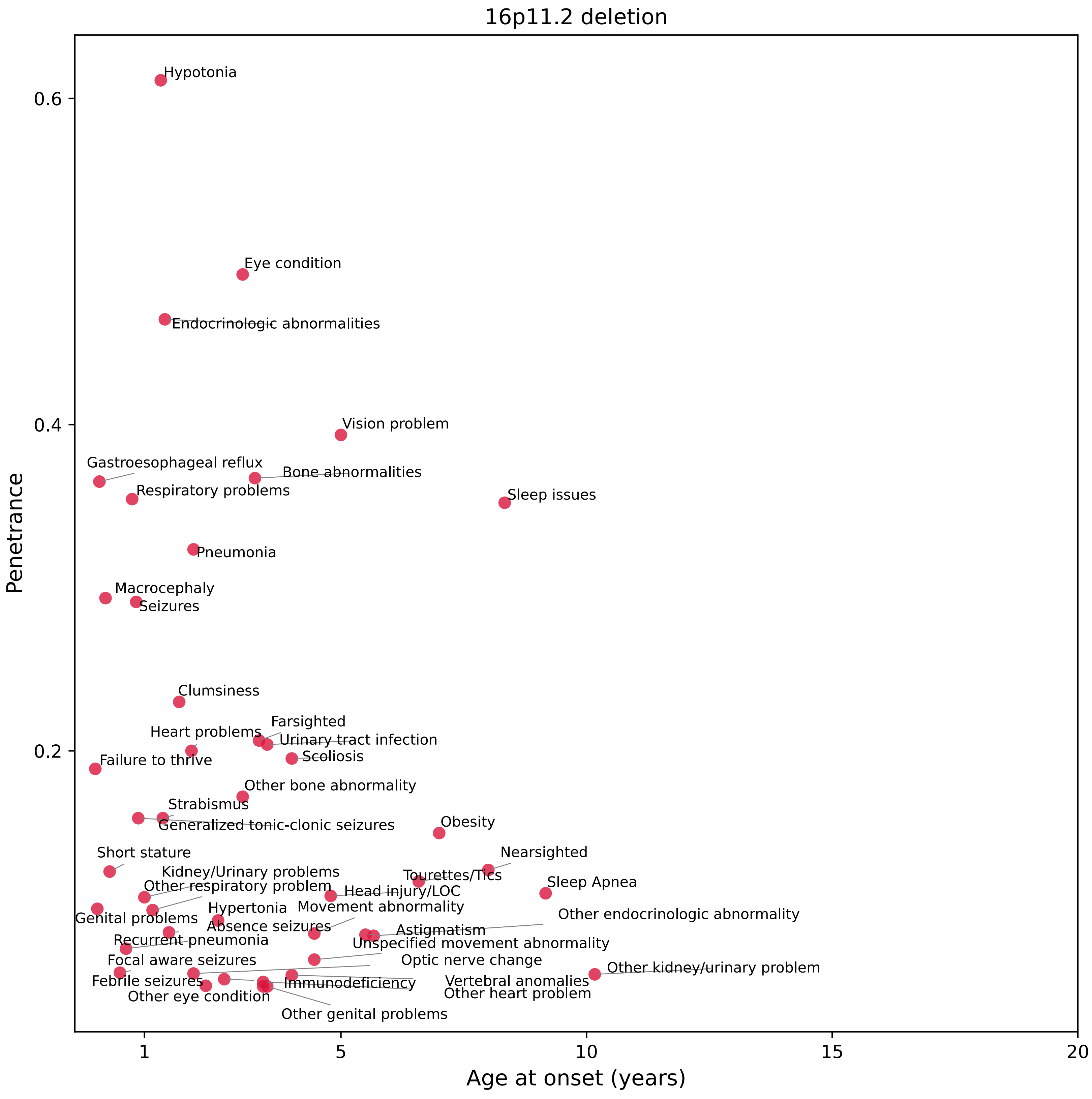

16p11.2 deletion
Penetrance
Age at onset (years)
0.6
0.4
0.2
1
5
10
15
20
Hypotonia
Eye condition
Endocrinologic abnormalities
Vision problem
Gastroesophageal reflux
Bone abnormalities
Respiratory problems
Sleep issues
Pneumonia
Macrocephaly
Seizures
Clumsiness
Farsighted
Heart problems
Urinary tract infection
Scoliosis
Failure to thrive
Other bone abnormality
Strabismus
Generalized tonic-clonic seizures
Obesity
Short stature
Nearsighted
Kidney/Urinary problems
Tourettes/Tics
Other respiratory problem
Head injury/LOC
Sleep Apnea
Hypertonia
Movement abnormality
Genital problems
Absence seizures
Other endocrinologic abnormality
Astigmatism
Recurrent pneumonia
Unspecified movement abnormality
Focal aware seizures
Optic nerve change
Other kidney/urinary problem
Febrile seizures
Immunodeficiency
Vertebral anomalies
Other eye condition
Other heart problem
Other genital problems

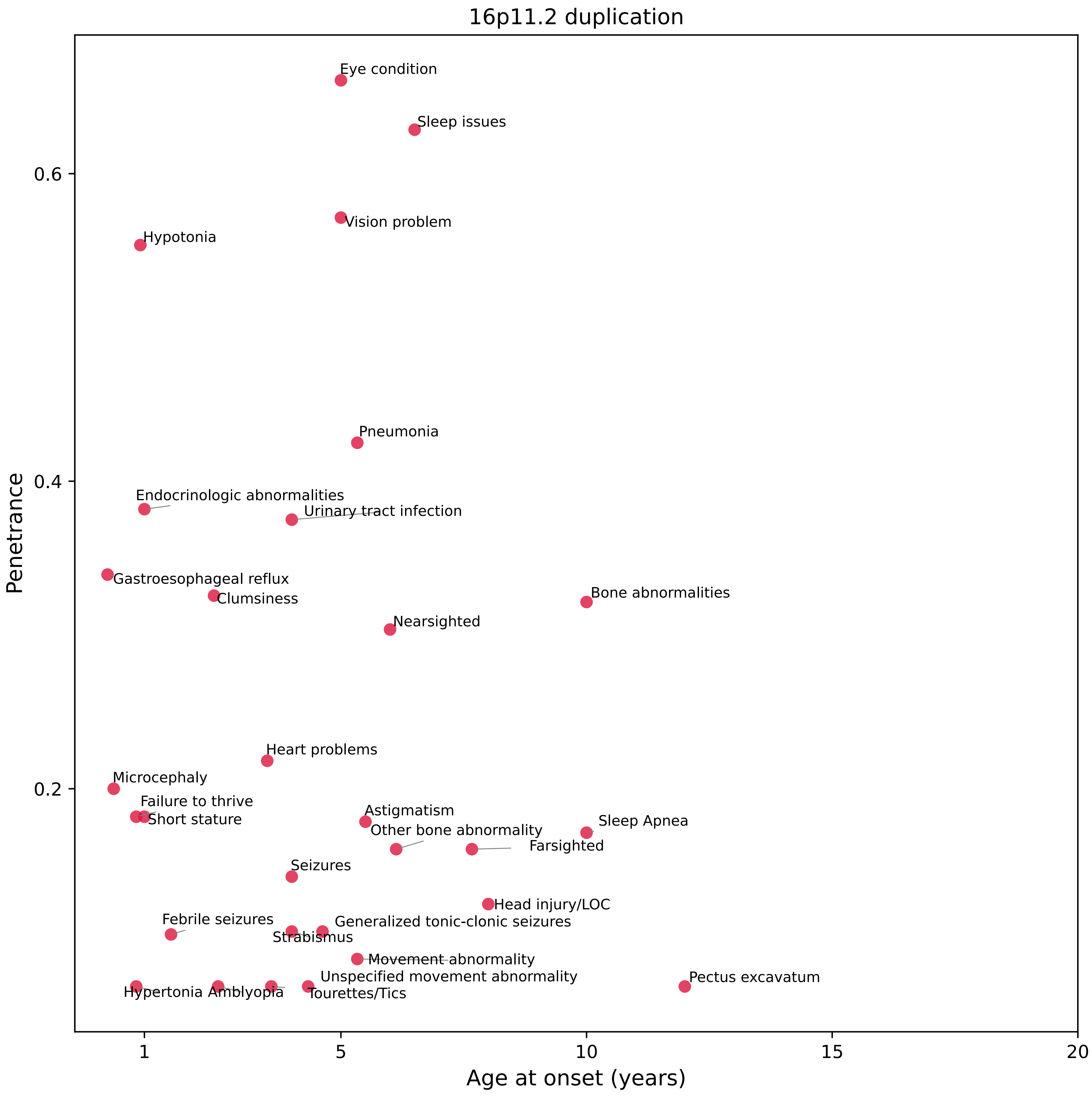

16p11.2 duplication
Penetrance
Age at onset (years)
0.2
0.4
0.6
1
5
10
15
20
Eye condition
Sleep issues
Vision problem
Hypotonia
Pneumonia
Endocrinologic abnormalities
Urinary tract infection
Gastroesophageal reflux
Clumsiness
Bone abnormalities
Nearsighted
Heart problems
Microcephaly
Failure to thrive
Short stature
Astigmatism
Other bone abnormality
Sleep Apnea
Farsighted
Seizures
Head injury/LOC
Febrile seizures
Generalized tonic-clonic seizures
Strabismus
Movement abnormality
Unspecified movement abnormality
Pectus excavatum
Hypertonia
Amblyopia
Tourettes/Tics

### Cumulative Penetrance for ASXL3 (page 1/3)

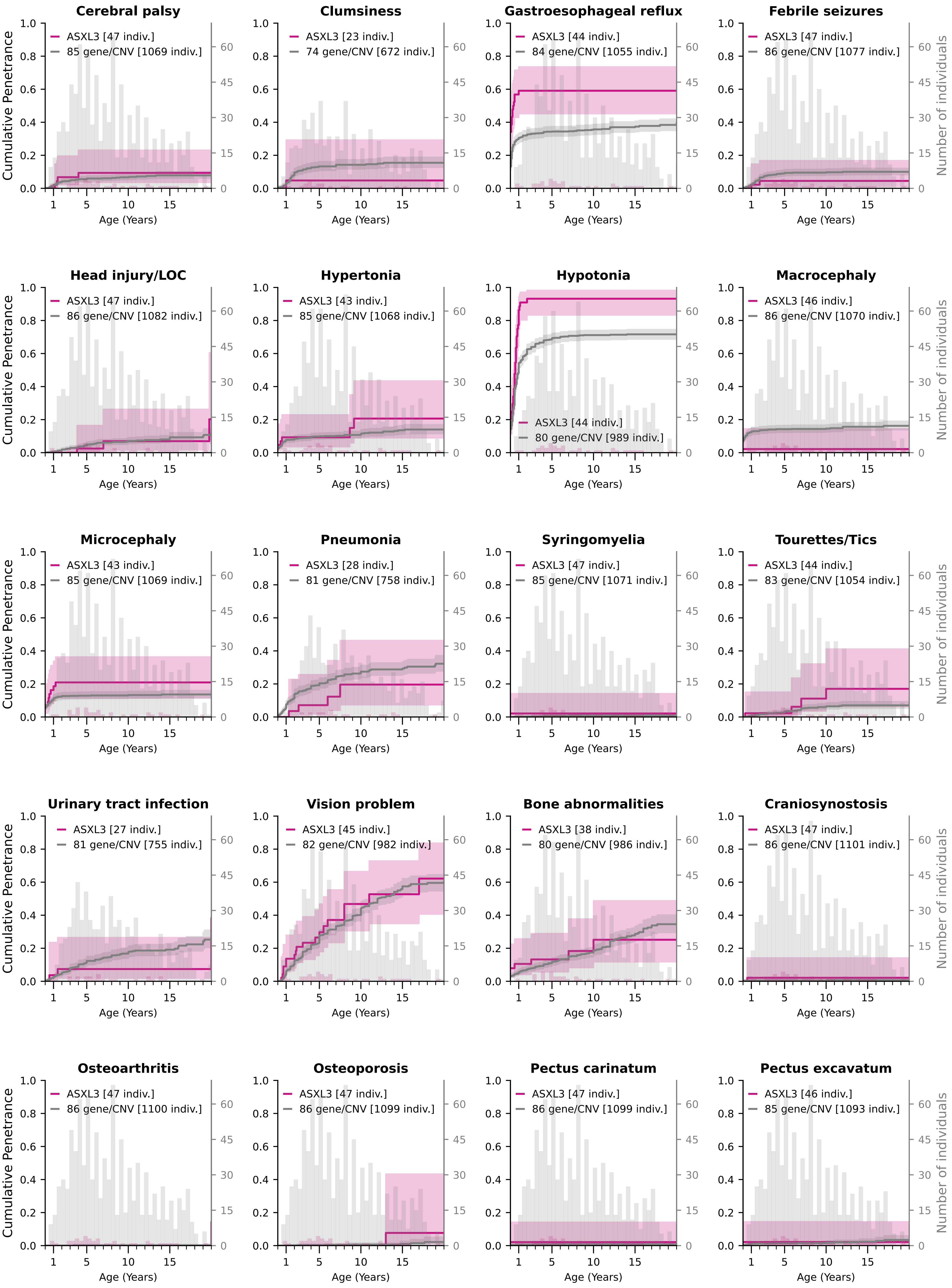

### Cumulative Penetrance for ASXL3 (page 2/3)

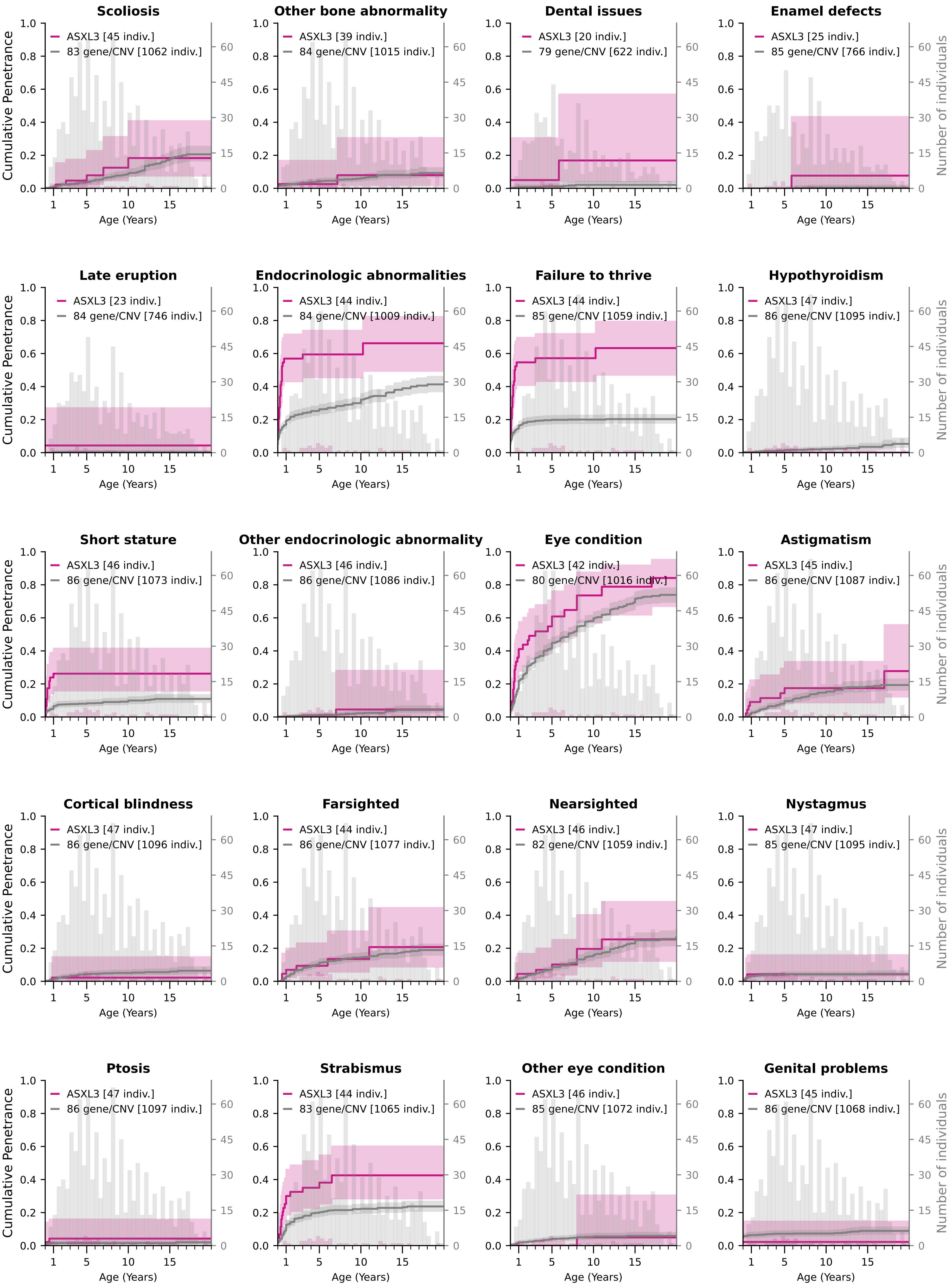

### Cumulative Penetrance for ASXL3 (page 3/3)

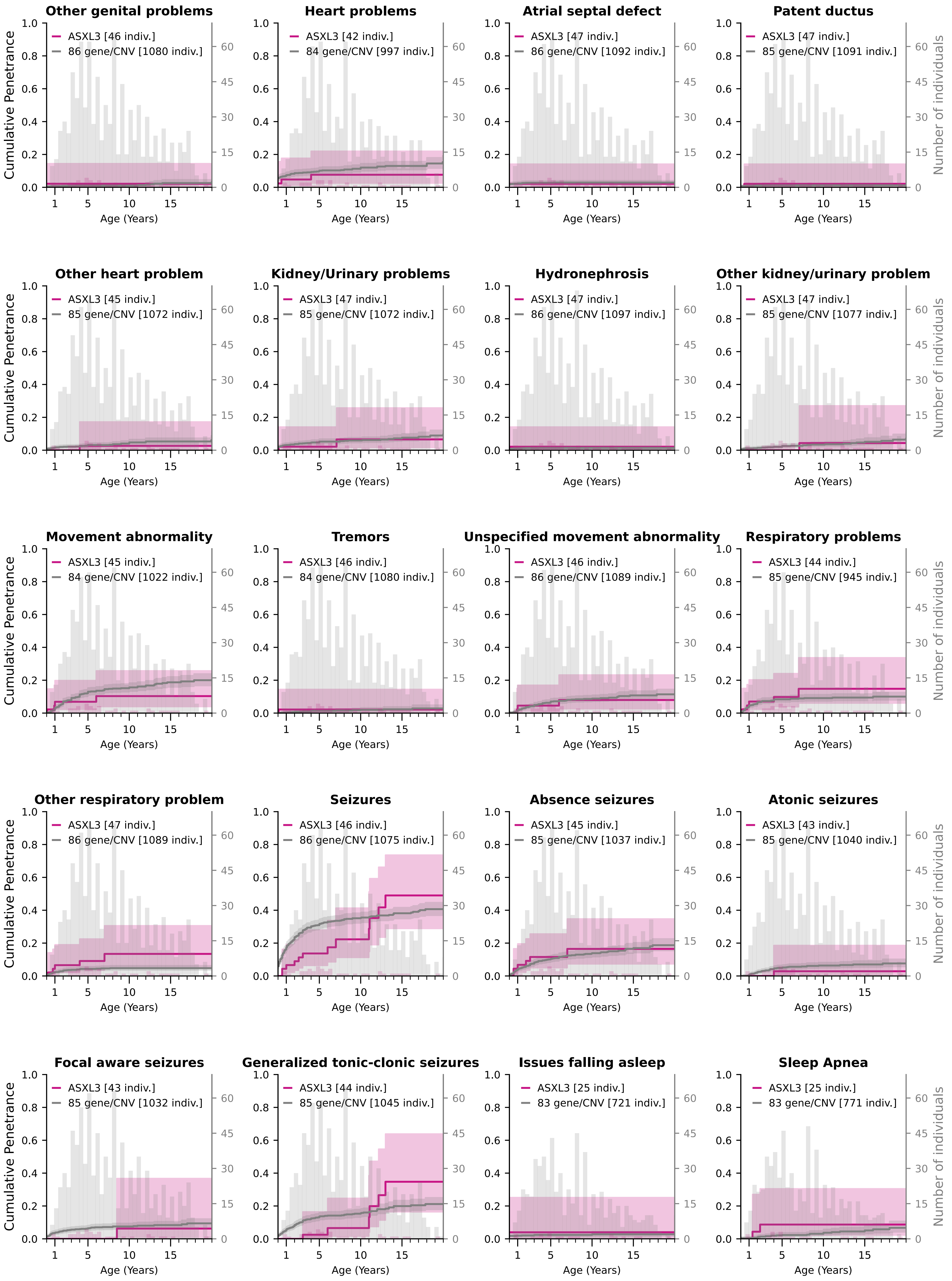

### Cumulative Penetrance for CHAMP1 (page 1/2)

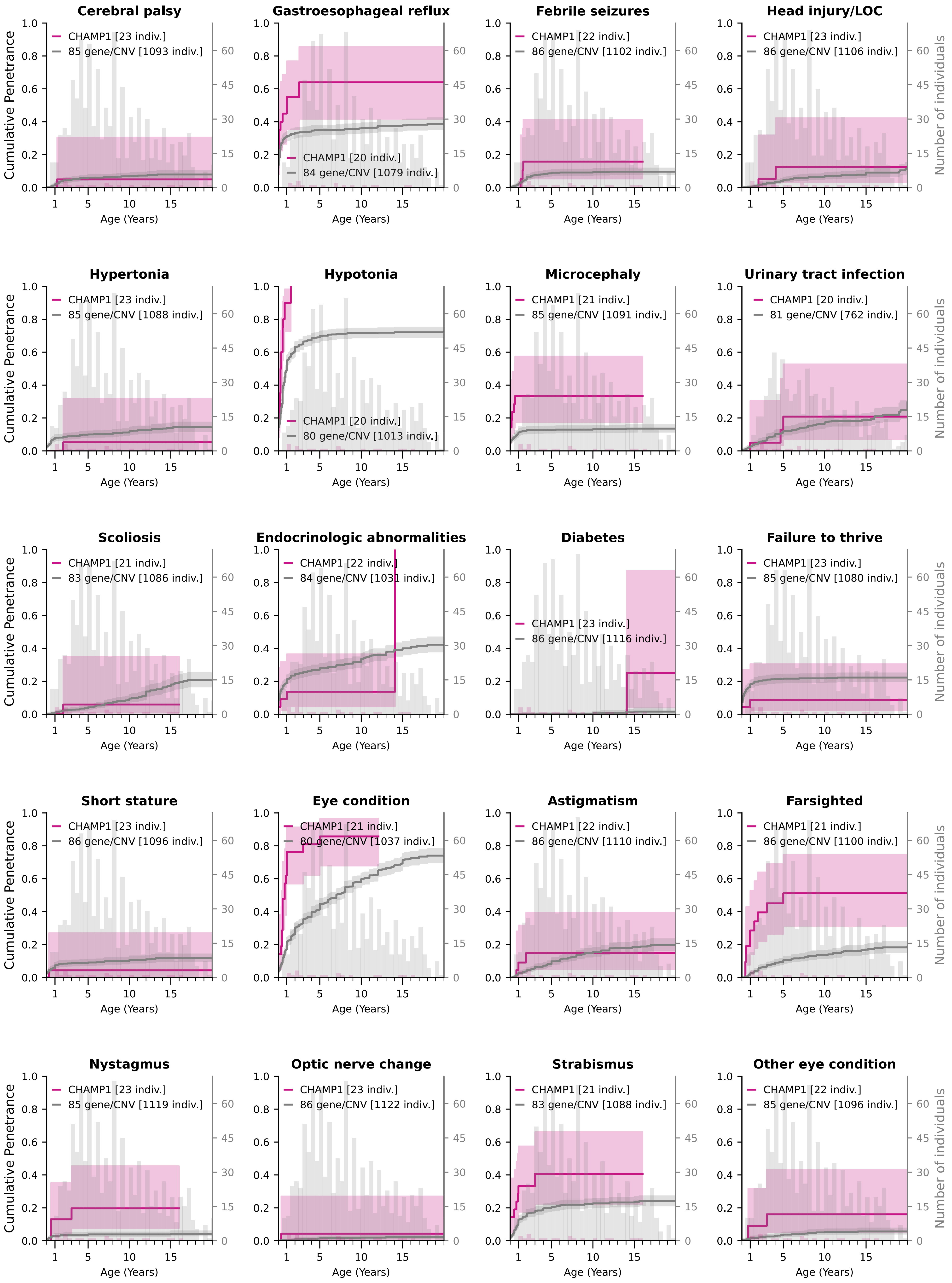

### Cumulative Penetrance for CHAMP1 (page 2/2)

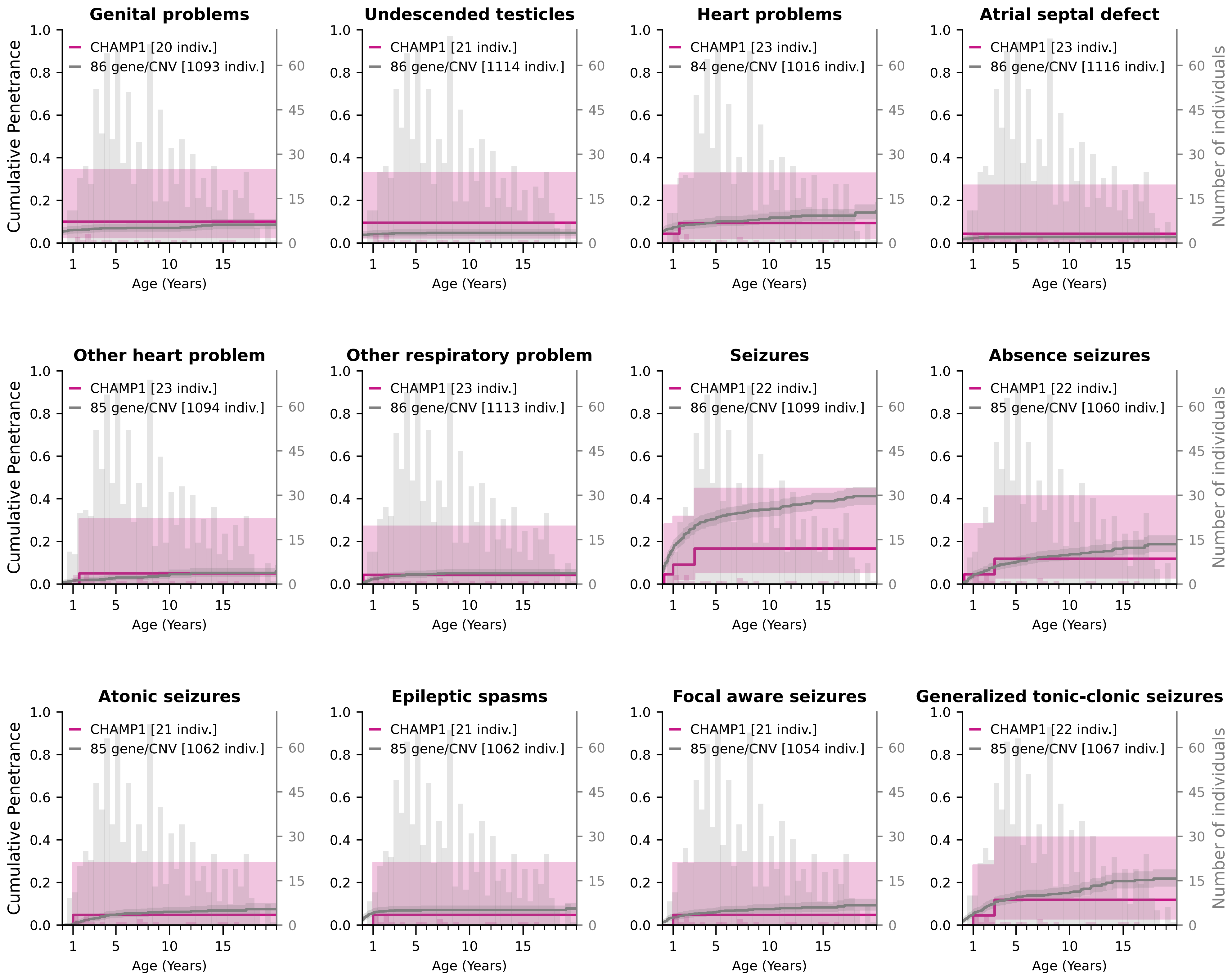

### Cumulative Penetrance for CSNK2A1 (page 1/3)

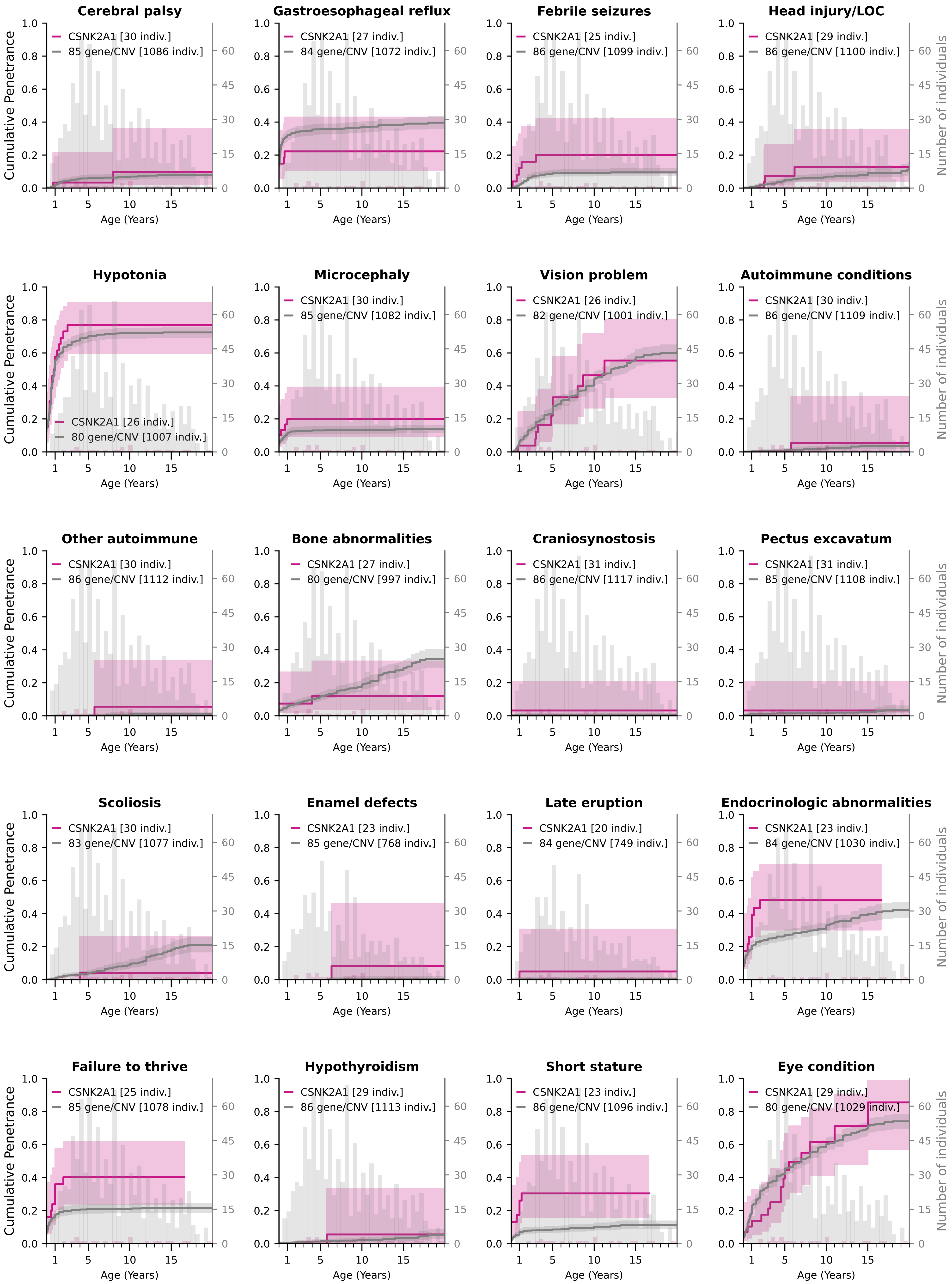

### Cumulative Penetrance for CSNK2A1 (page 2/3)

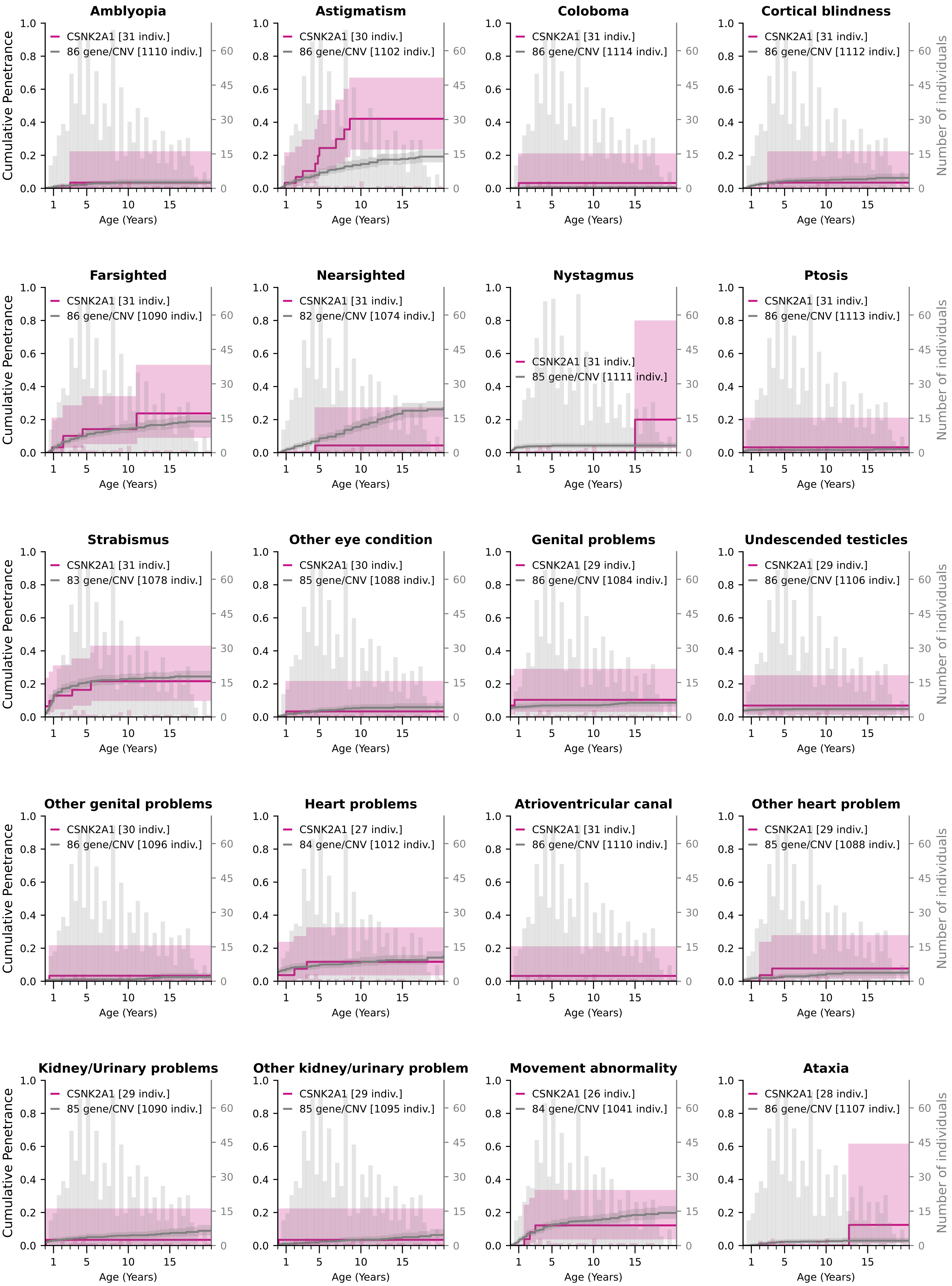

### Cumulative Penetrance for CSNK2A1 (page 3/3)

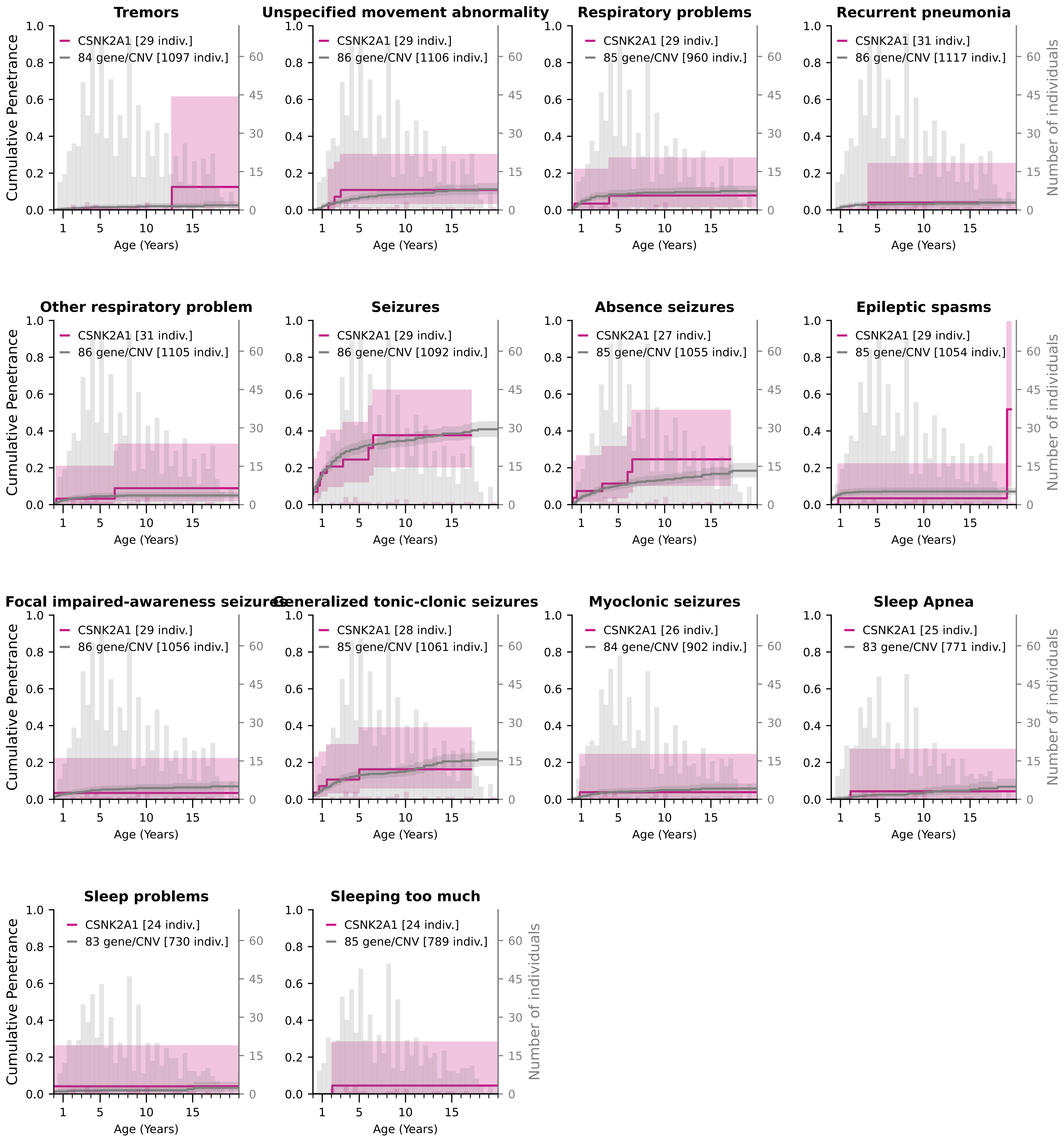

### Cumulative Penetrance for CTNNB1 (page 1/3)

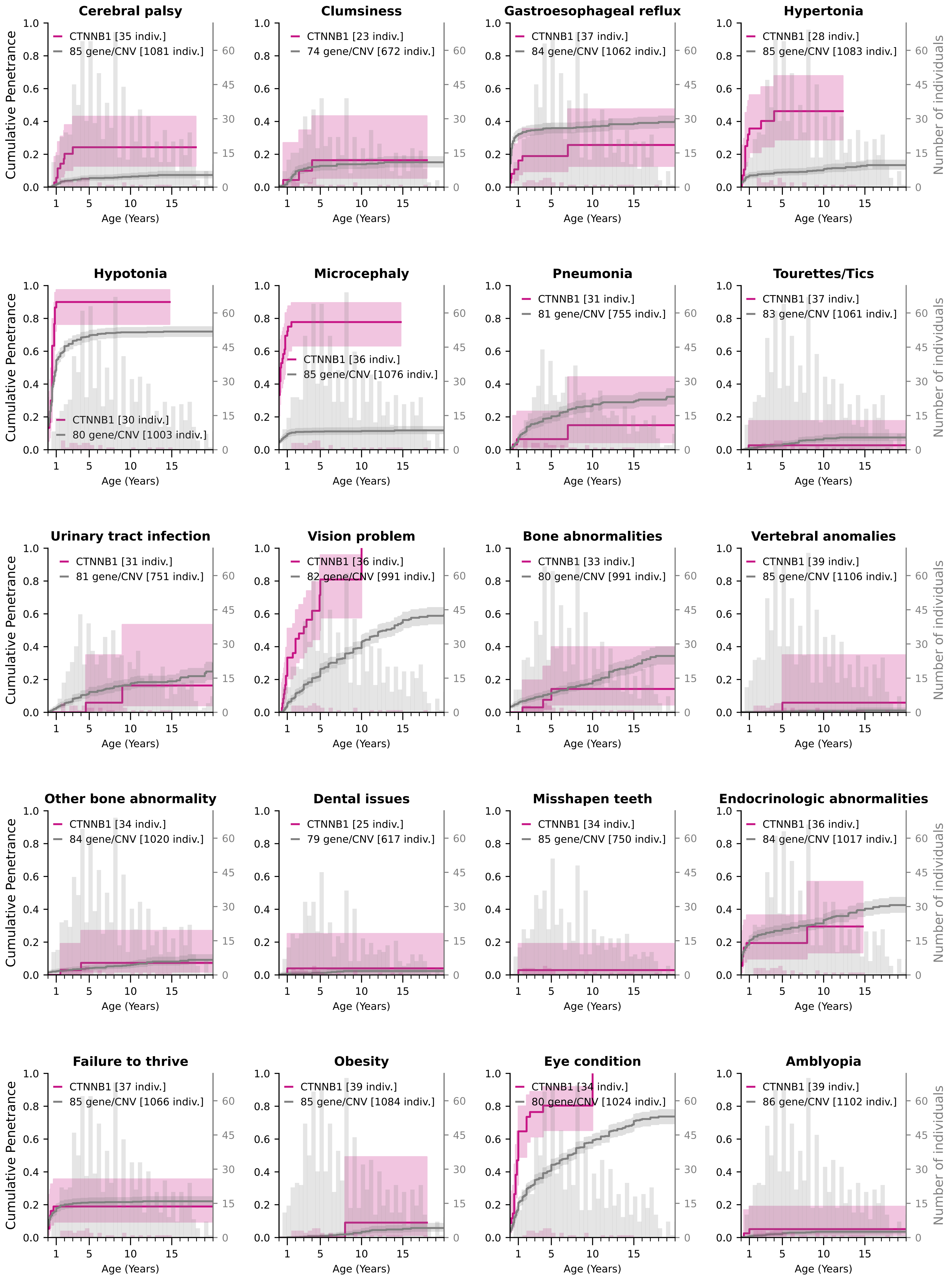

### Cumulative Penetrance for CTNNB1 (page 2/3)

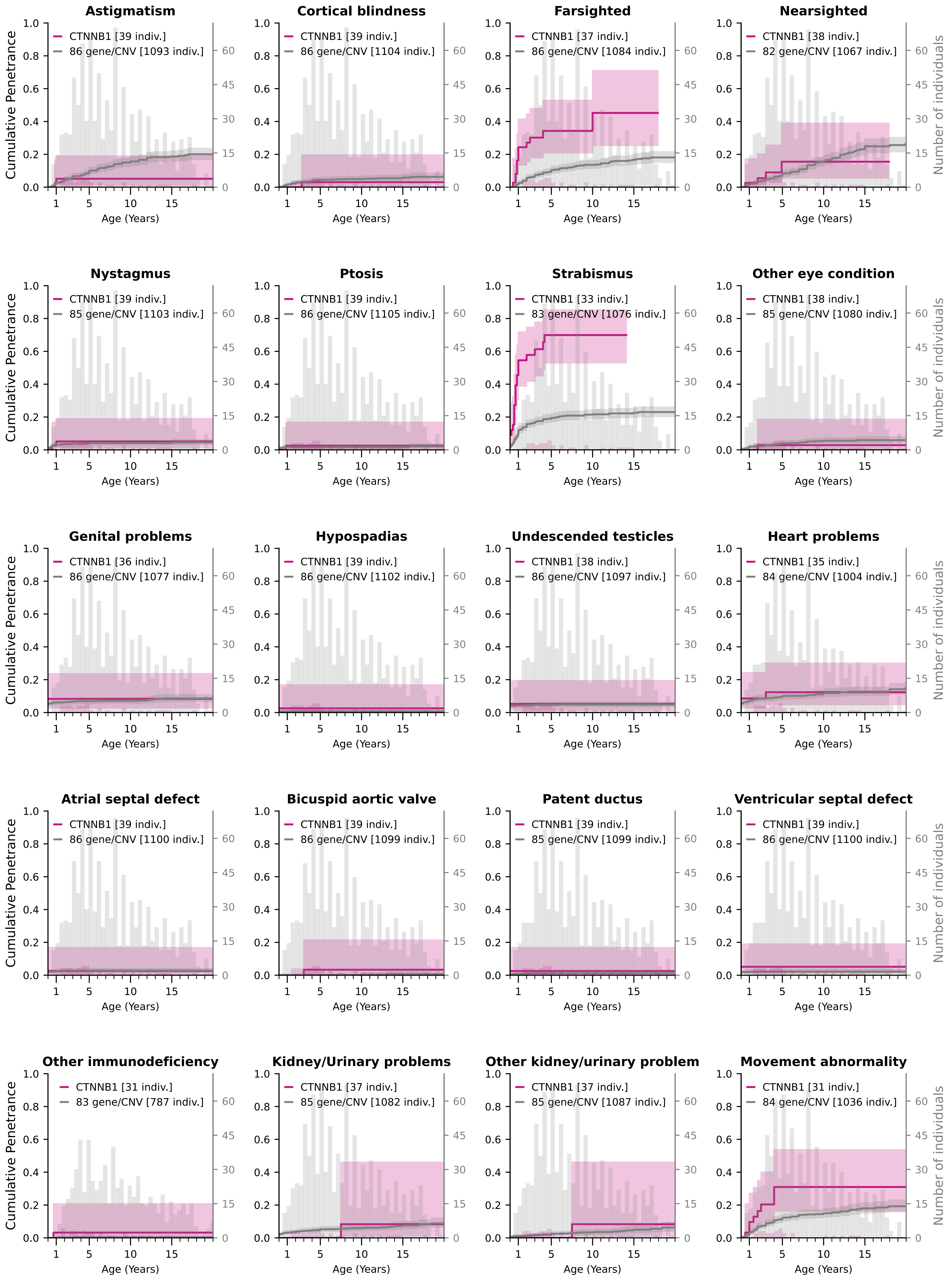

### Cumulative Penetrance for CTNNB1 (page 3/3)

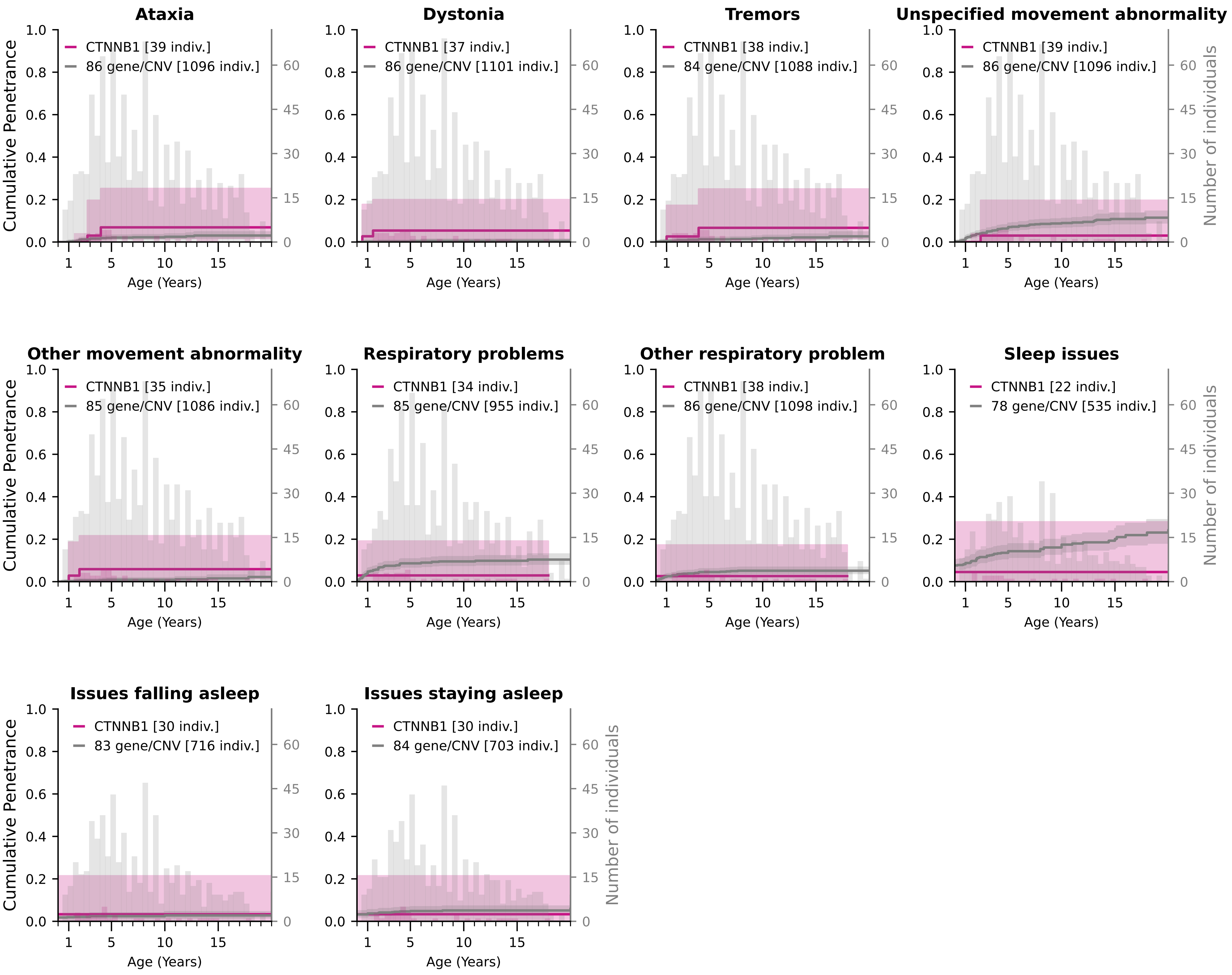

### Cumulative Penetrance for DYRK1A (page 1/3)

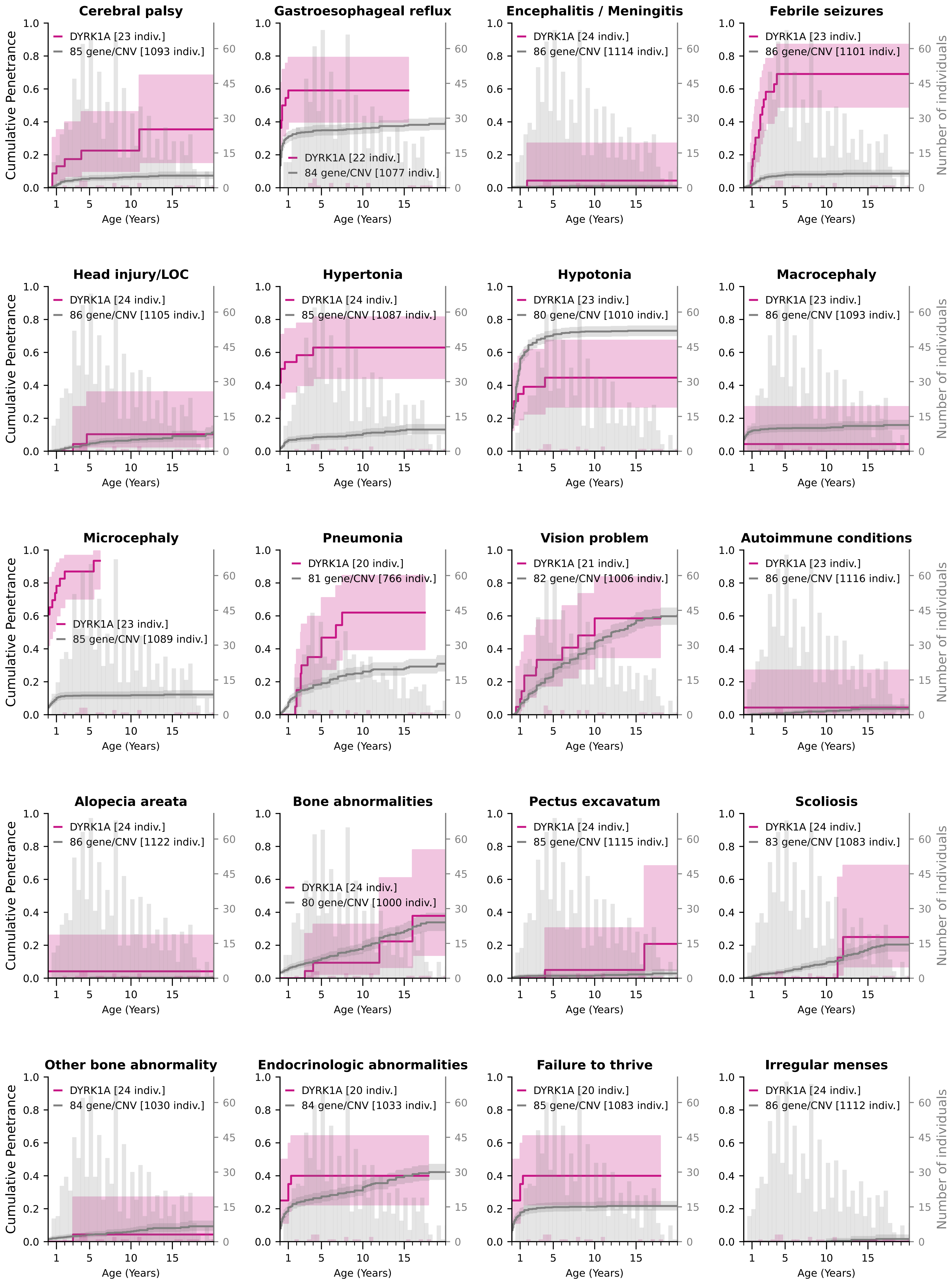

### Cumulative Penetrance for DYRK1A (page 2/3)

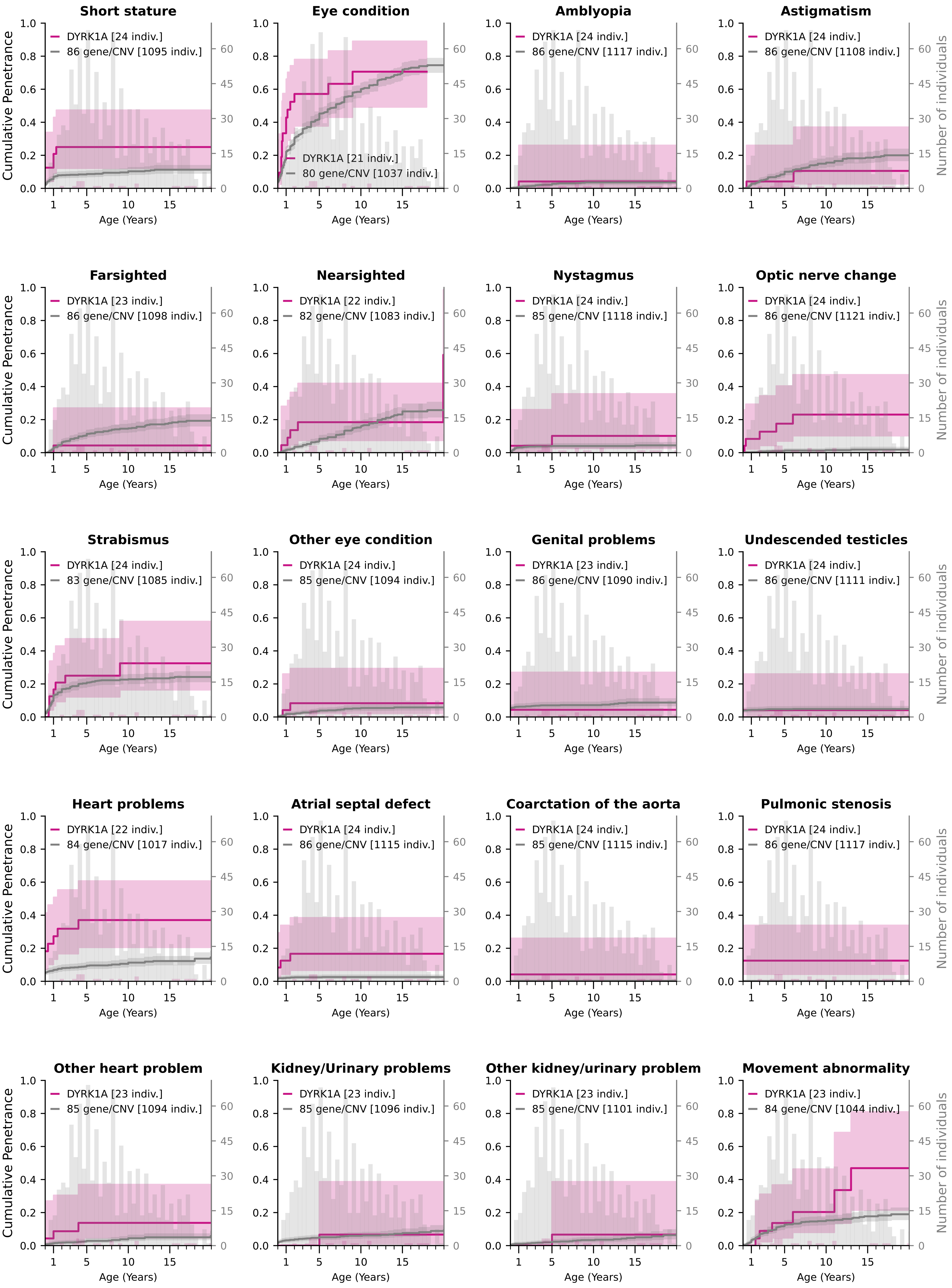

### Cumulative Penetrance for DYRK1A (page 3/3)

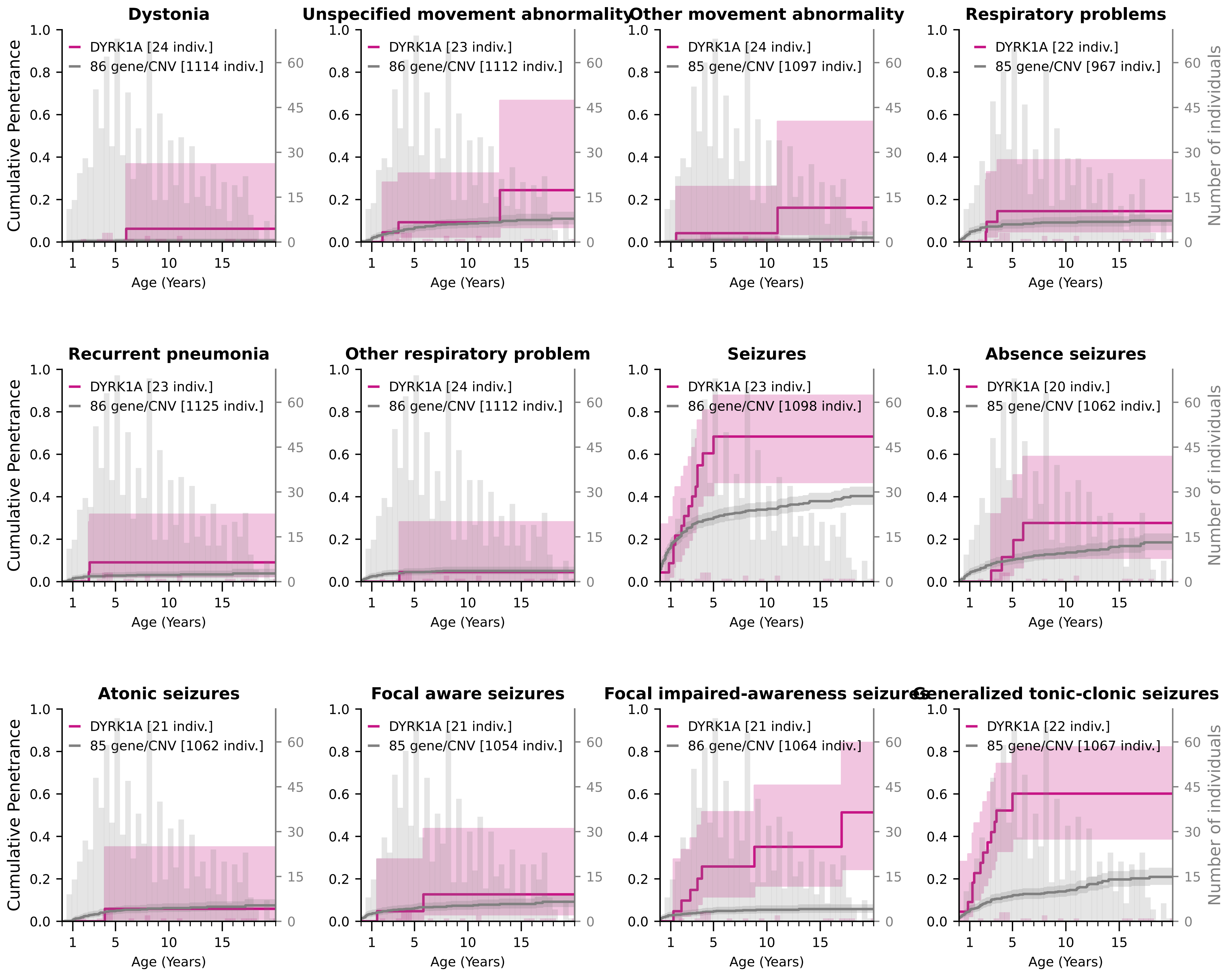

### Cumulative Penetrance for GRIN2B (page 1/3)

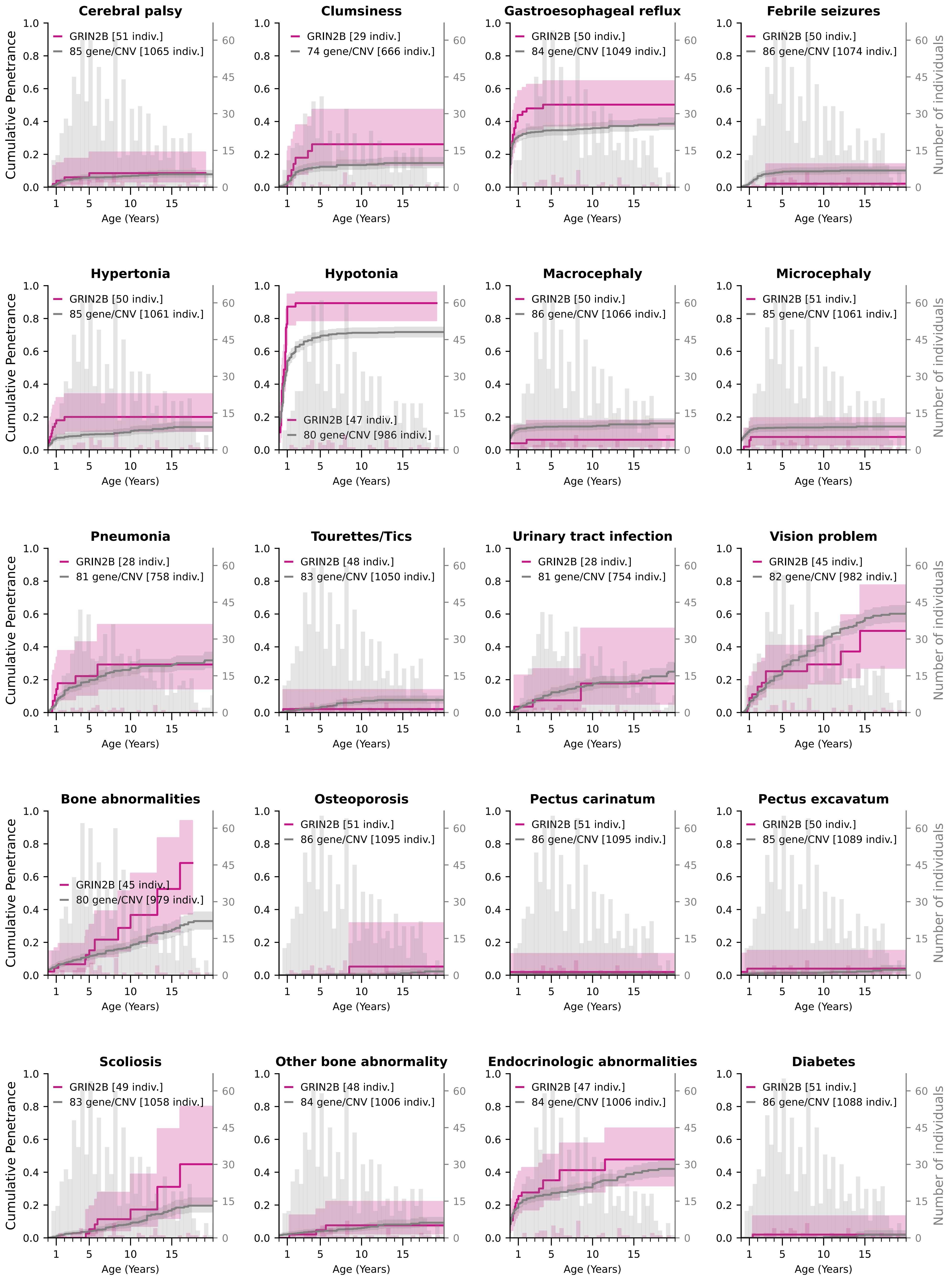

### Cumulative Penetrance for GRIN2B (page 2/3)

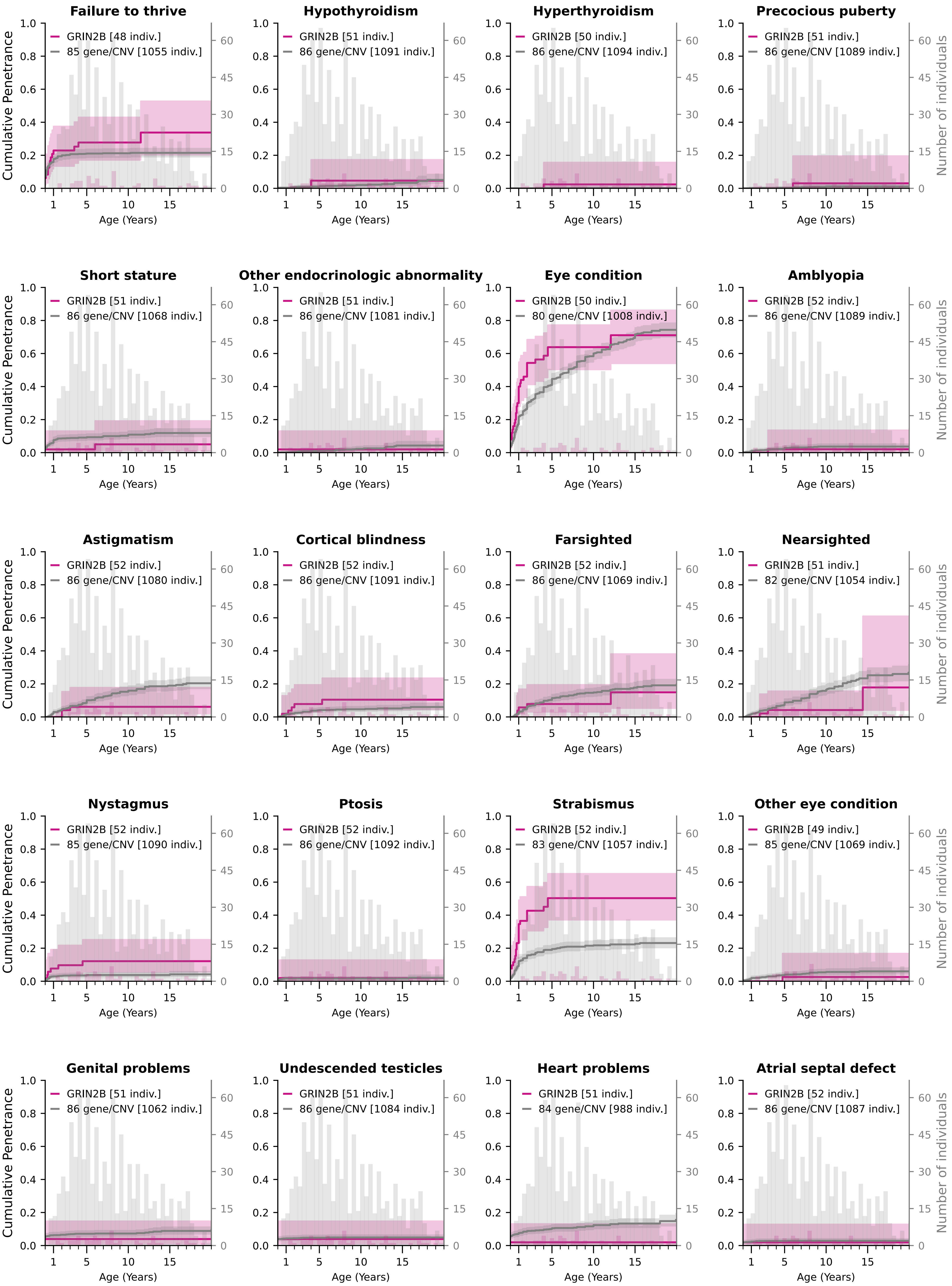

### Cumulative Penetrance for GRIN2B (page 3/3)

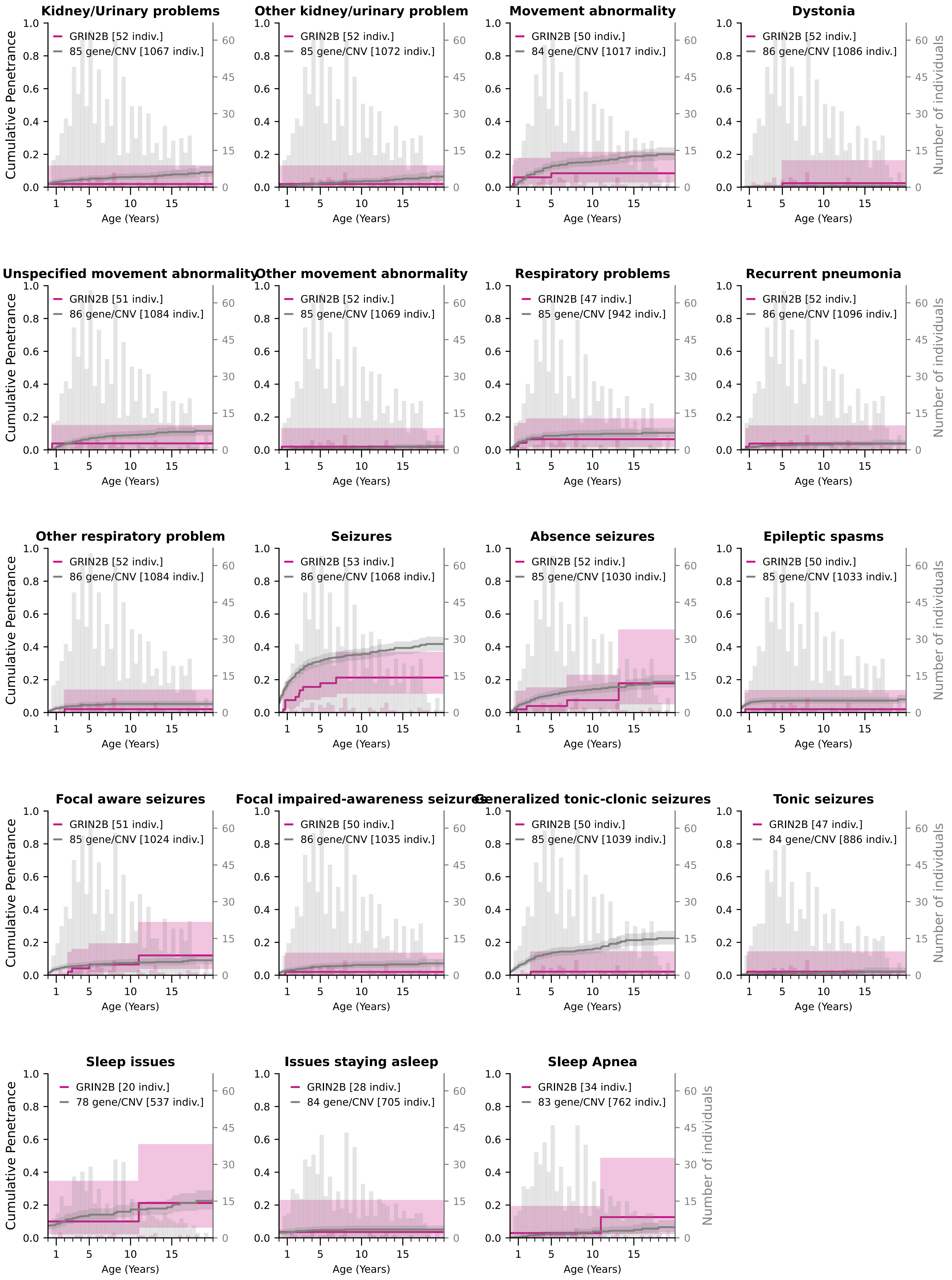

### Cumulative Penetrance for HNRNPH2 (page 1/2)

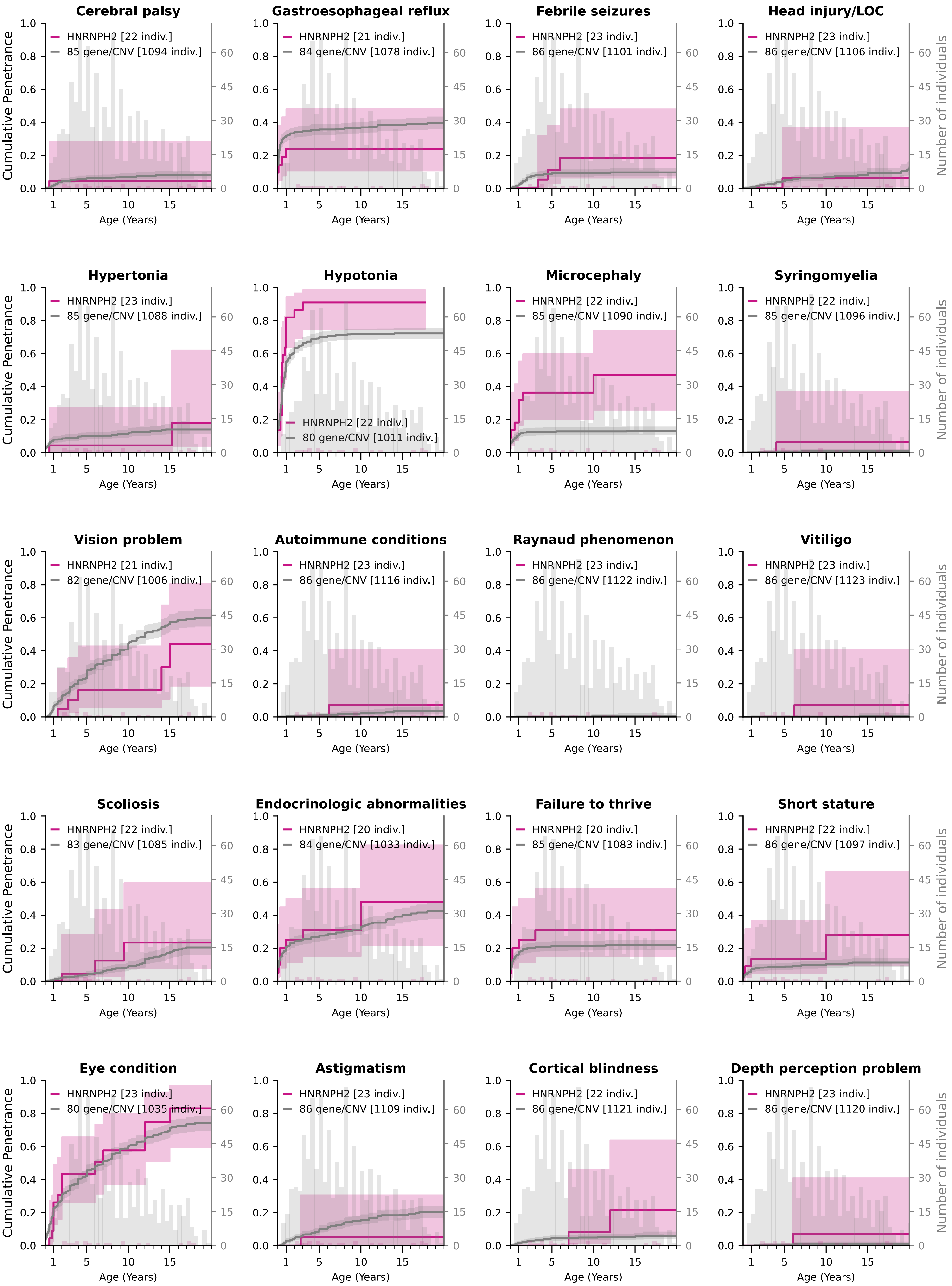

### Cumulative Penetrance for HNRNPH2 (page 2/2)

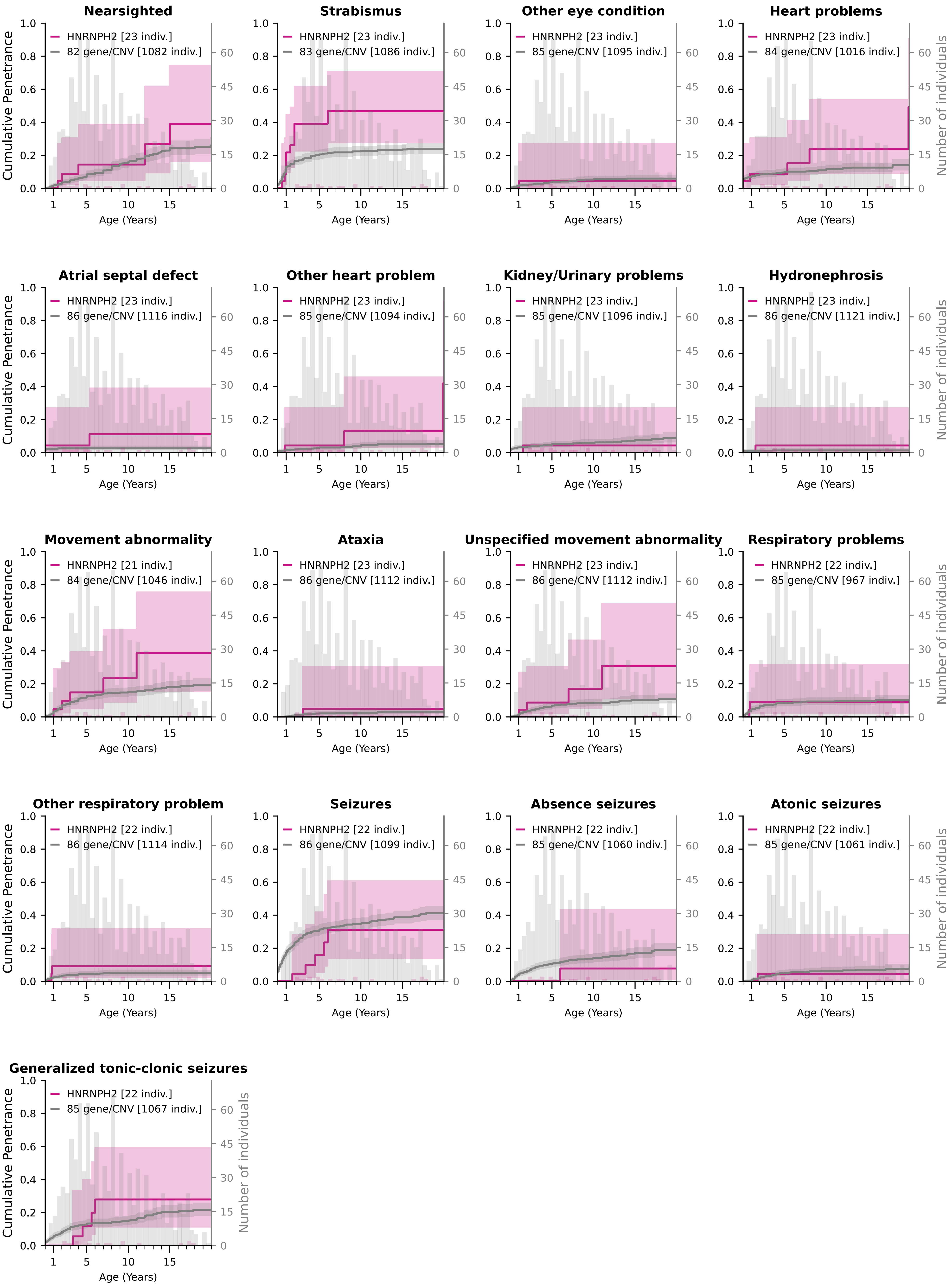

### Cumulative Penetrance for MED13L (page 1/2)

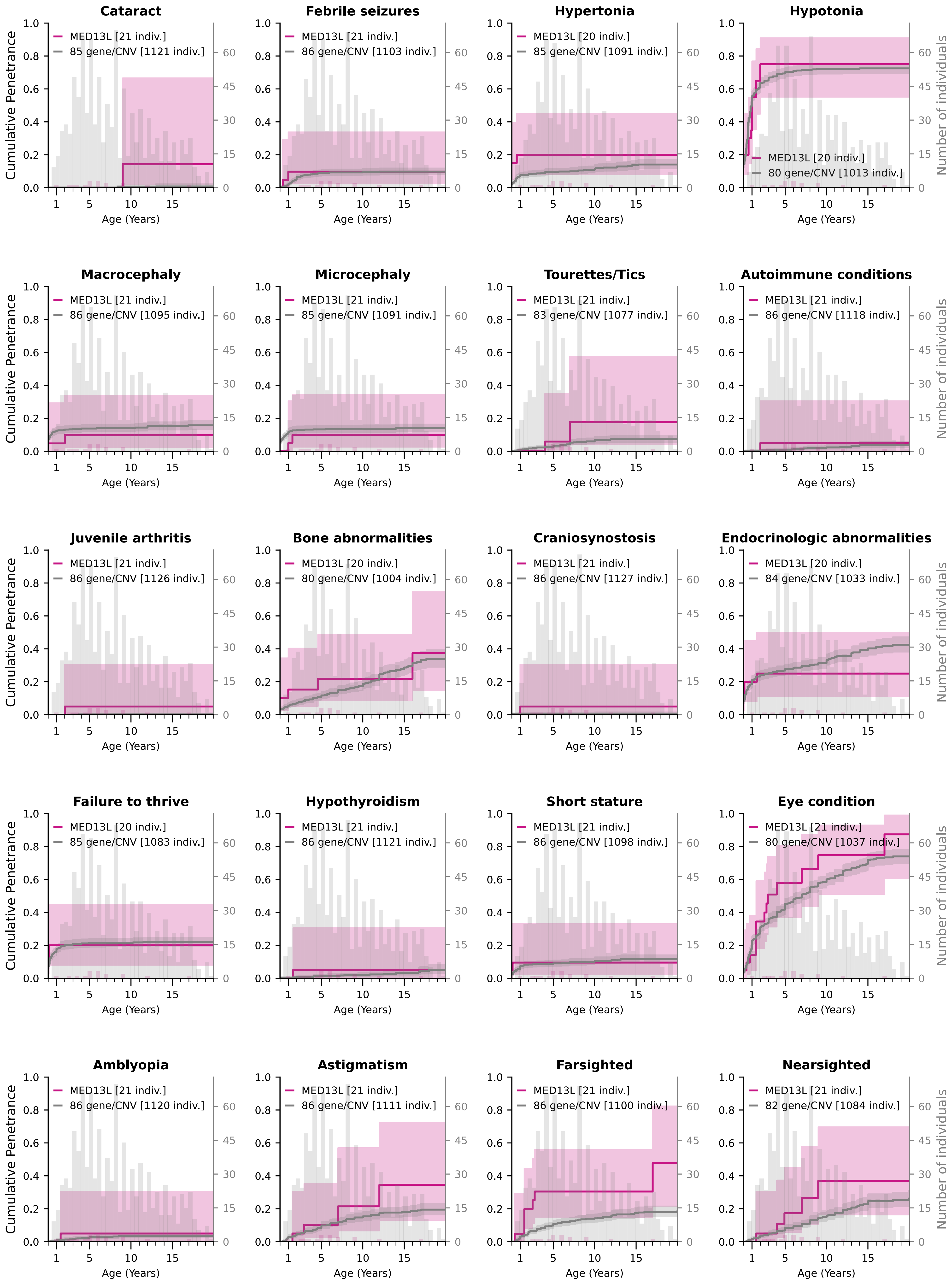

### Cumulative Penetrance for MED13L (page 2/2)

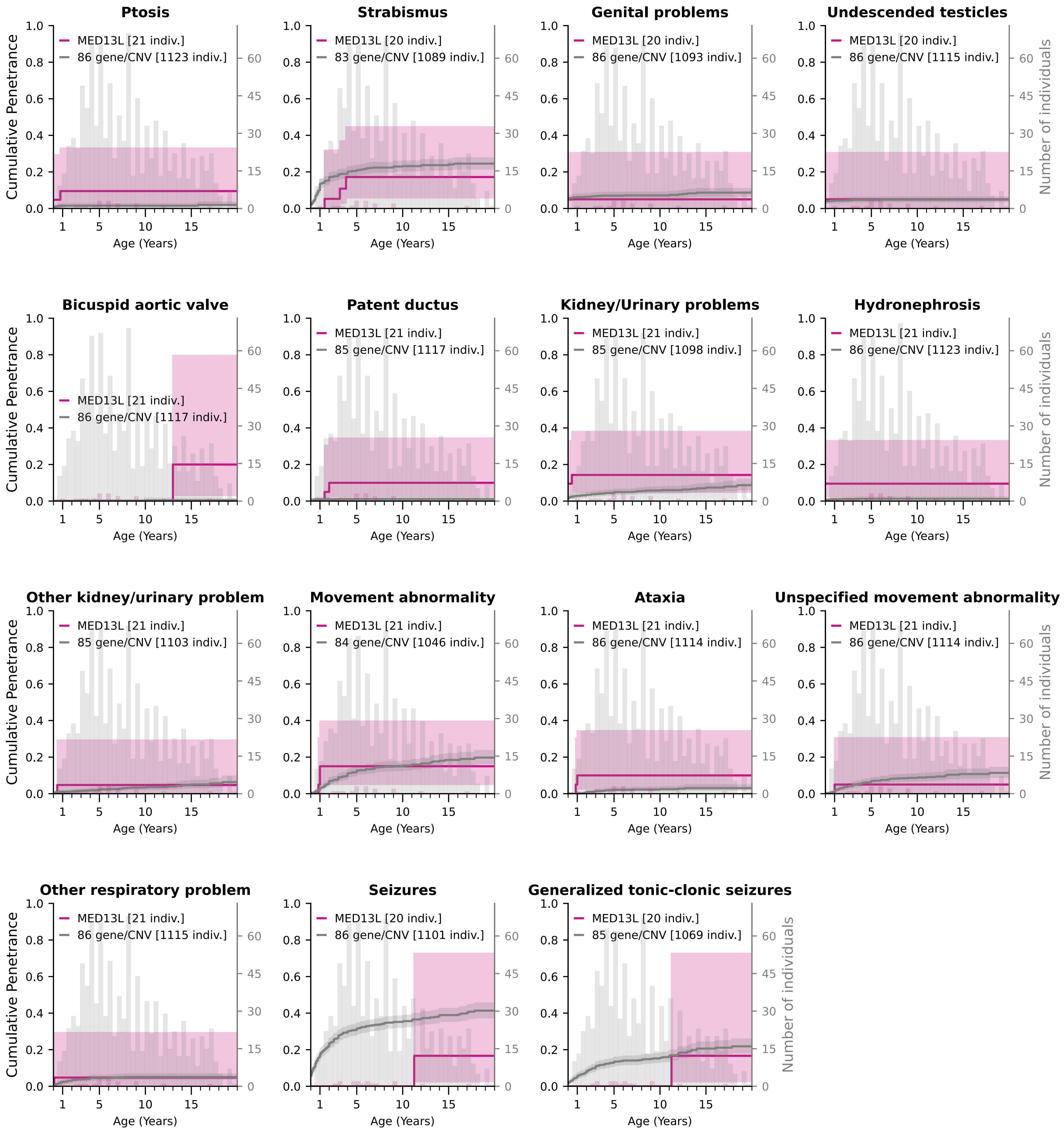

### Cumulative Penetrance for PACS1 (page 1/3)

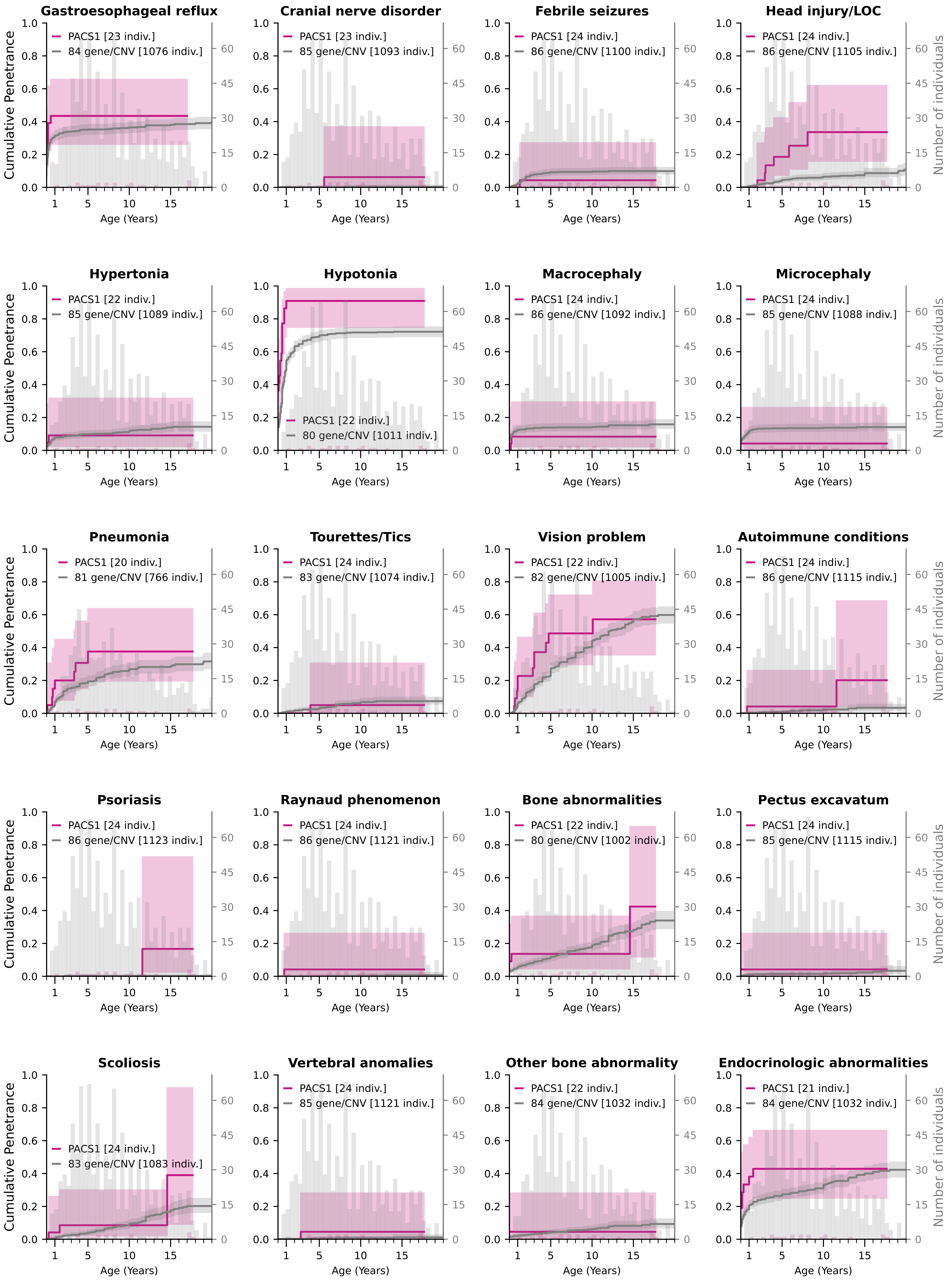

### Cumulative Penetrance for PACS1 (page 2/3)

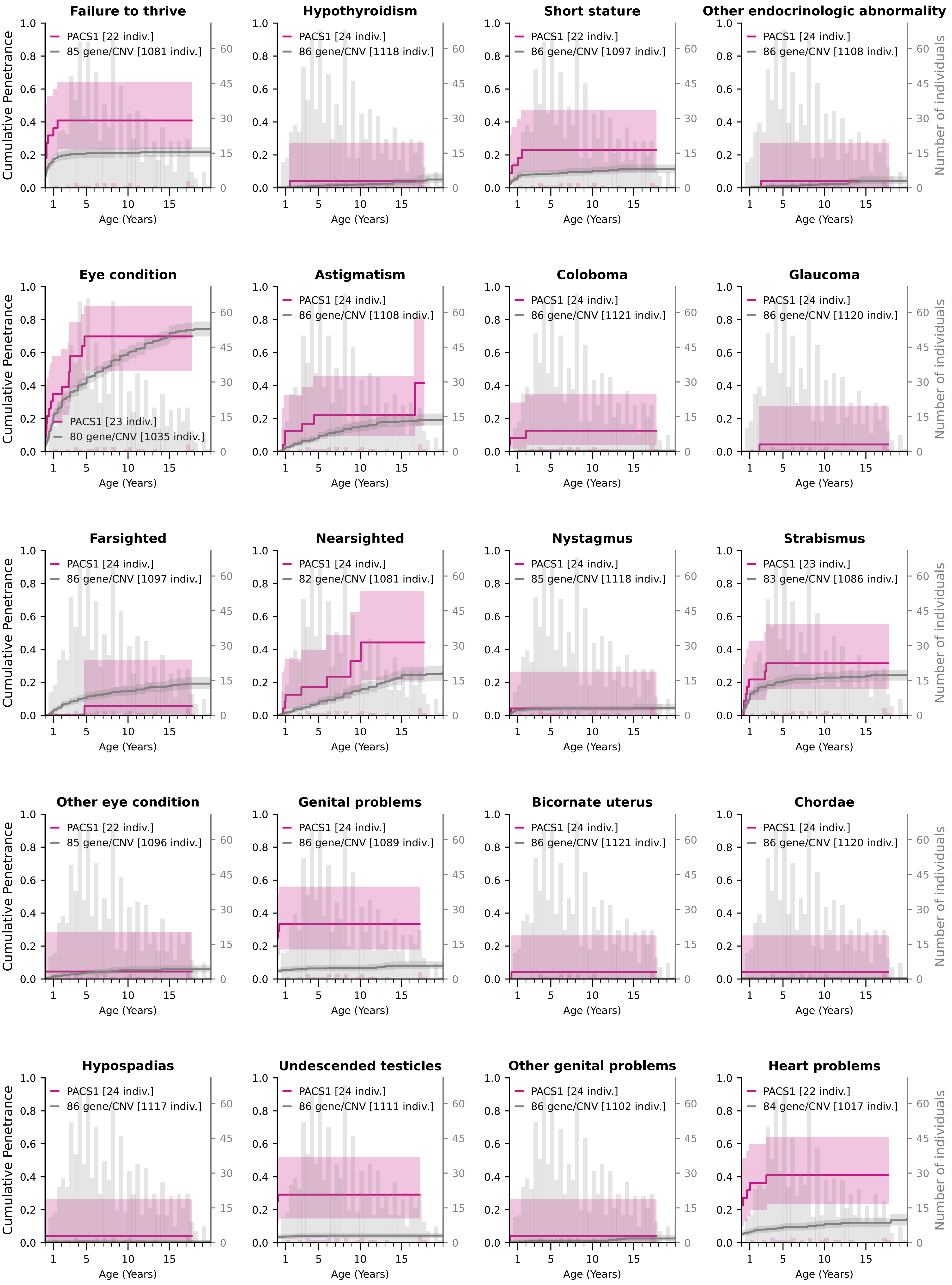

### Cumulative Penetrance for PACS1 (page 3/3)

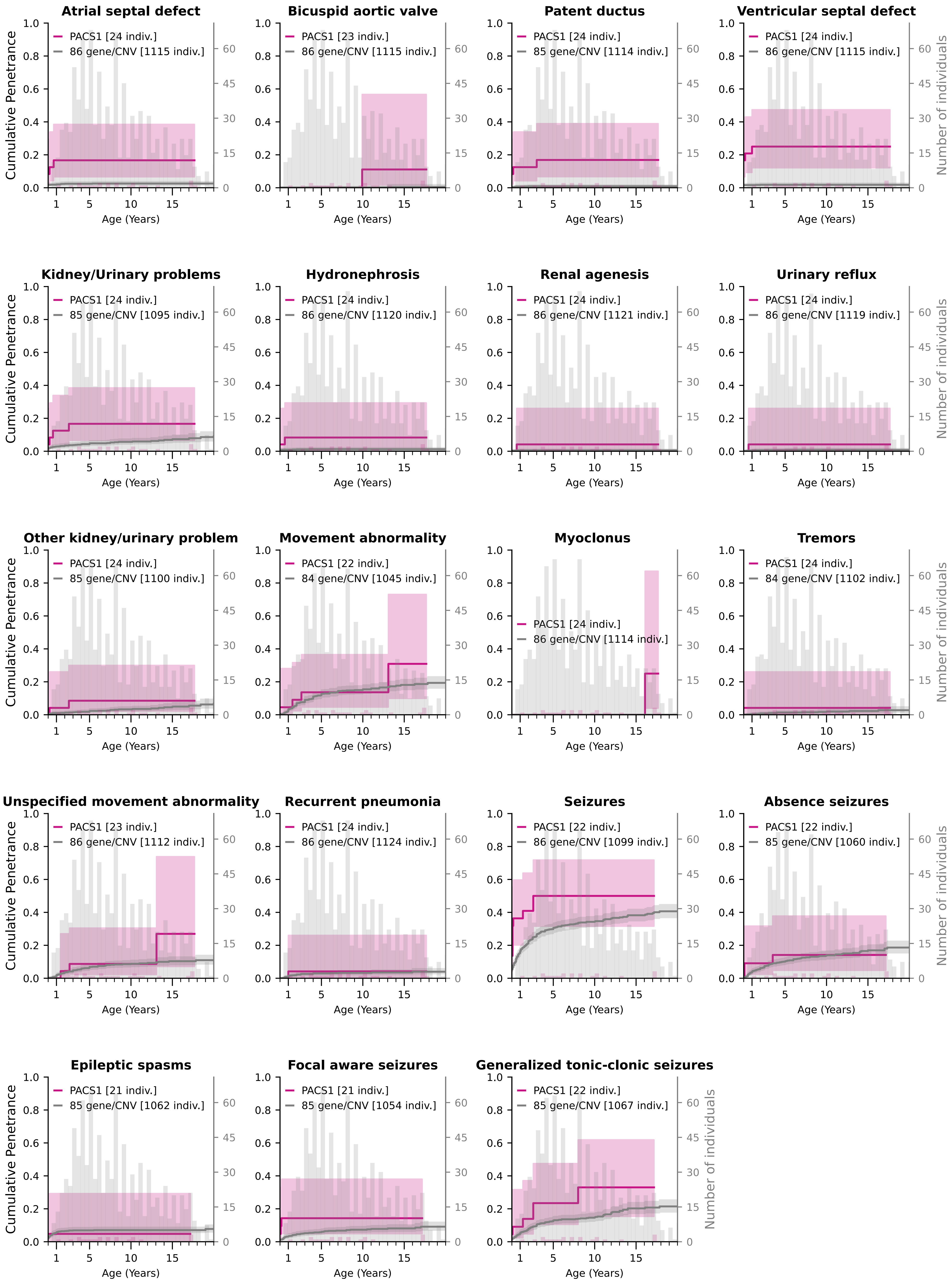

### Cumulative Penetrance for PPP2R5D (page 1/4)

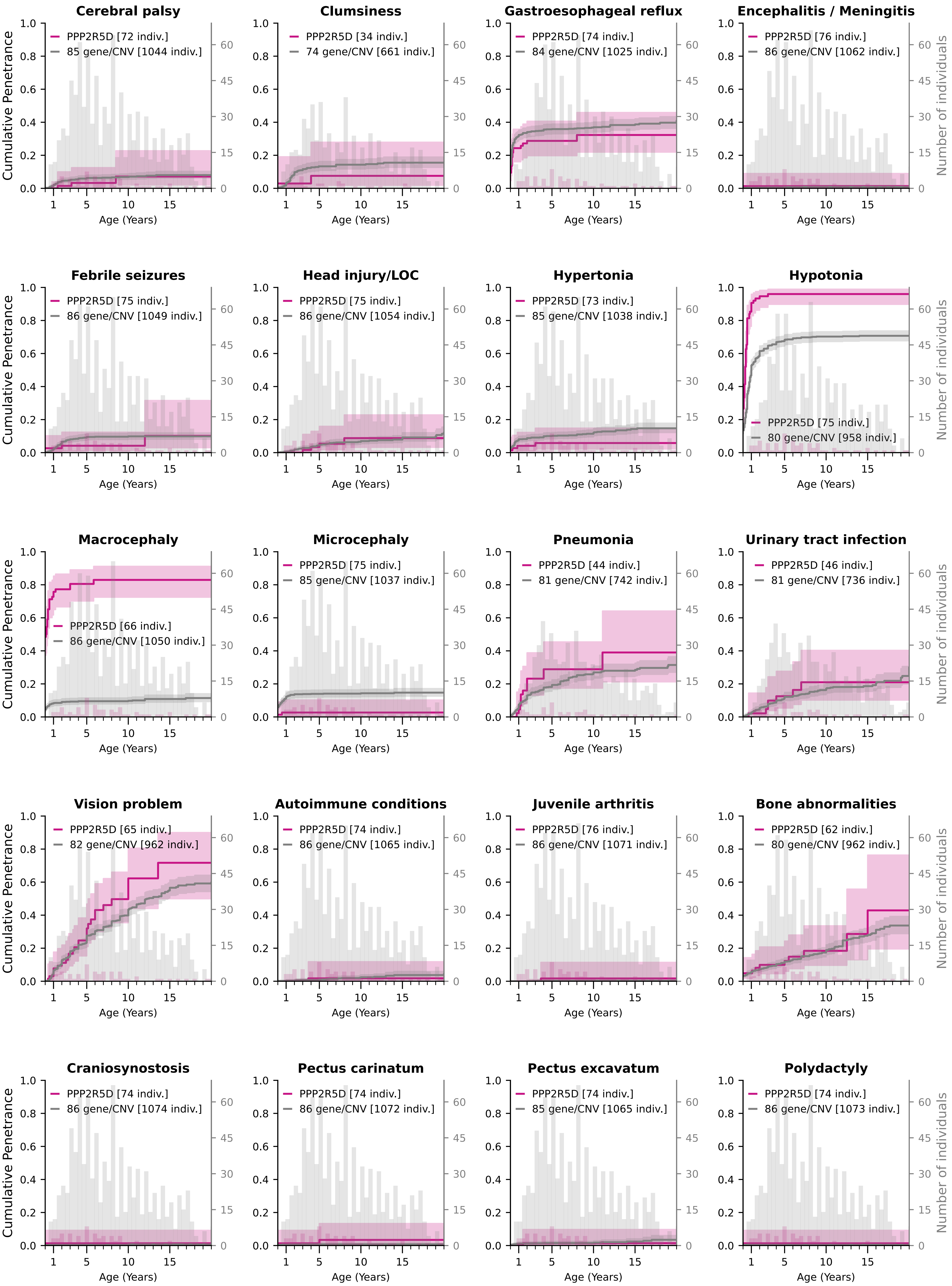

### Cumulative Penetrance for PPP2R5D (page 2/4)

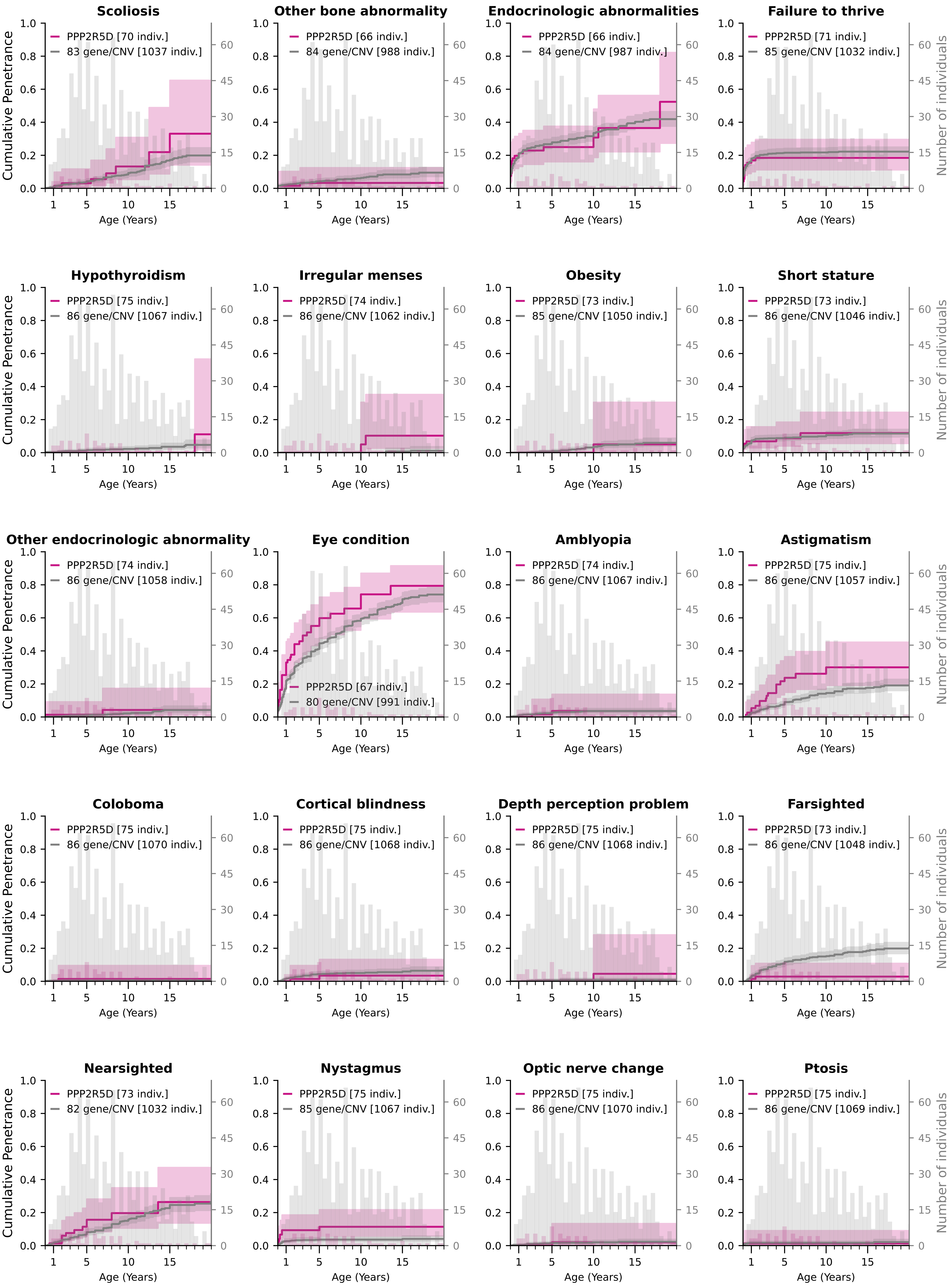

### Cumulative Penetrance for PPP2R5D (page 3/4)

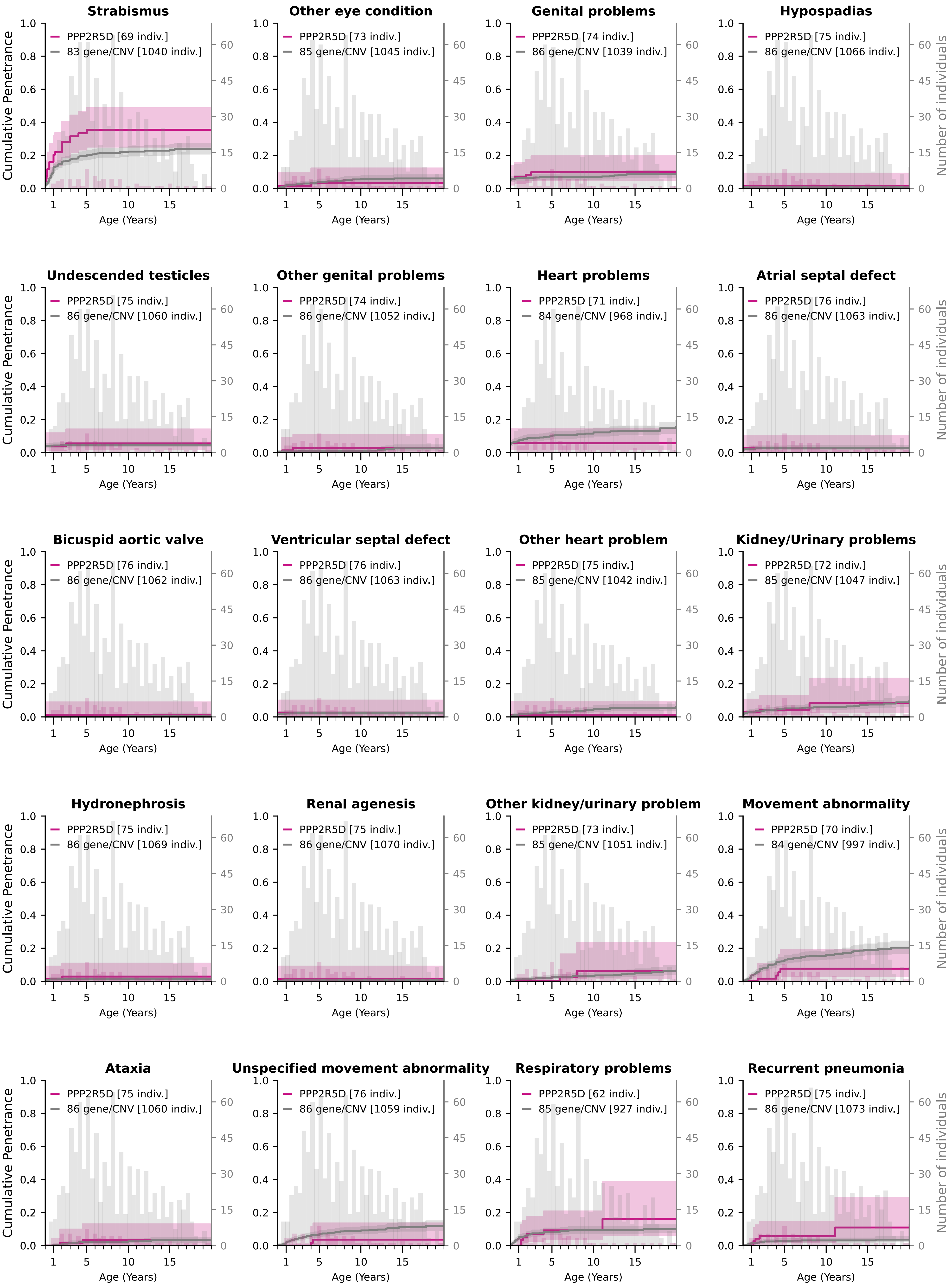

### Cumulative Penetrance for PPP2R5D (page 4/4)

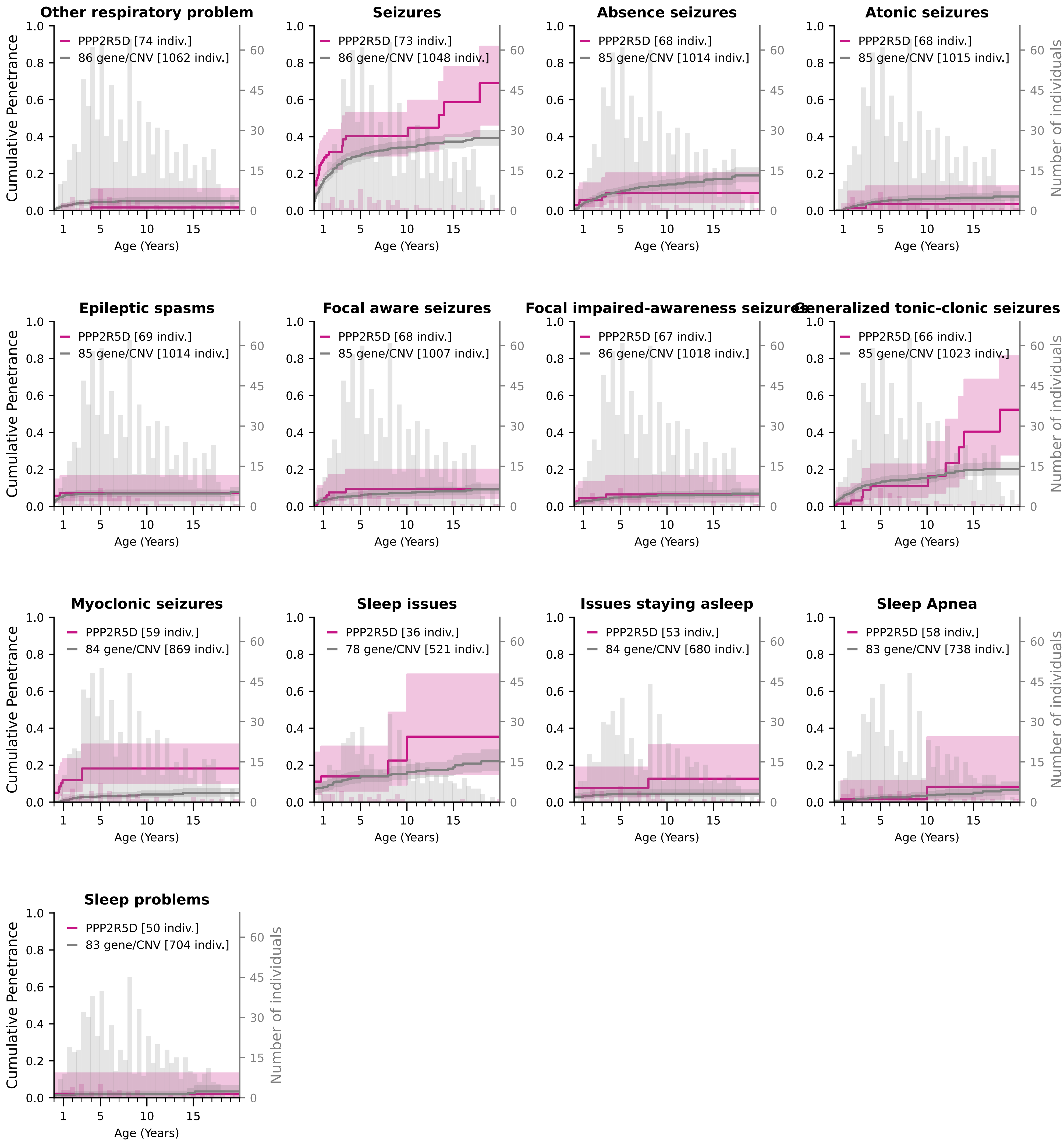

### Cumulative Penetrance for SCN2A (page 1/5)

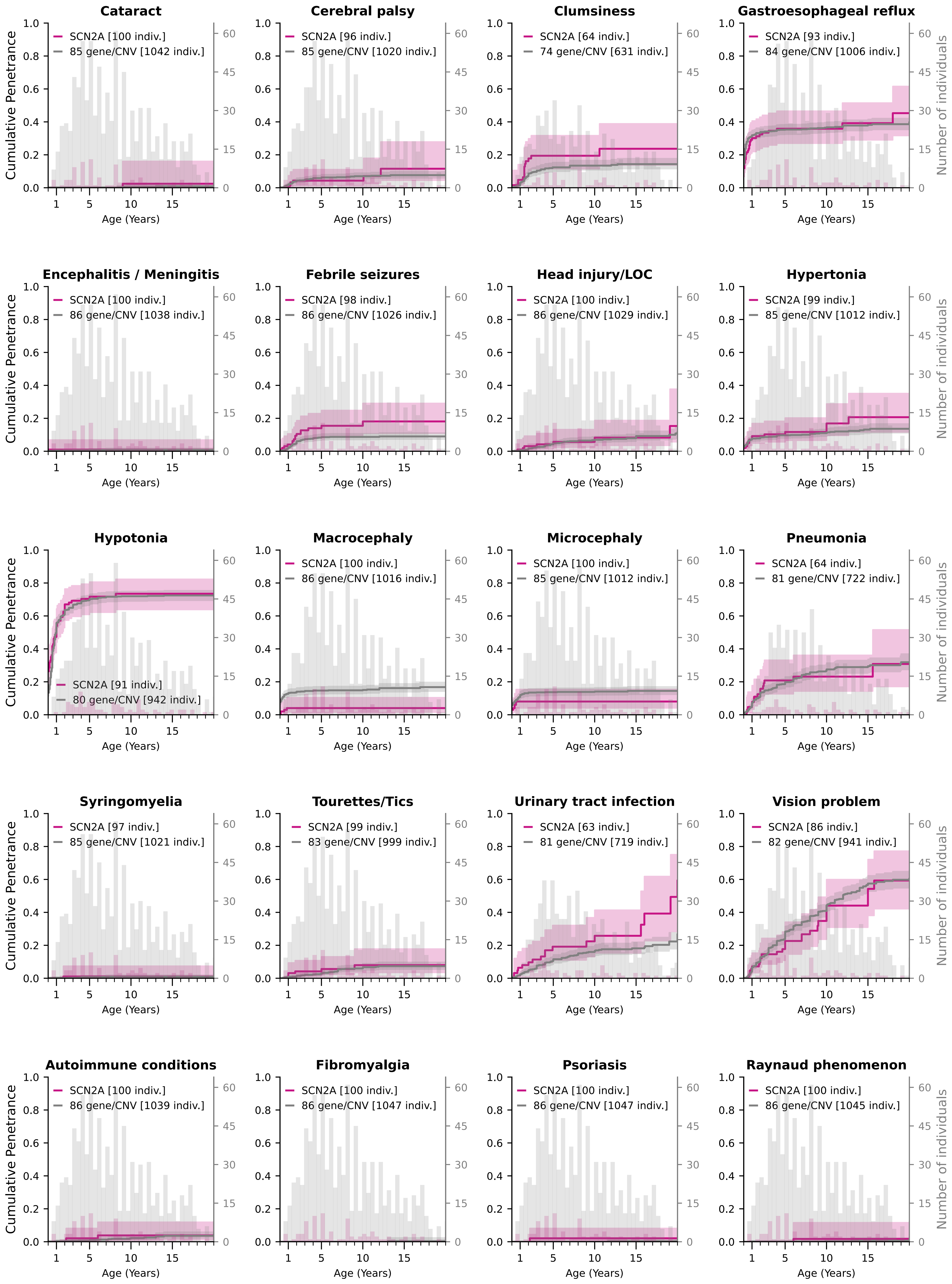

### Cumulative Penetrance for SCN2A (page 2/5)

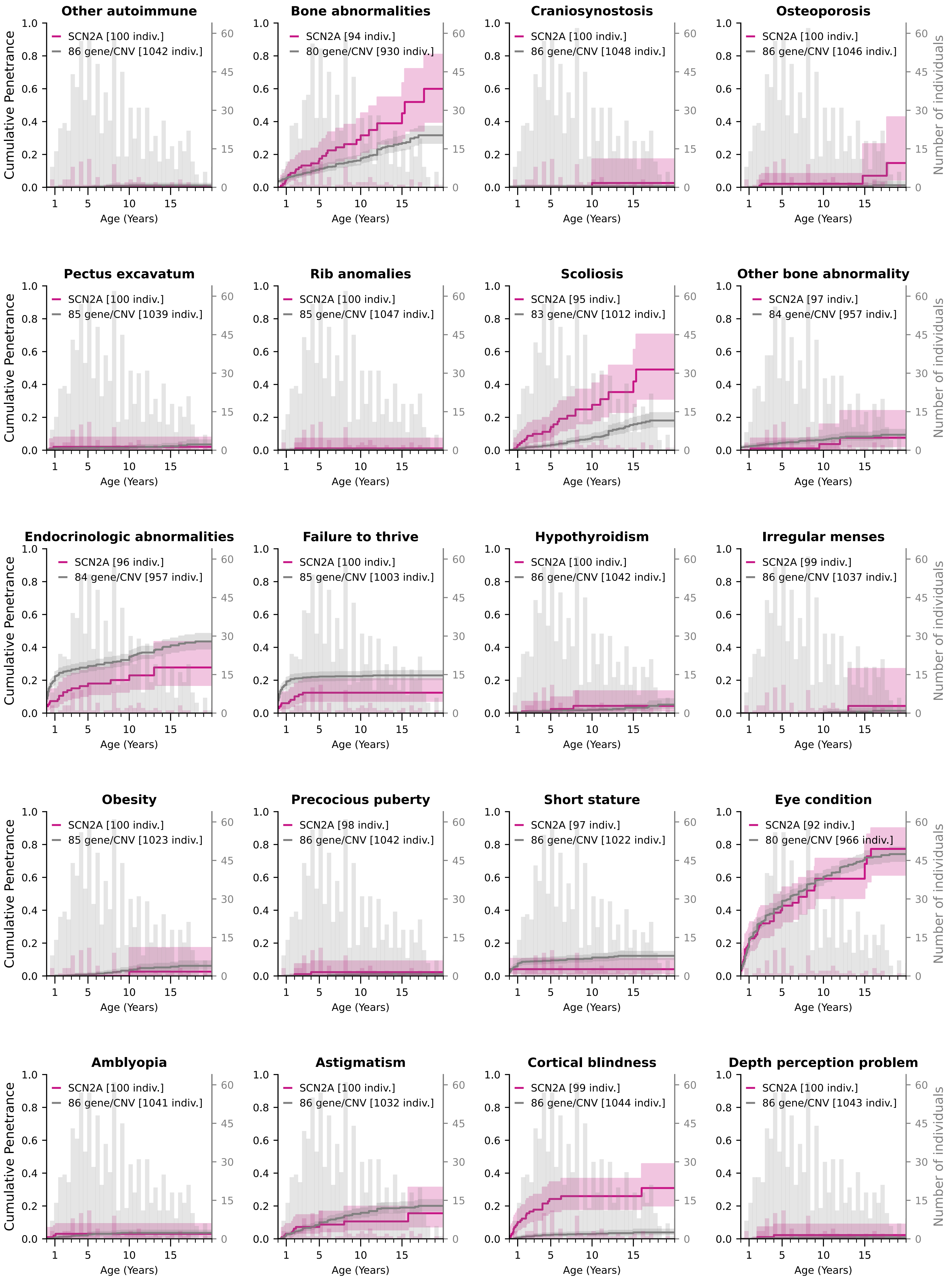

### Cumulative Penetrance for SCN2A (page 3/5)

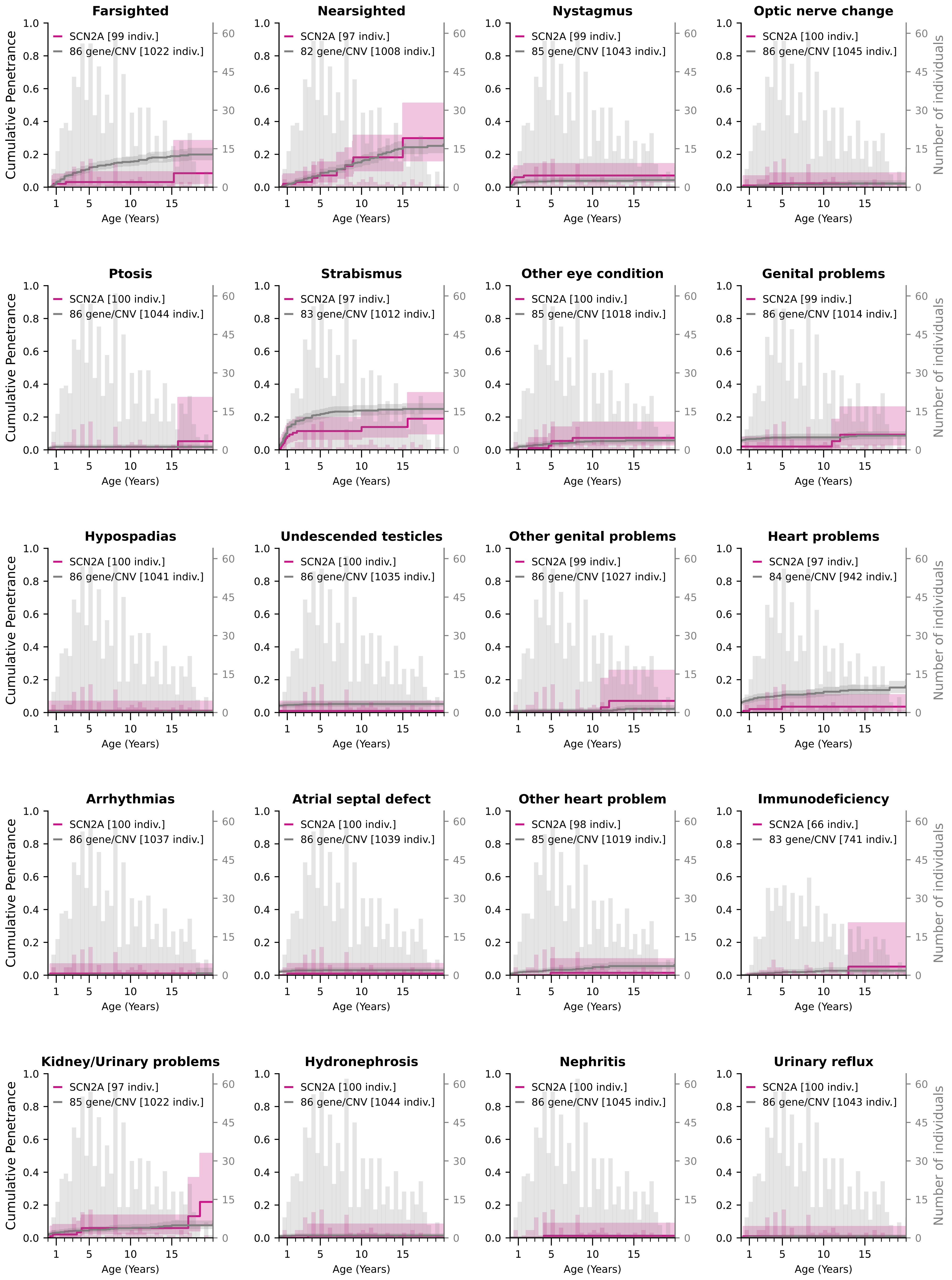

### Cumulative Penetrance for SCN2A (page 4/5)

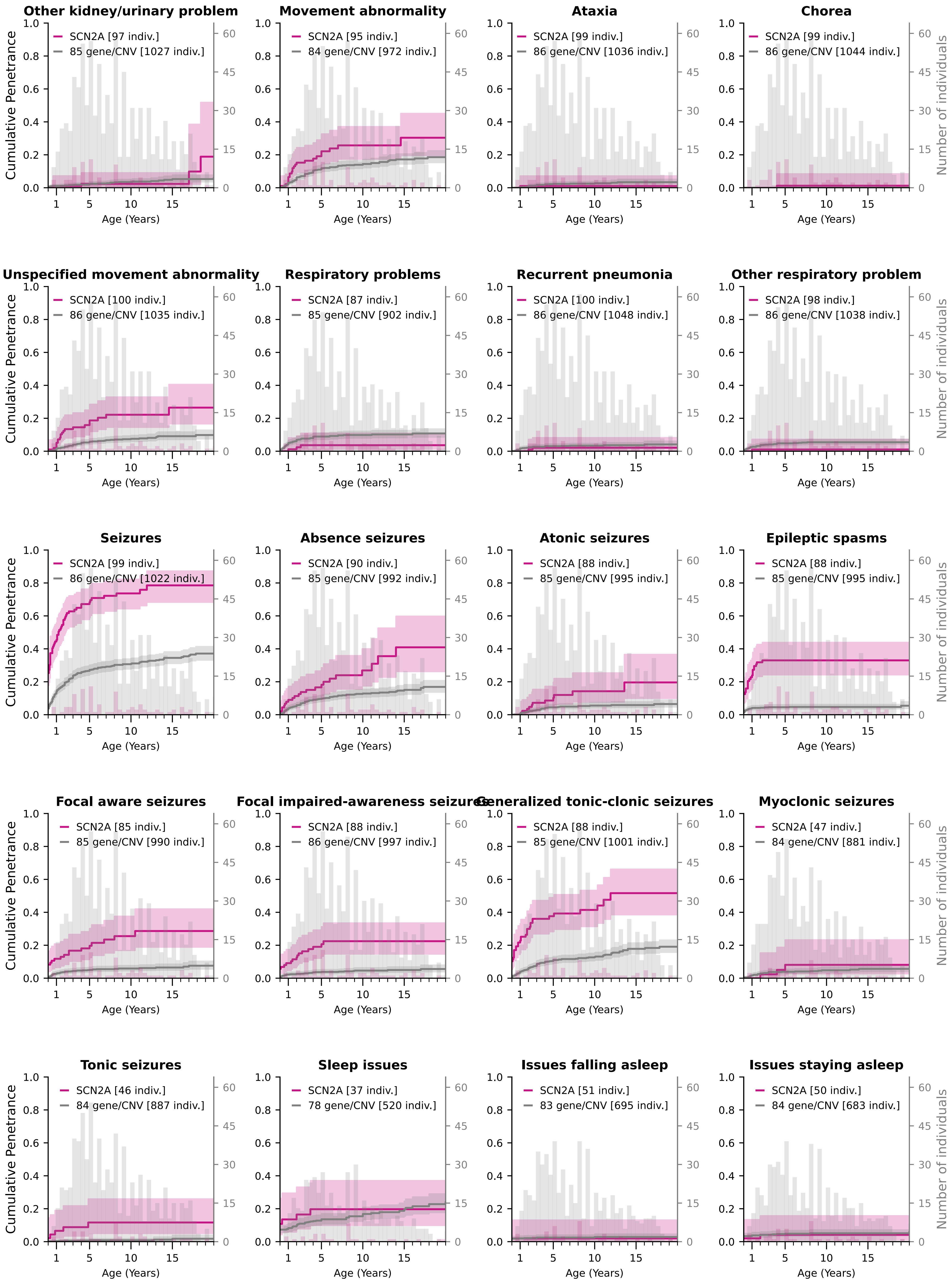

### Cumulative Penetrance for SCN2A (page 5/5)

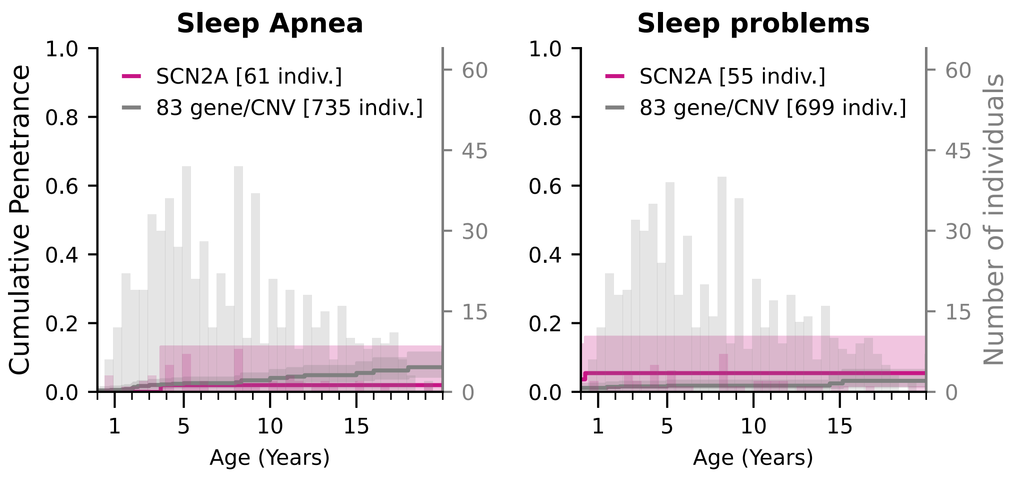

### Cumulative Penetrance for SETBP1

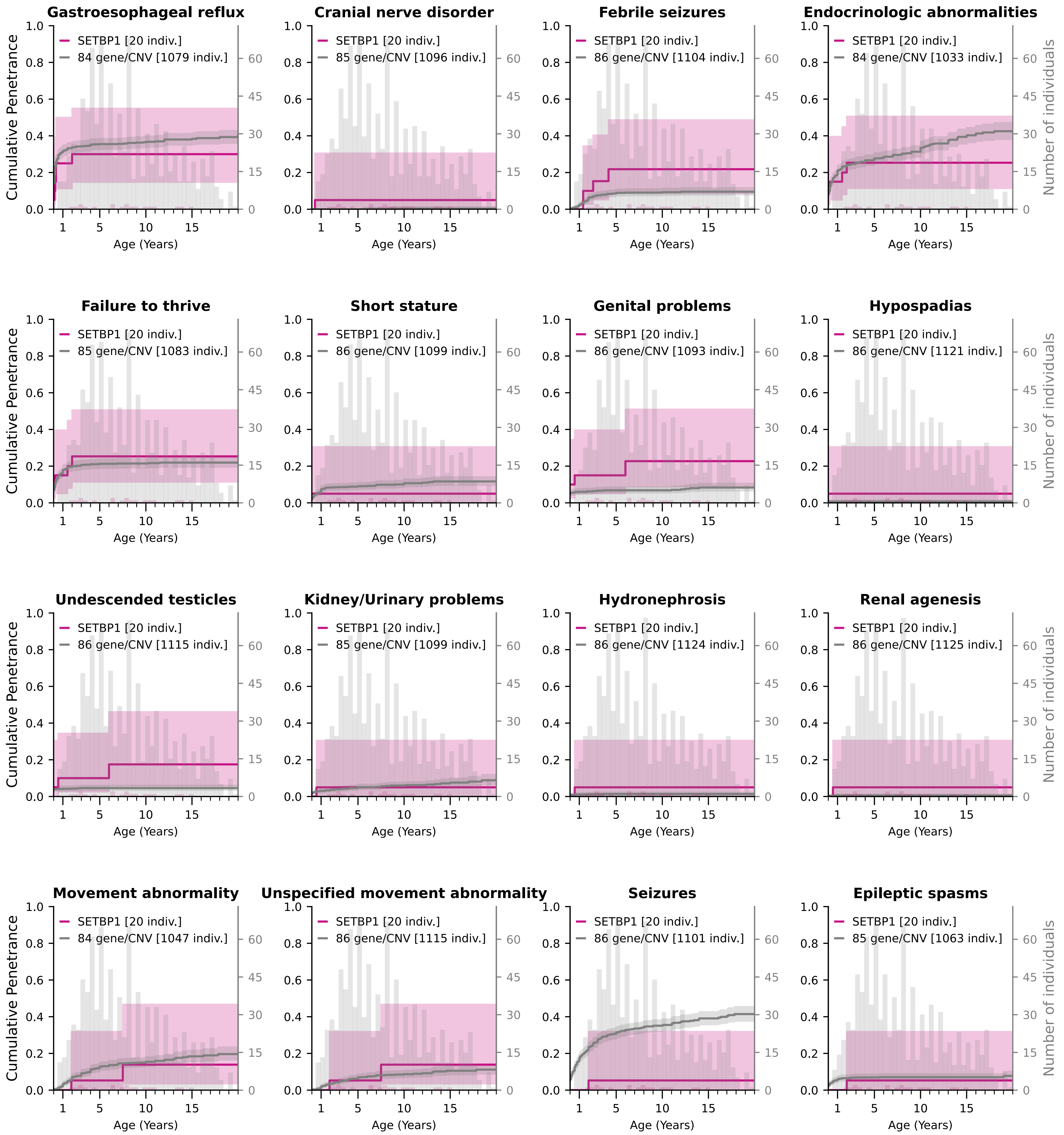

### Cumulative Penetrance for SLC6A1 (page 1/3)

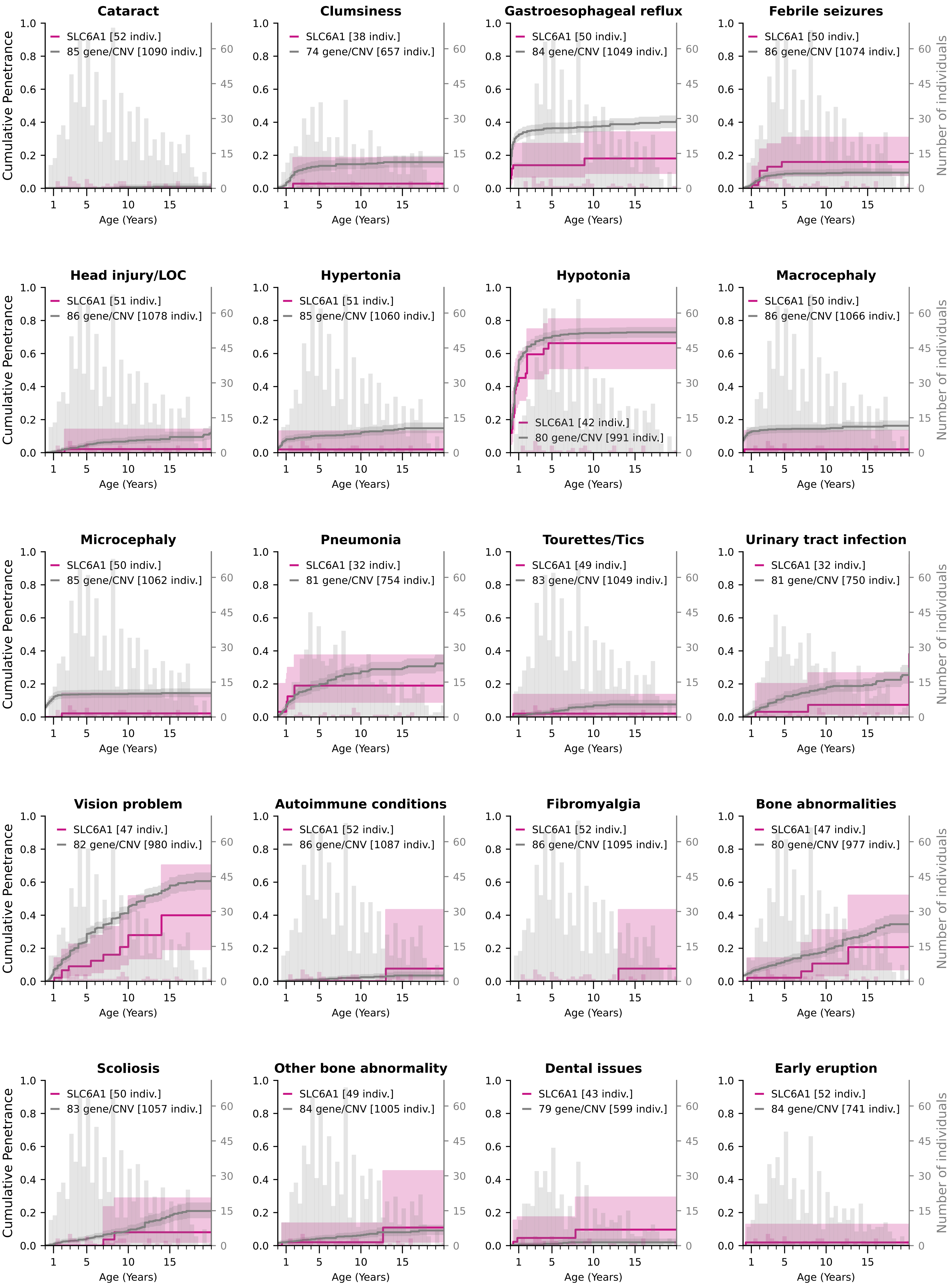

### Cumulative Penetrance for SLC6A1 (page 2/3)

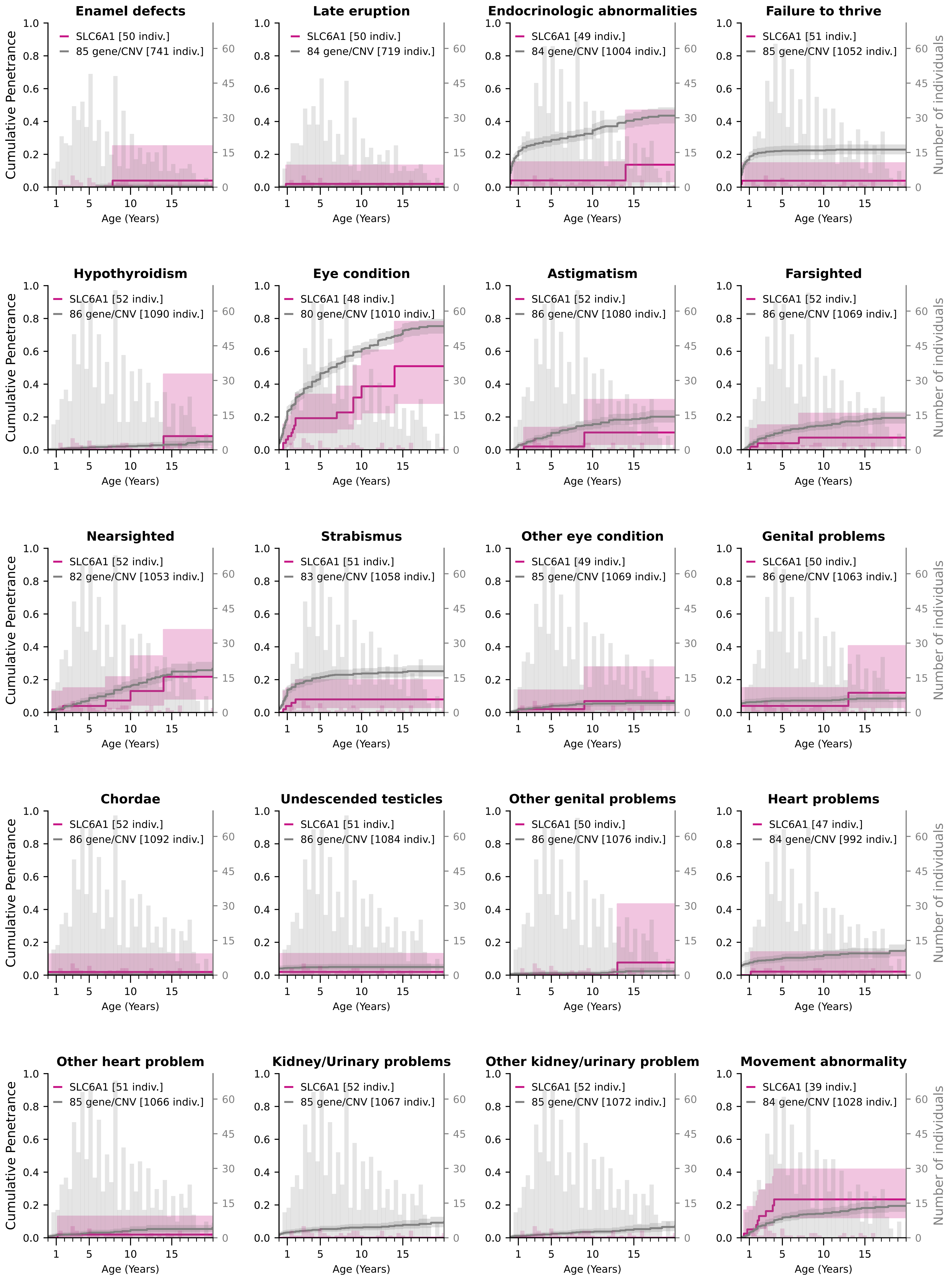

### Cumulative Penetrance for SLC6A1 (page 3/3)

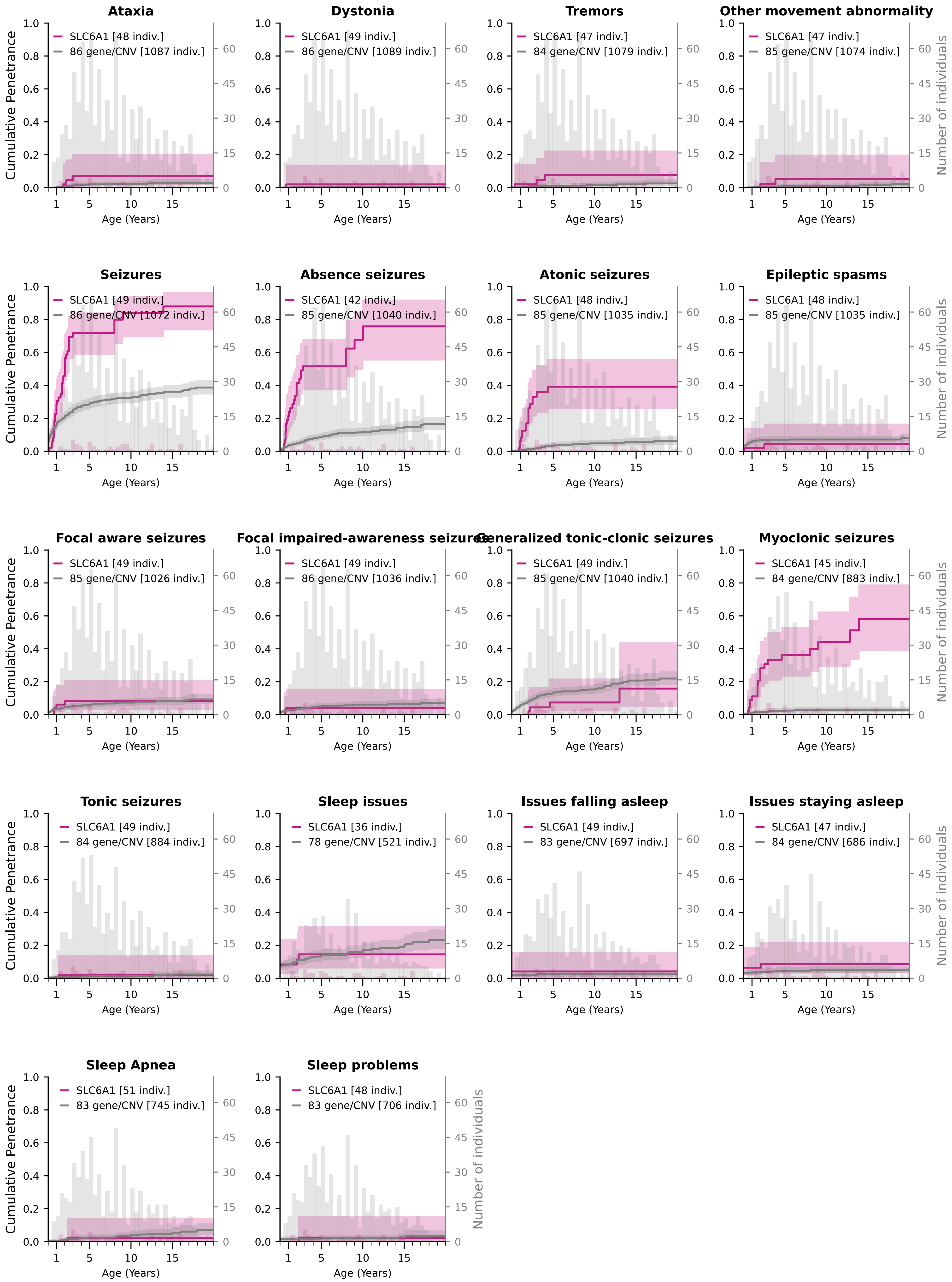

### Cumulative Penetrance for STXBP1 (page 1/4)

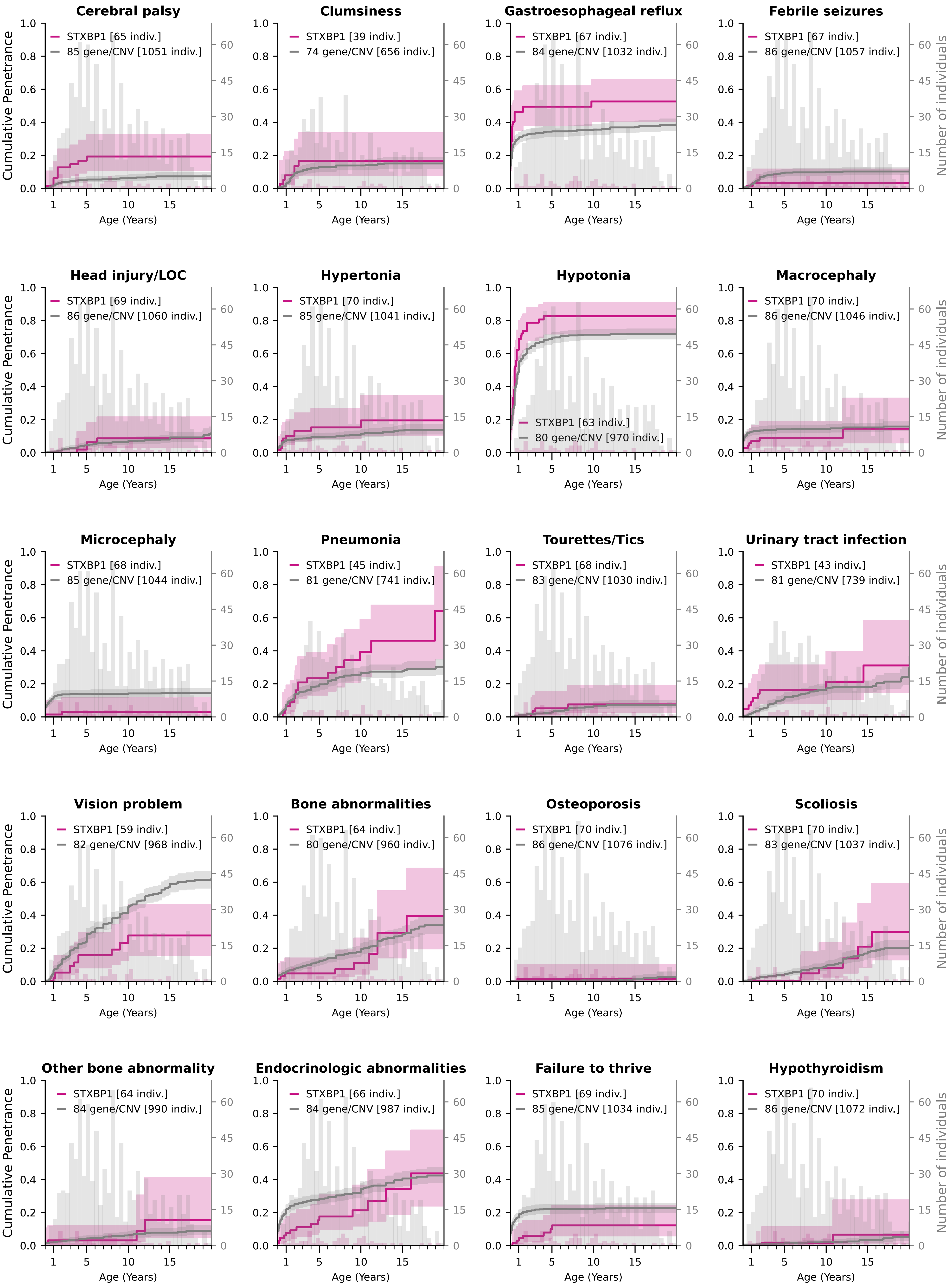

### Cumulative Penetrance for STXBP1 (page 2/4)

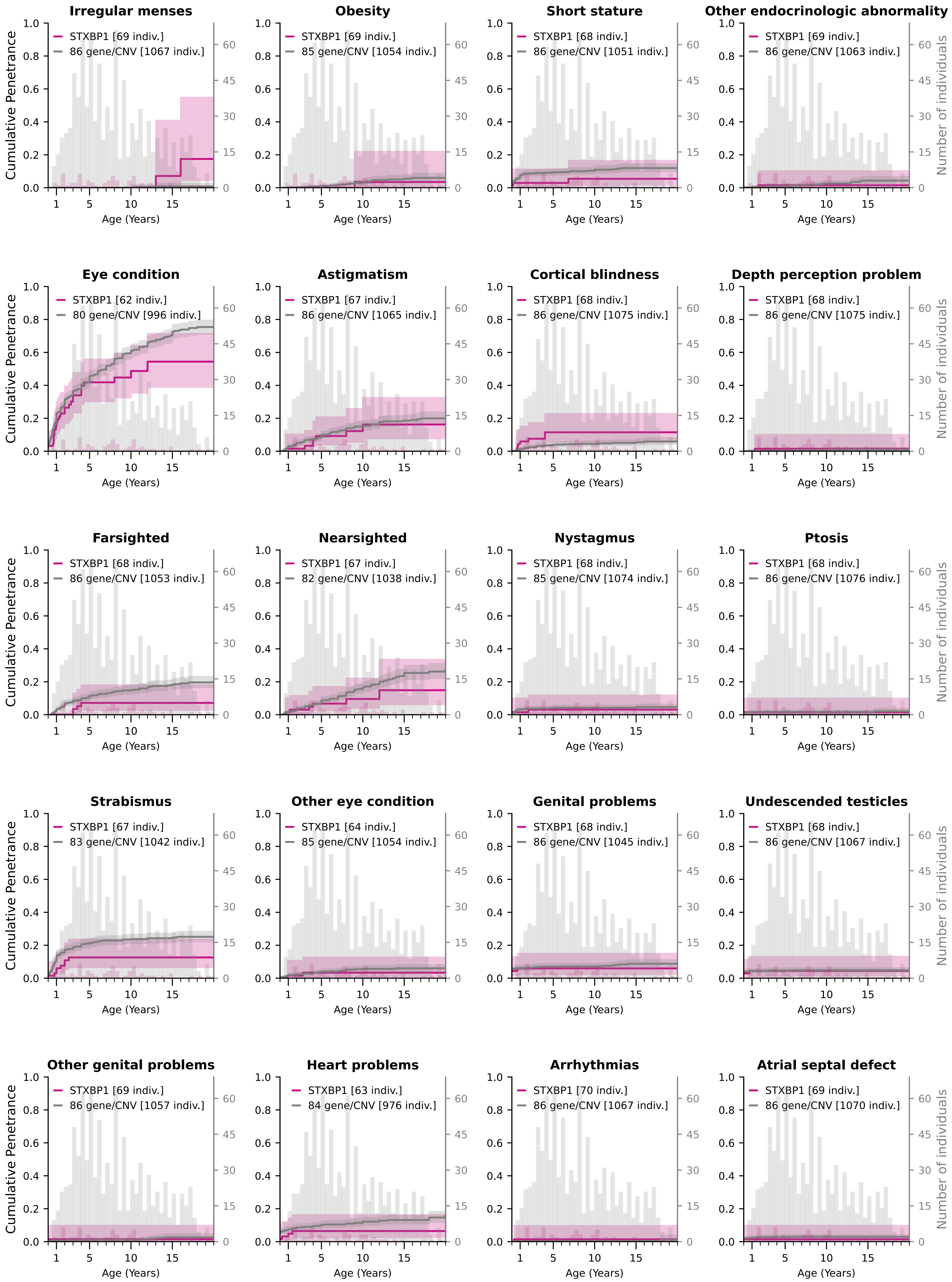

### Cumulative Penetrance for STXBP1 (page 3/4)

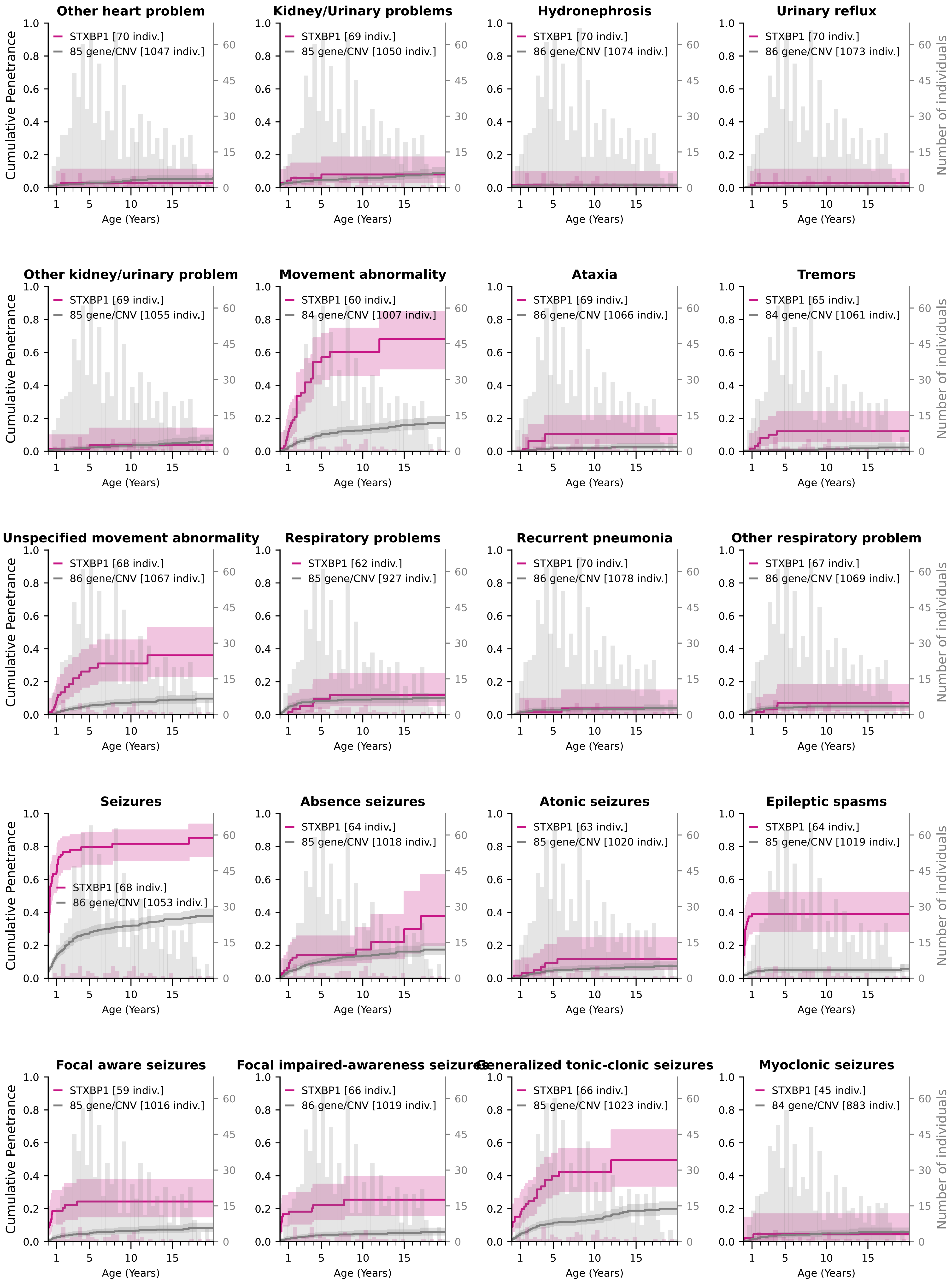

### Cumulative Penetrance for STXBP1 (page 4/4)

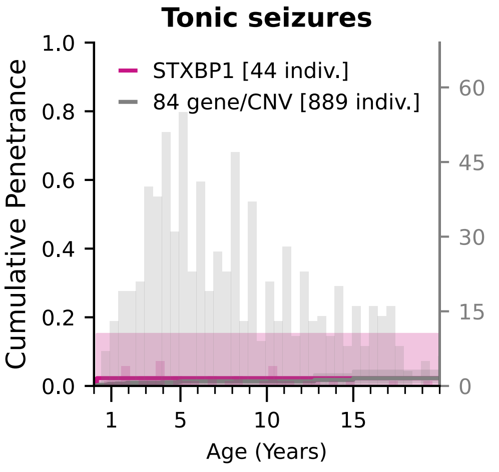


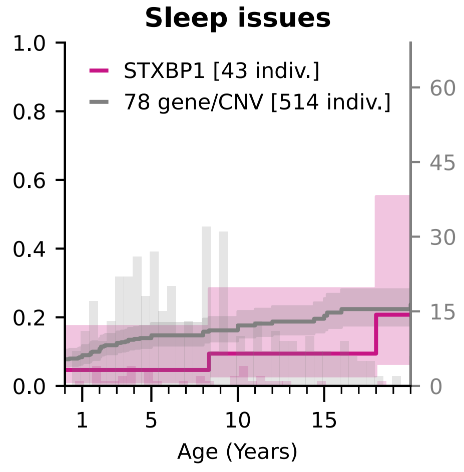


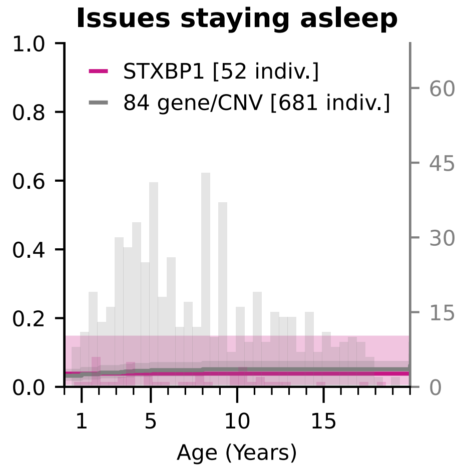


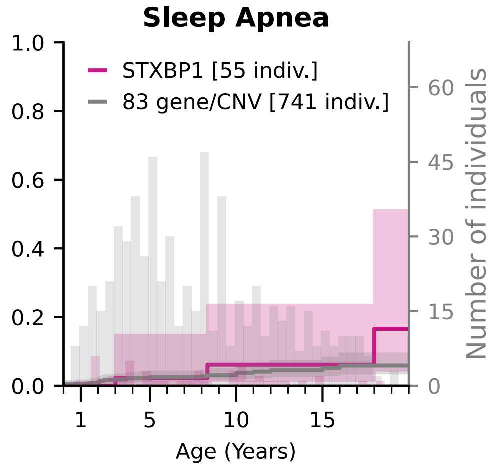


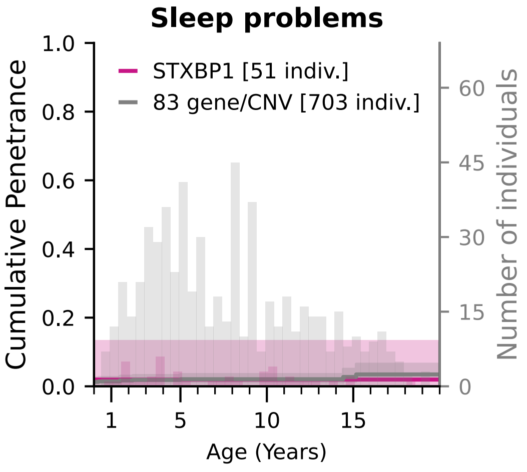

### Cumulative Penetrance for SYNGAP1 (page 1/3)

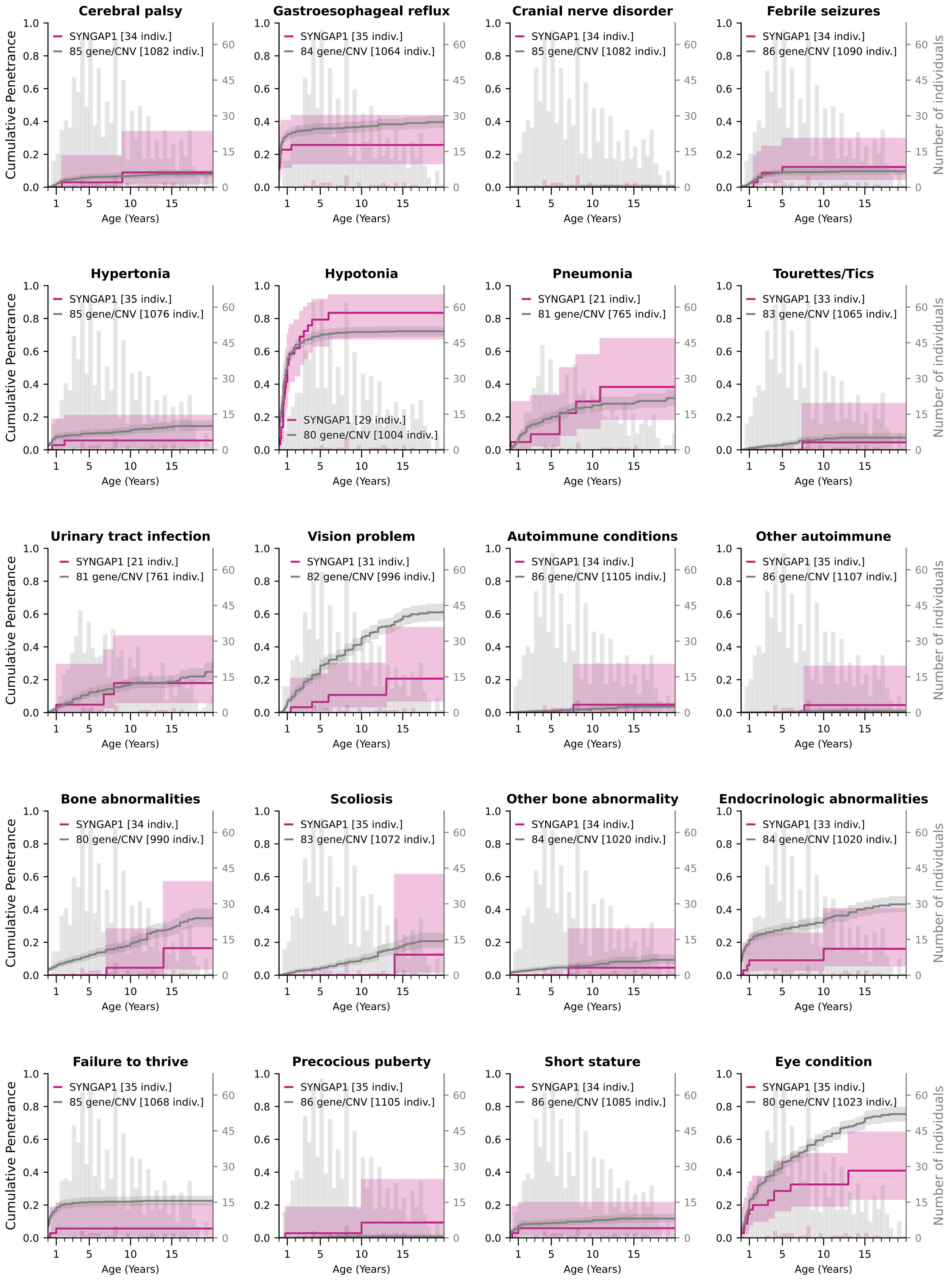

### Cumulative Penetrance for SYNGAP1 (page 2/3)

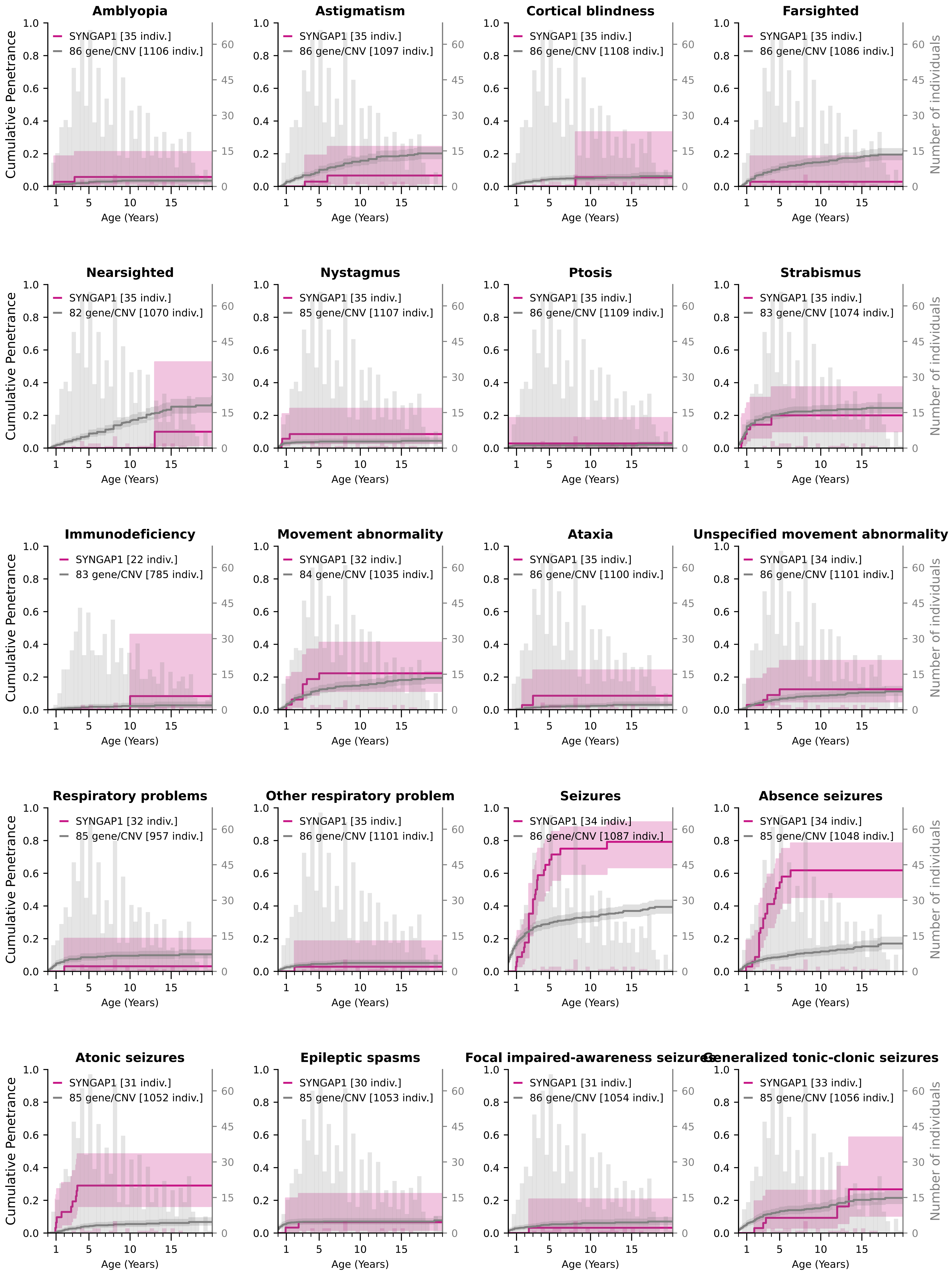

### Cumulative Penetrance for SYNGAP1 (page 3/3)

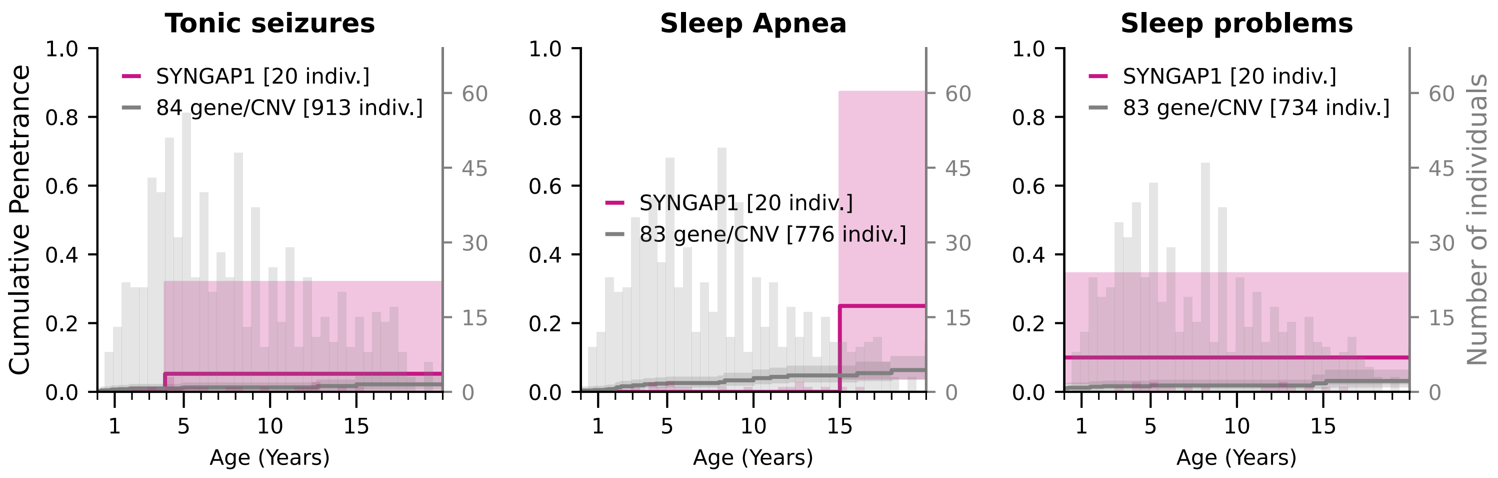

### Cumulative Penetrance for 1q21.1 deletion (page 1/4)

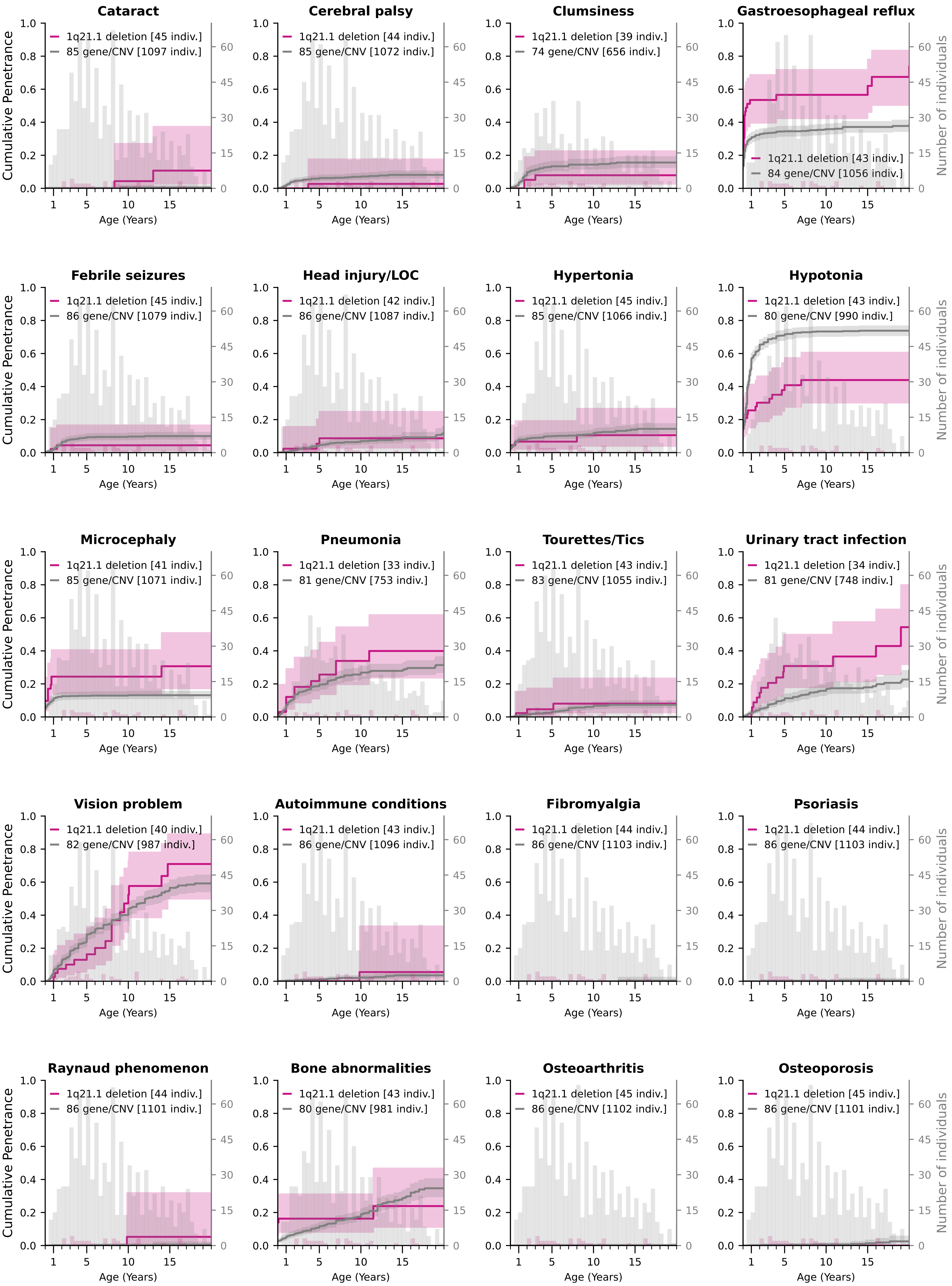

### Cumulative Penetrance for 1q21.1 deletion (page 2/4)

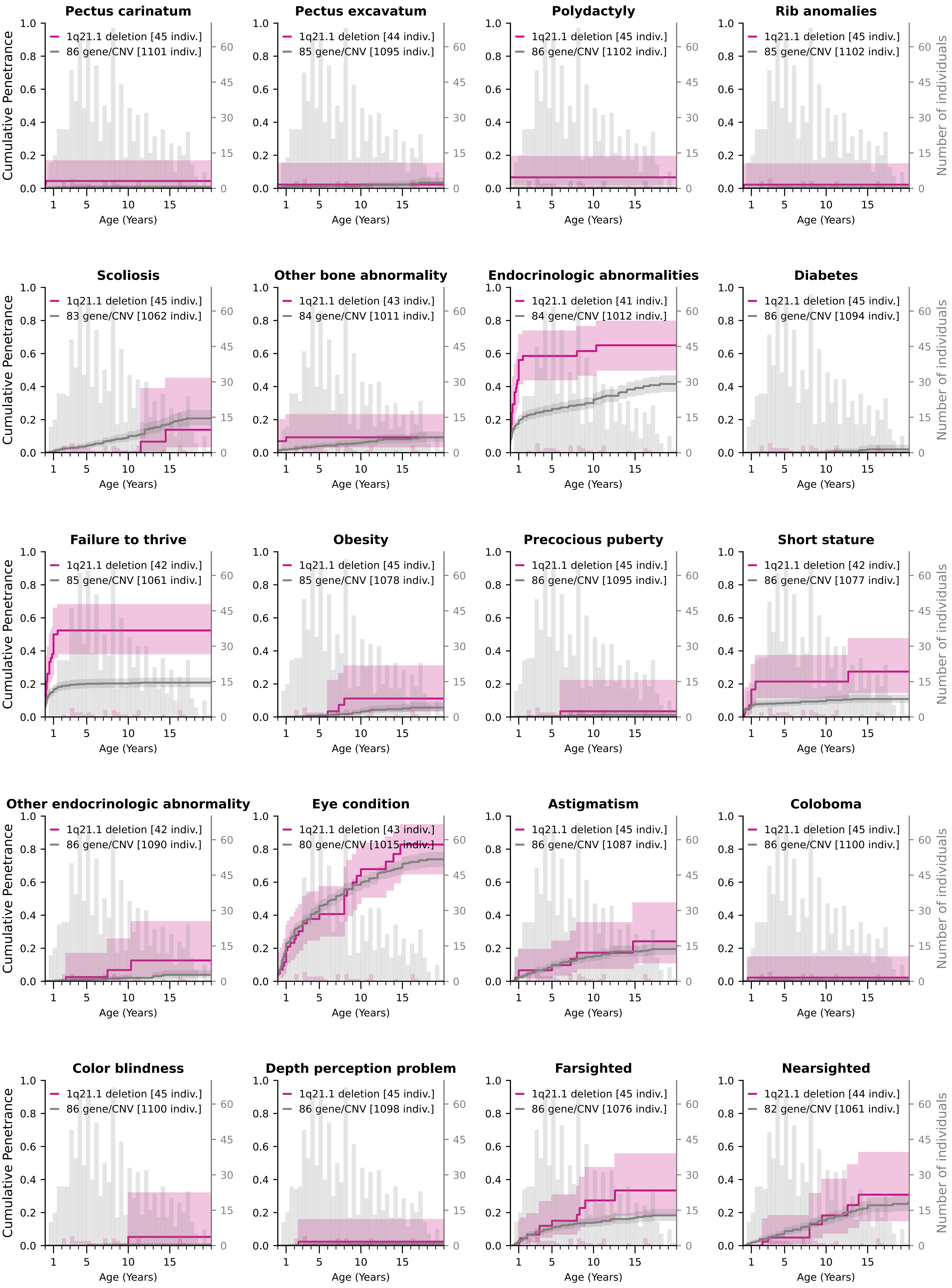

### Cumulative Penetrance for 1q21.1 deletion (page 3/4)

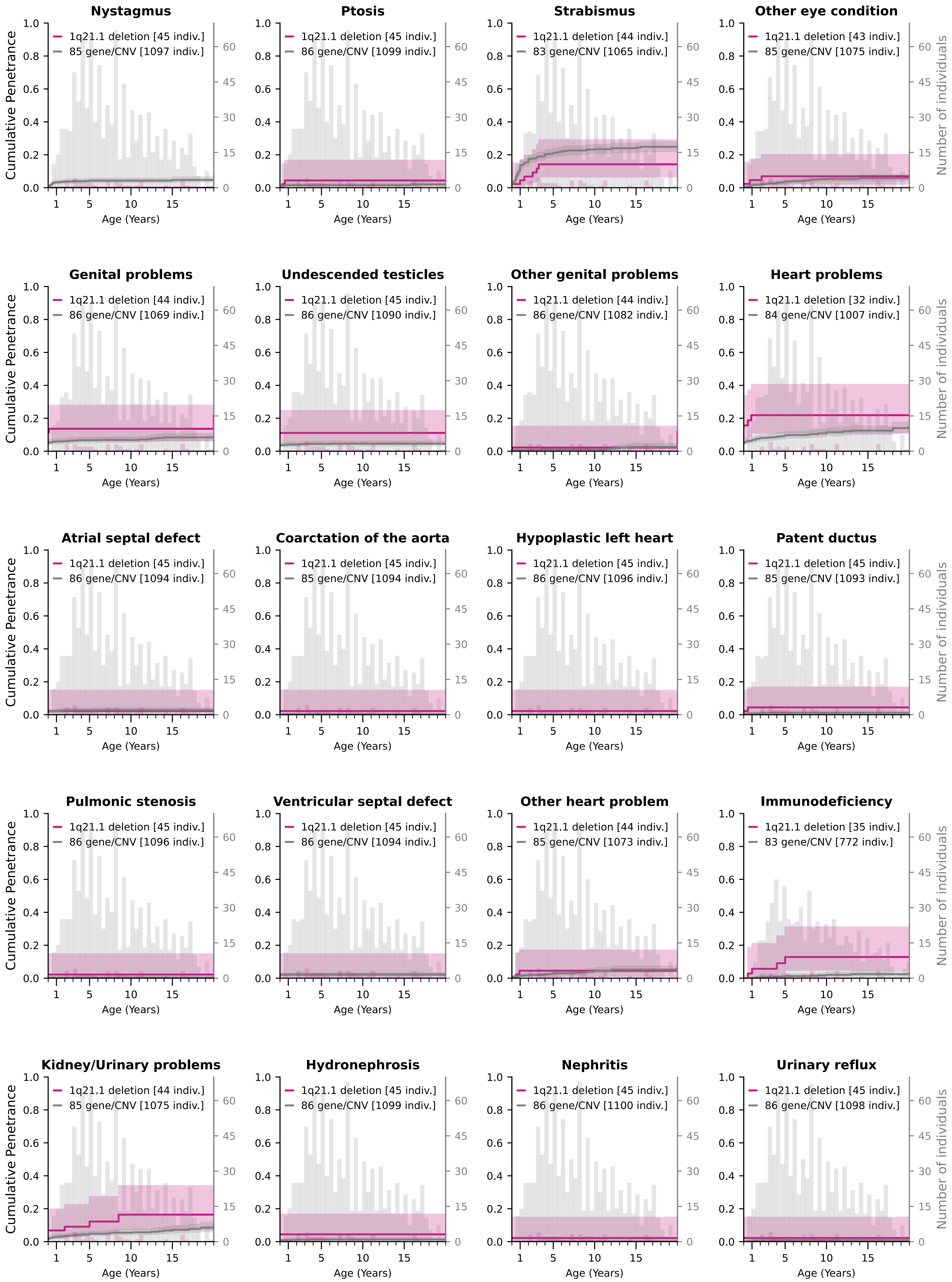

### Cumulative Penetrance for 1q21.1 deletion (page 4/4)

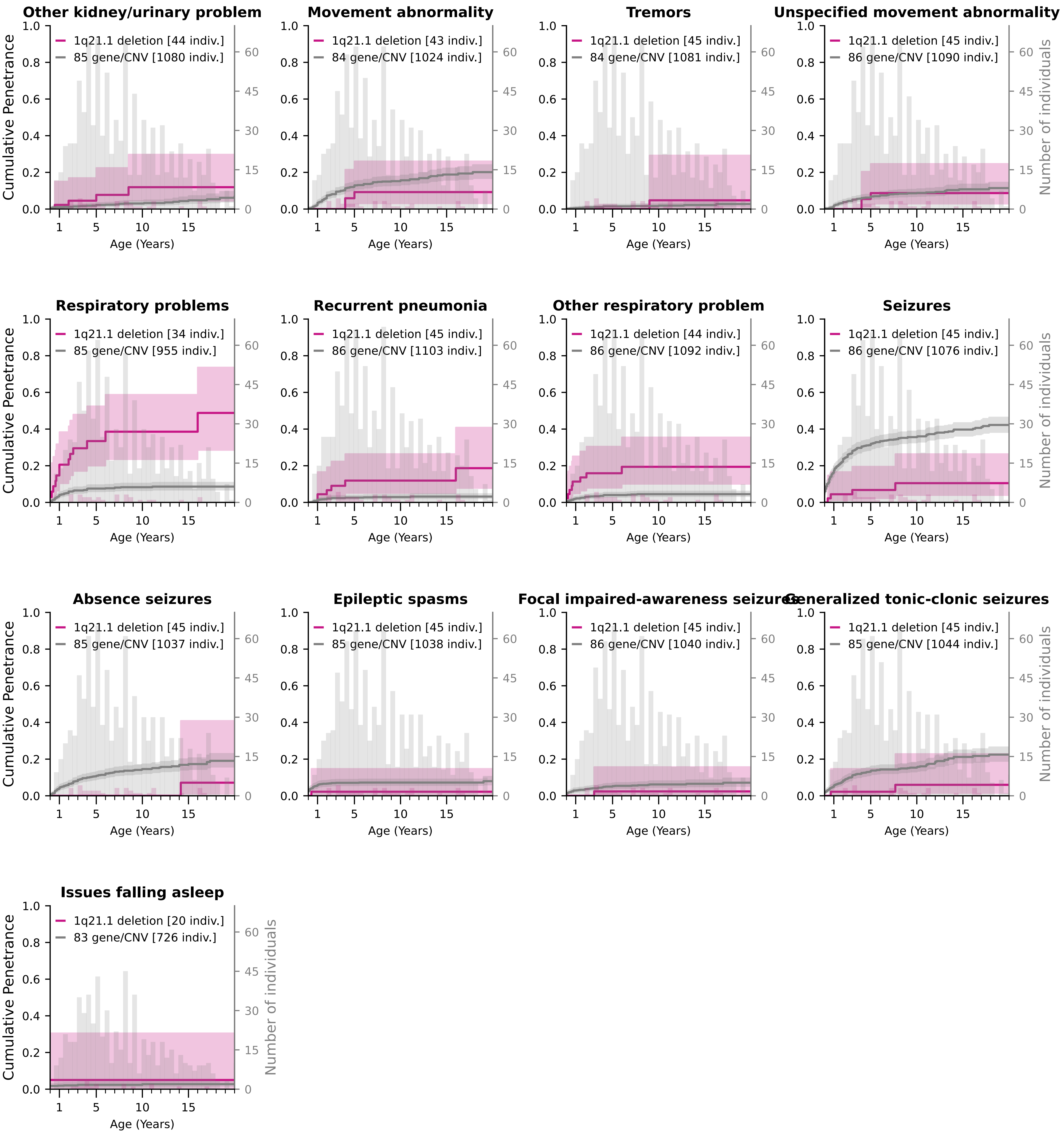

### Cumulative Penetrance for 1q21.1 duplication (page 1/4)

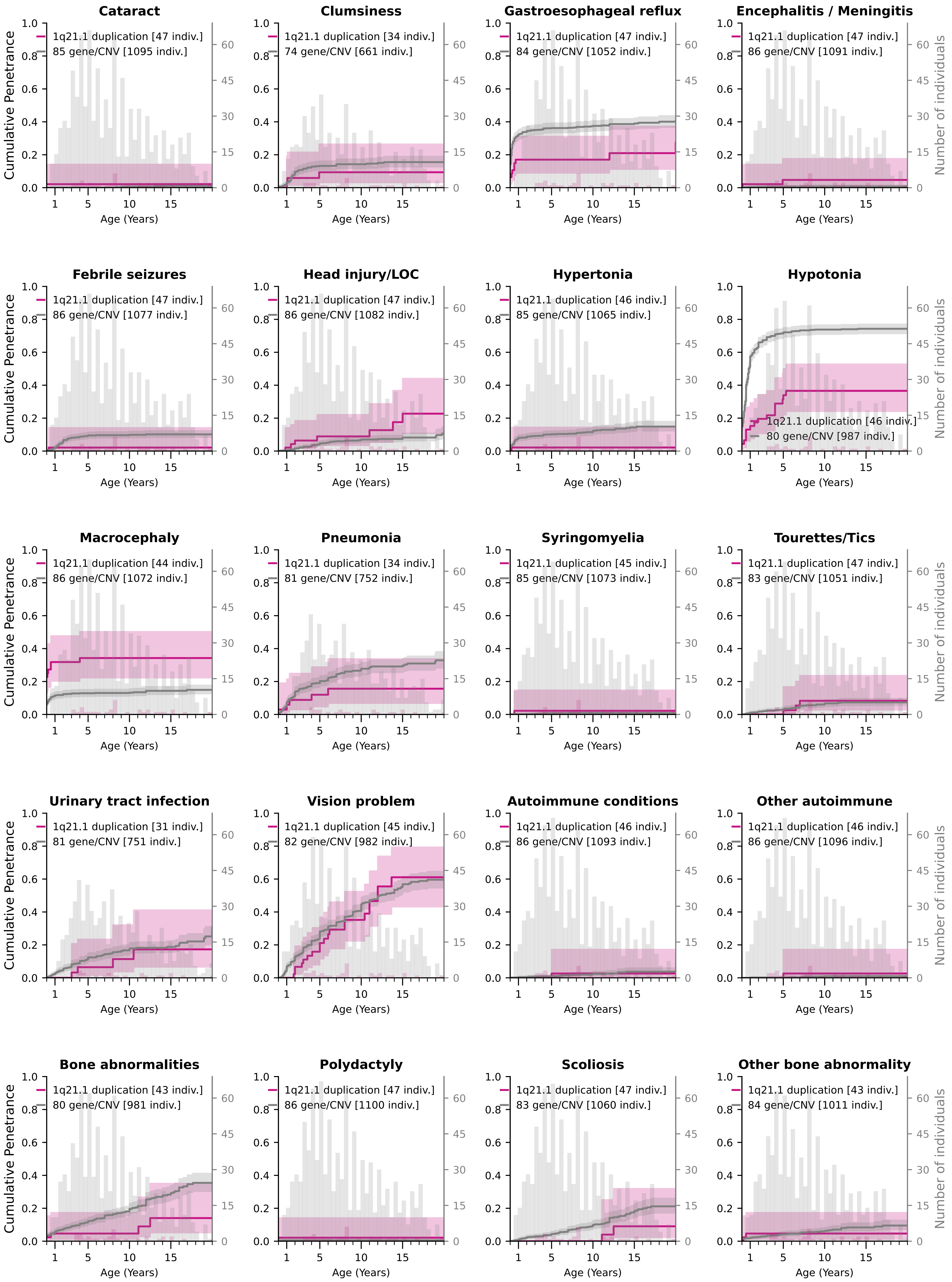

### Cumulative Penetrance for 1q21.1 duplication (page 2/4)

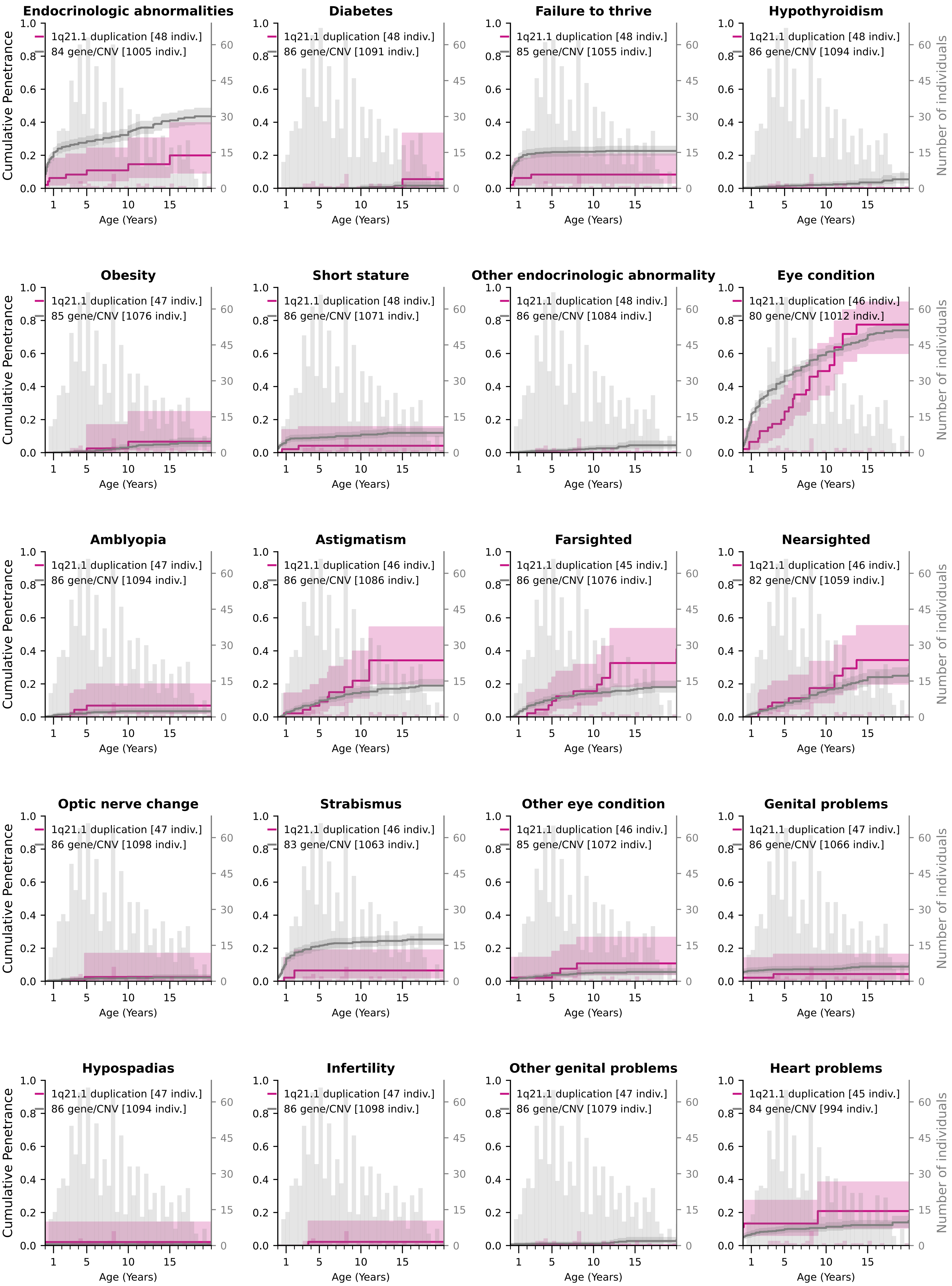

### Cumulative Penetrance for 1q21.1 duplication (page 3/4)

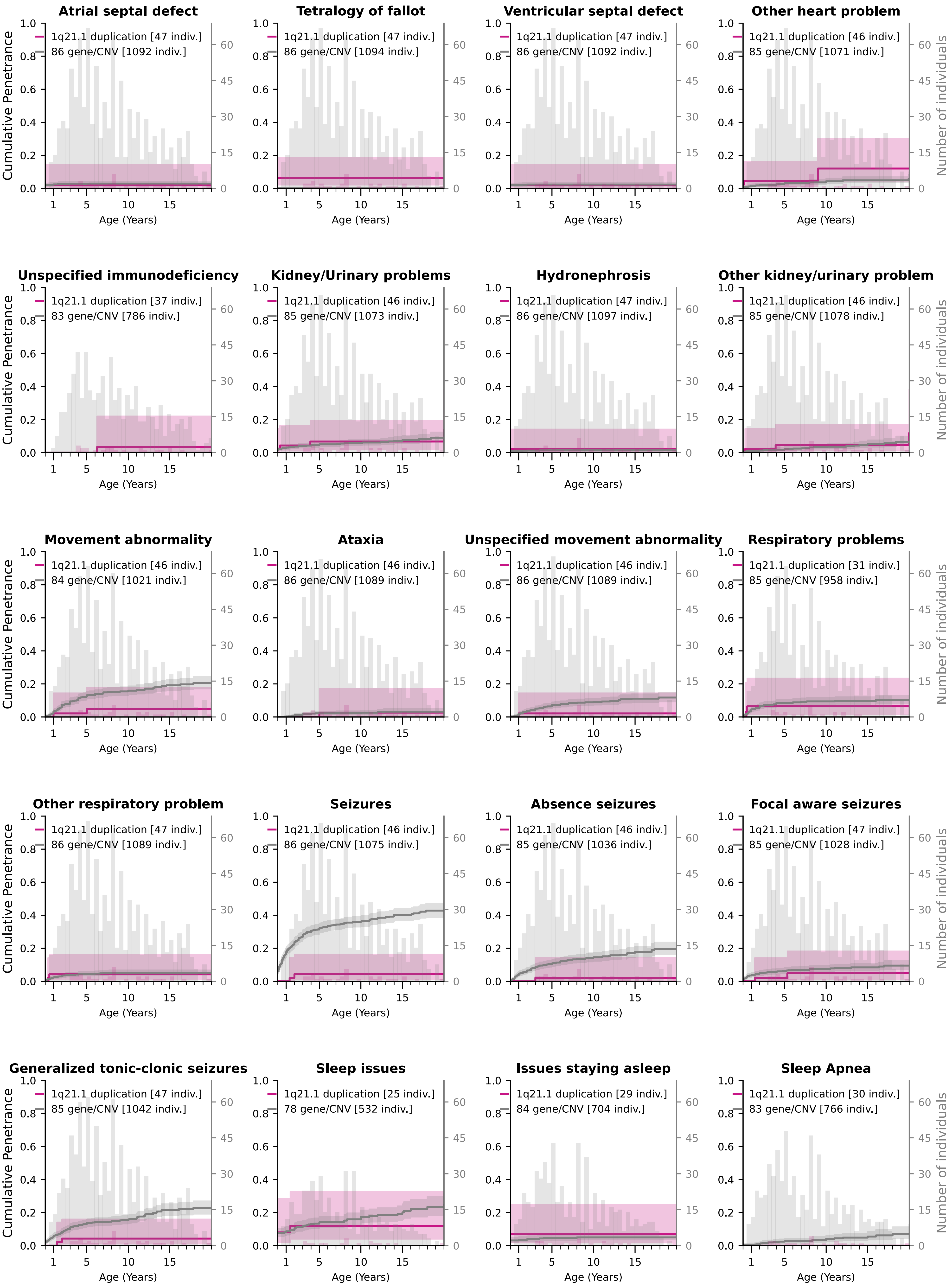

### Cumulative Penetrance for 1q21.1 duplication (page 4/4)

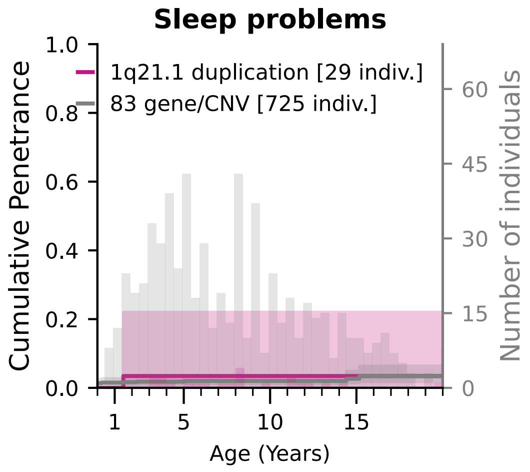

### Cumulative Penetrance for 16p11.2 deletion (page 1/4)

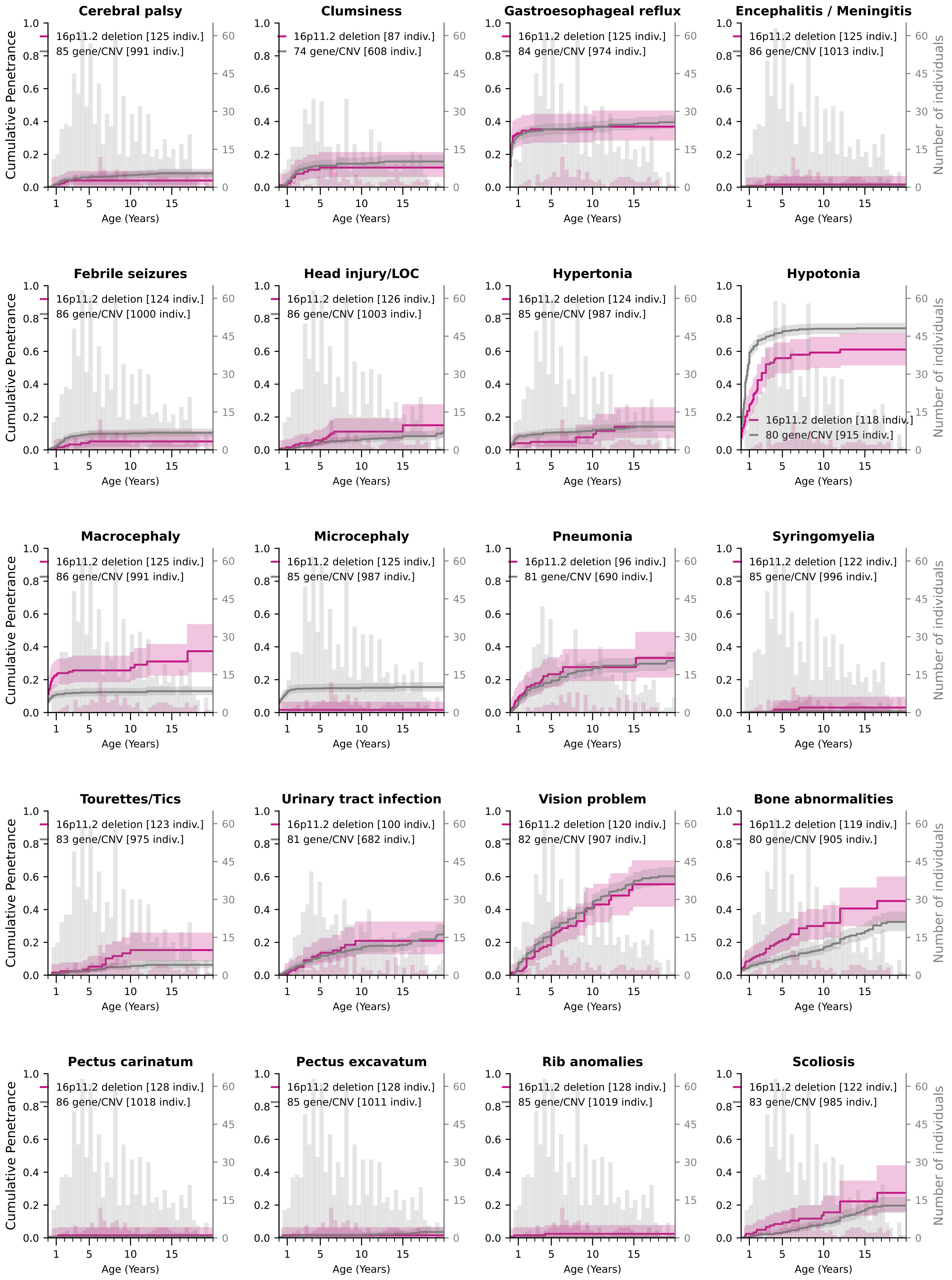

### Cumulative Penetrance for 16p11.2 deletion (page 2/4)

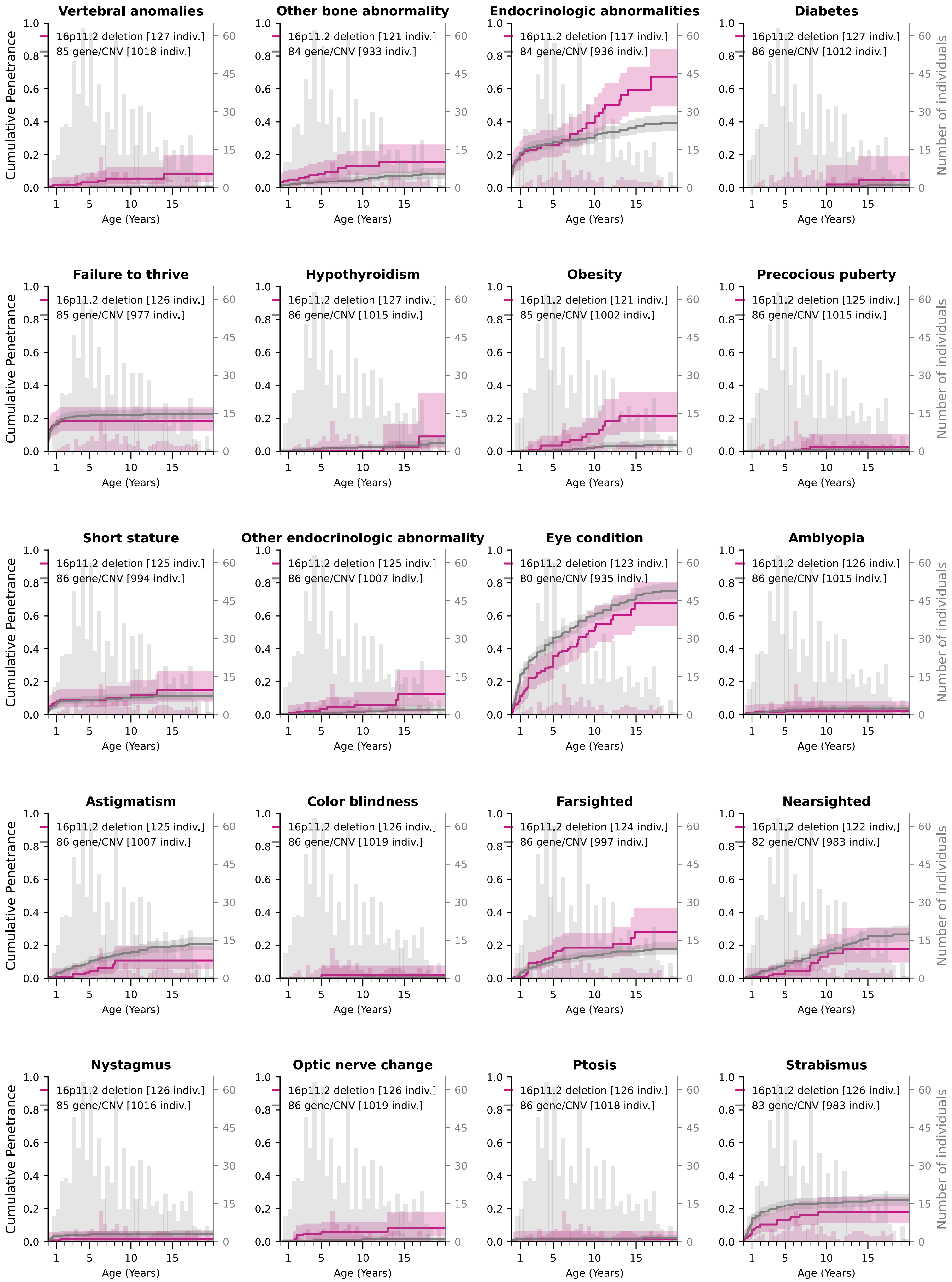

### Cumulative Penetrance for 16p11.2 deletion (page 3/4)

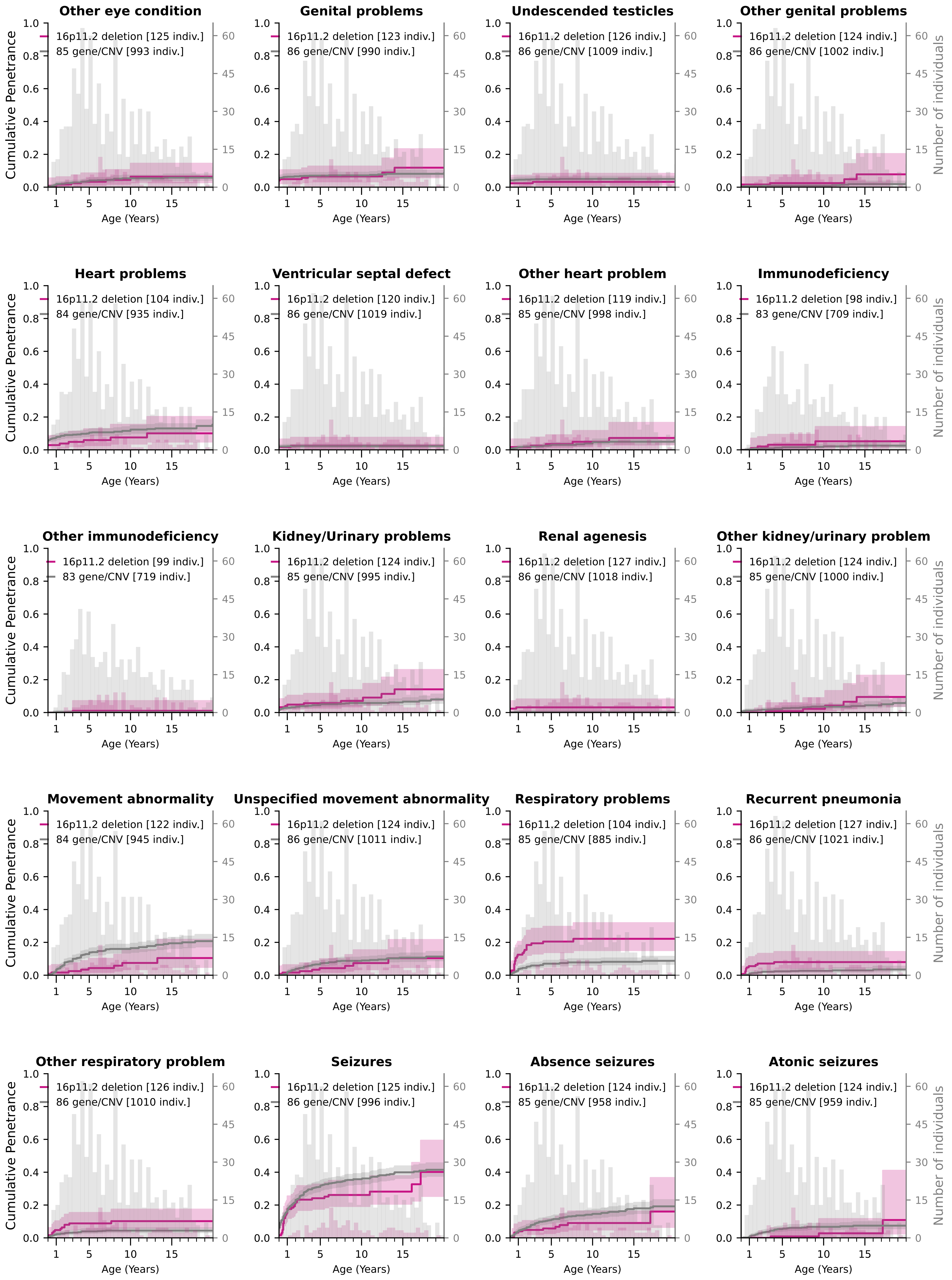

### Cumulative Penetrance for 16p11.2 deletion (page 4/4)

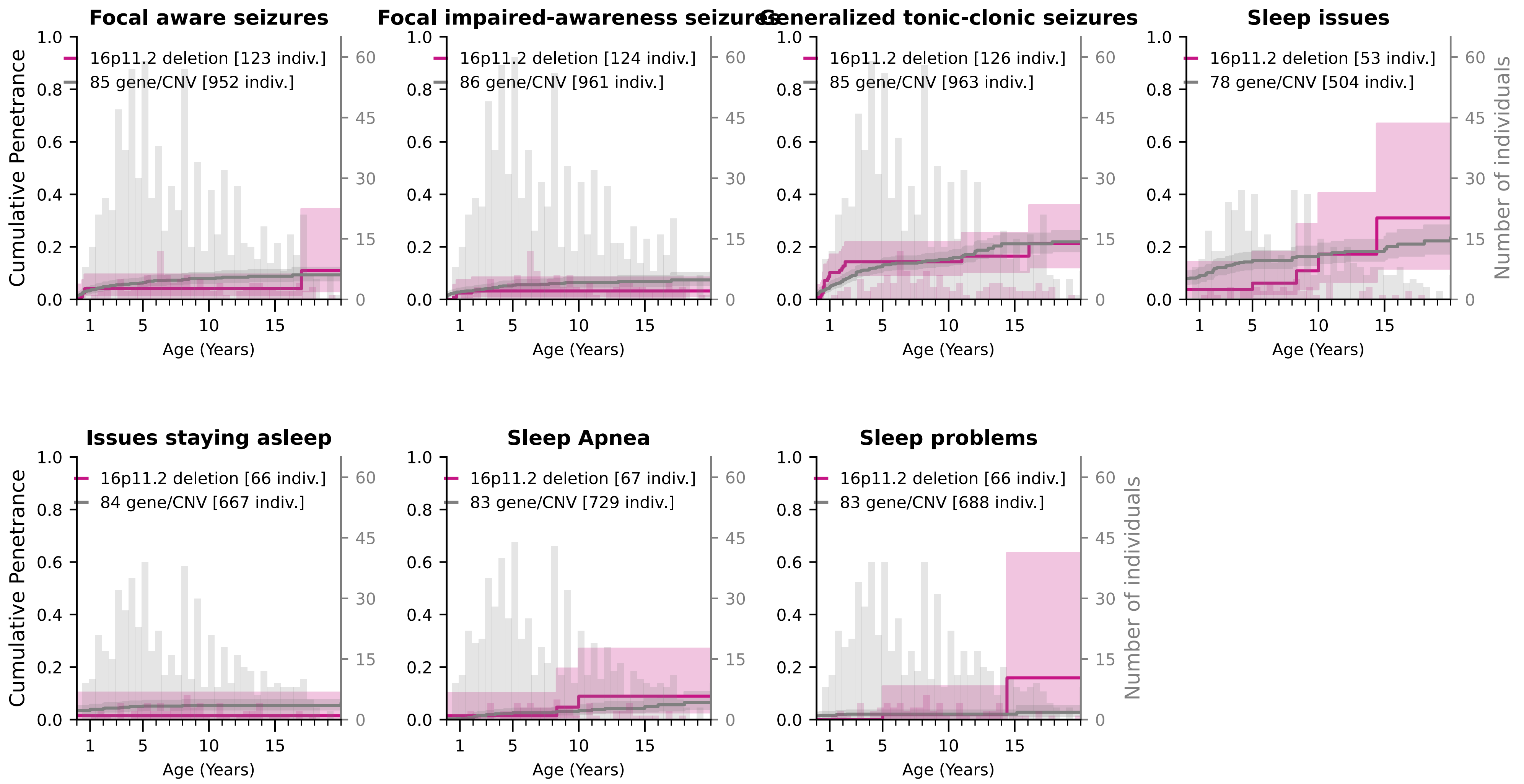

### Cumulative Penetrance for 16p11.2 duplication (page 1/4)

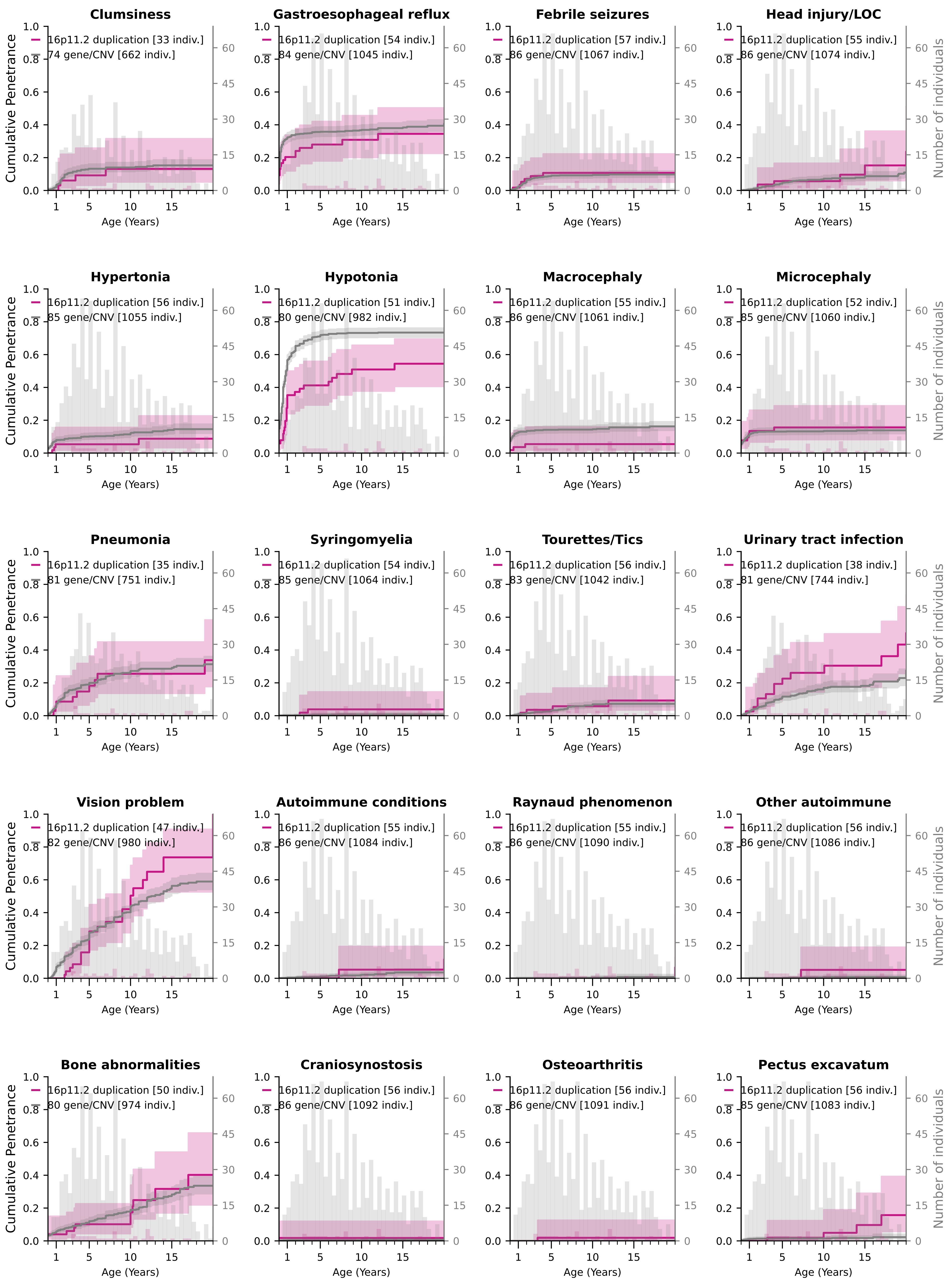

### Cumulative Penetrance for 16p11.2 duplication (page 2/4)

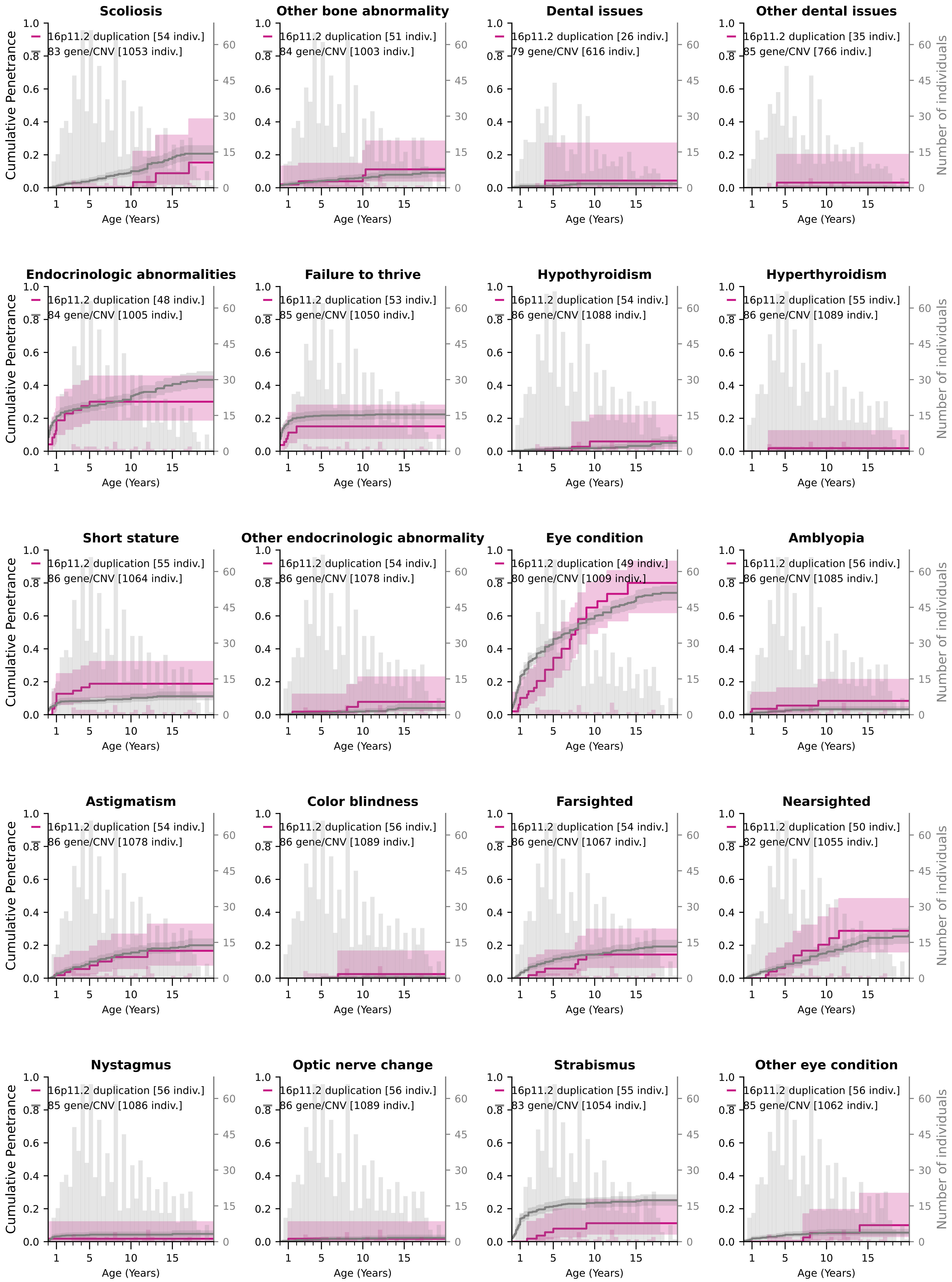

### Cumulative Penetrance for 16p11.2 duplication (page 3/4)

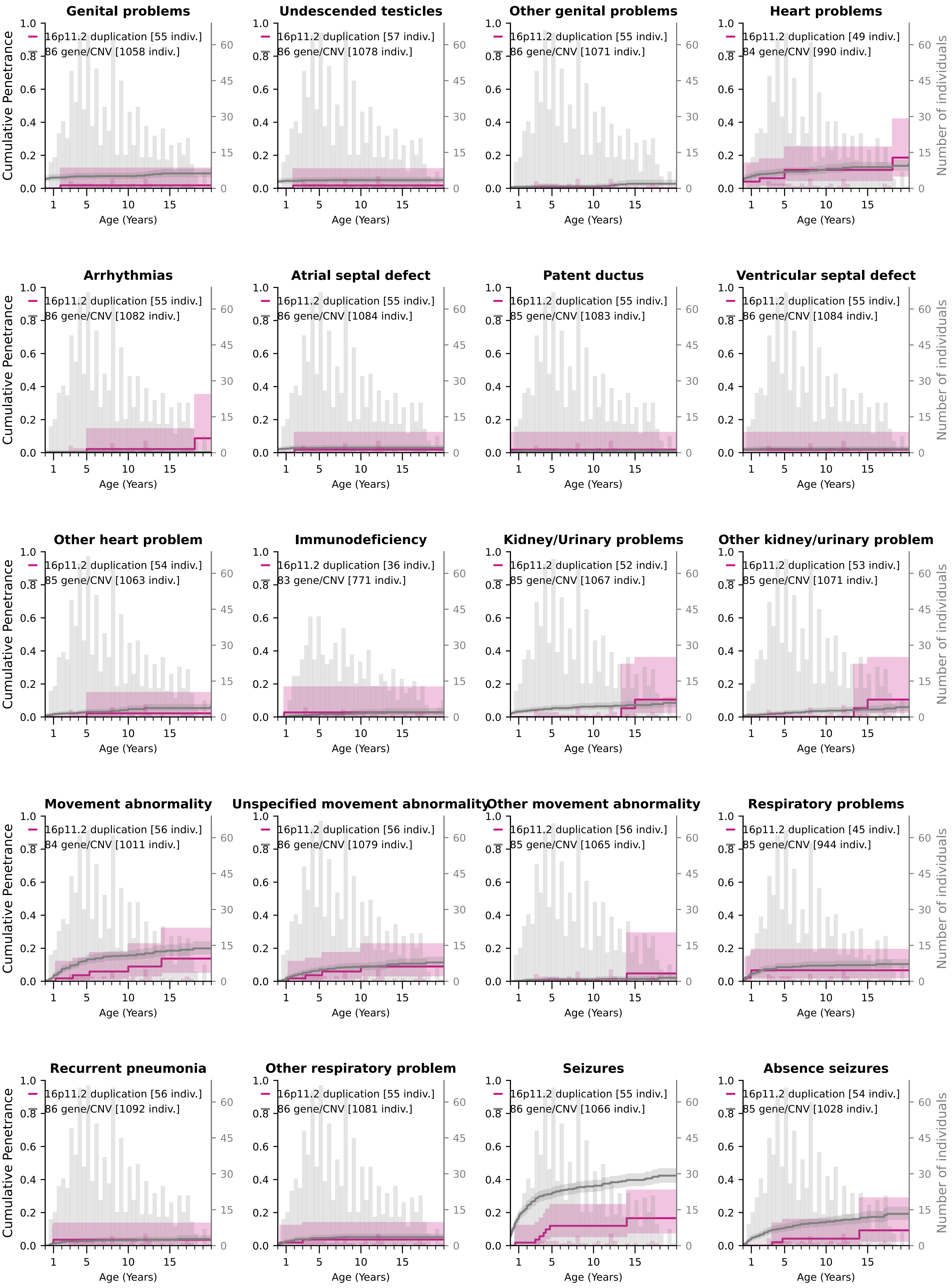

### Cumulative Penetrance for 16p11.2 duplication (page 4/4)

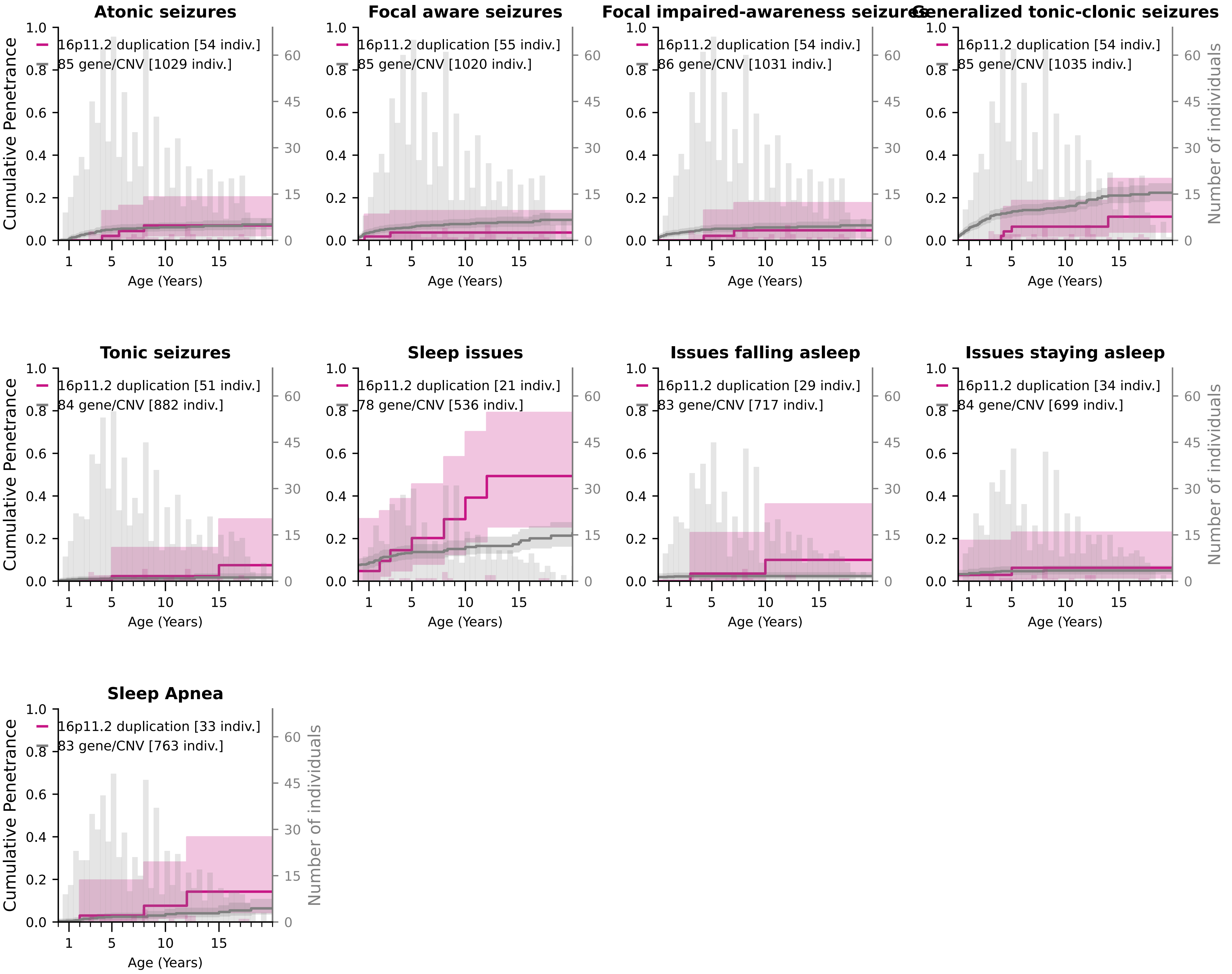
